# Place, gender, and uneven progress in pediatric and adolescent HIV across sub-Saharan Africa: a regional meta-analytic assessment (2000–2024)

**DOI:** 10.64898/2025.12.29.25343147

**Authors:** Chigozie Louisa J. Ugwu, Woldegebriel Assefa Woldegerima, Nicola Luigi Bragazzi

## Abstract

**Background:** Marked declines in pediatric HIV have been achieved across sub-Saharan Africa, yet progress among adolescents remains uneven and strongly patterned by place. Drawing on place-based perspectives in health geography, we conceptualize Eastern and Southern Africa (ESA) and West and Central Africa (WCA) not merely as epidemiological regions, but as distinct assemblages of health systems, gender norms, and historical investment trajectories shaping HIV risk and care. Using UNICEF-harmonized indicators from 2000 to 2024, we examined how place structures progress toward the 2030 Sustainable Development Goal (SDG 3.3) target.

**Methods:** We pooled country-level estimates using inverse-variance random-effects meta-analysis (restricted maximum likelihood for *τ* ^2^ with Hartung–Knapp adjustment). HIV incidence and AIDS-related mortality were analysed on the log scale, and mother-to-child transmission (MTCT) on the logit scale. Outcomes were pooled by region, age group (0–14; 15–19), sex, and year. We quantified percentage change from 2010 to 2024, the achieved annualized rate of change (ARC), and the required ARC from 2024 to 2030 to achieve a 90% reduction from 2010 levels. Robustness was assessed using prespecified sensitivity analyses.

**Findings:** Between 2010 and 2024, child HIV incidence declined by 72% in ESA and 66% in WCA, while adolescent incidence declined by 54% and 62%, respectively. In 2024, MTCT remained above elimination thresholds in both regions (9.9% in ESA; 17.6% in WCA). Adolescent girls experienced substantially higher HIV incidence than boys, with pooled female-to-male incidence rate ratios of 4.13 (95% CI: 3.32–5.13) in ESA and 4.92 (4.04–5.99) in WCA. Despite progress, achieved declines among adolescents (ARC: −5.4%/year in ESA; −6.6%/year in WCA) fall well short of the acceleration required to meet 2030 targets.

**Conclusion:** HIV progress among children and adolescents in sub-Saharan Africa is deeply place-dependent. While ESA reflects the benefits of earlier and sustained health system investments, persistent structural and gendered vulnerabilities continue to constrain adolescent outcomes, particularly in WCA. Achieving SDG 3.3 will require place-responsive strategies that integrate gender-sensitive adolescent prevention, differentiated care, and strengthened PMTCT within the specific social, political, and health system contexts shaping risk and access.

**Highlights:**

- HIV declines among children and adolescents show strong place-based patterning.
- Child HIV incidence declined faster in ESA than in WCA, 2010–2024.
- Adolescent girls had four- to five-fold higher HIV incidence than boys.
- MTCT remains above elimination thresholds, especially in West and Central Africa.
- Meeting 2030 targets requires accelerated, place-responsive strategies.

## 1. Introduction

Ending AIDS as a public health threat by 2030 (SDG 3.3) depends not only on biomedical advances, but on how health risks, prevention, and care are shaped by place. In sub-Saharan Africa, home to nearly 86% of the world’s children and adolescents living with HIV, geographic context continues to structure unequal exposure, access to services, and epidemiological trajectories [1, 2, 3]. In 2024, an estimated 2.42 million children and adolescents (0–19 years) were living with HIV globally, with approximately 712 new pediatric infections and 250 AIDS-related deaths occurring each day [1, 4, 5]. These patterns underscore that HIV outcomes among younger populations remain deeply embedded in place-specific health systems, social relations, and political economies.

Despite substantial declines since 2010, progress has slowed unevenly. Adolescents, particularly adolescent girls and young women (AGYW), have benefited less from overall epidemic control, accounting for a growing share of new infections [6, 7]. Gendered spatial vulnerability, shaped by schooling environments, labor markets, household power relations, and health service accessibility, places adolescent girls at heightened risk in many settings [8, 9]. These vulnerabilities are not uniformly distributed, but vary systematically across regions and countries, reflecting broader geographies of inequality.

Regional contrasts within sub-Saharan Africa further illustrate the importance of place. Eastern and Southern Africa (ESA) has achieved faster epidemiological declines and broader antiretroviral treatment (ART) coverage, while West and Central Africa (WCA) continues to lag across multiple indicators [1, 4]. In 2024, pediatric ART coverage in WCA remained around one-third on average, substantially lower than in ESA. Although prevention of mother-to-child transmission (PMTCT) has expanded across both regions, MTCT rates remain above World Health Organization validation thresholds in many countries, particularly in WCA [10, 11]. These disparities reflect historically uneven investments in health infrastructure, supply chains, workforce capacity, and governance, core elements of the regional political economy of HIV response.

Existing comparative studies often aggregate pediatric and adult populations or provide limited age- and sex-disaggregated analyses, obscuring how adolescent disparities intersect with place [12, 13, 14]. While adolescent incidence differentials by sex are well documented, few studies formally synthesize uncertainty across countries or quantify the pace of change required to meet SDG 3.3 benchmarks [9, 15]. As a result, it remains unclear how much acceleration is needed, and where, to realign trajectories toward elimination.

In this study, we explicitly foreground place as a structural determinant of pediatric and adolescent HIV outcomes. We conceptualize ESA and WCA not merely as epidemiological regions, but as distinct place-based assemblages of health systems, gender norms, and historical investment patterns that shape exposure, prevention, and care. Using UNICEF-harmonized indicators from 2000 to 2024, we: (i) trace regional trajectories in HIV incidence, AIDS-related mortality, and MTCT by age and sex; (ii) estimate the achieved annualized rate of change (2010–2024) and the required rate of change (2024–2030) to meet SDG 3.3 targets; and (iii) meta-analyse adolescent female-to-male incidence rate ratios to quantify gendered spatial inequality. By linking epidemiological trends to place-based structures, this analysis provides policy-relevant benchmarks for designing context-sensitive strategies to accelerate progress among children and adolescents across sub-Saharan Africa.

## 2. Materials and methods

### 2.1. Data source and study design

We conducted a comparative, place-based regional synthesis of pediatric and adolescent HIV indicators using publicly available country-year estimates curated by UNICEF and produced from UNAIDS 2025 Spectrum (AIDS Impact Model; AIM) outputs [16, 17, 18]. Spectrum estimates are generated annually using multiple data inputs (e.g., programme monitoring via Global AIDS Monitoring, population-based surveys such as DHS/MICS/PHIA, surveillance systems, and vital registration where available) [19, 20]. We treat countries as analytically meaningful places in which health system capacity, gendered risk environments, and historical investment patterns shape HIV risk and service access.

### 2.2. Geographic scope and study population

We restricted analyses to UNICEF regions, Eastern and Southern Africa (ESA) and West and Central Africa (WCA), using UNICEF’s regional taxonomy [16]. To avoid double-counting, we used only country-level records. Study populations were: (i) children aged 0–14 years and (ii) adolescents aged 15–19 years, each stratified by sex, (Sex = Female, Male, Both) when available. For mother-to-child transmission, the UNICEF reports MTCT as (Age = Age 0-4) and (Sex = Both); we analysed MTCT using this definition and label it explicitly as (MTCT (0–4) throughout. For mortality, the data also includes (Age = Age 10-19); to maintain consistency with adolescent reporting in this paper, we restricted mortality to (Age 0–14 and Age 15–19). Eligible observations were all country-years in 2000–2024, missing or suppressed values were treated as missing and were not imputed.

### 2.3. Outcomes

We analysed three primary outcomes: (i) Estimated incidence rate (new HIV infections per 1,000 uninfected population) for ages 0–14 and 15–19; (ii) Estimated AIDS-related mortality rate (per 100,000 population) for ages 0–14 and 15–19; and (iii) Estimated MTCT rate (%) for age 0–4 (Sex = Both). All outcomes were accompanied by 95% uncertainty bounds (Lower, Upper) used to derive sampling variances for pooling.

### 2.4. Data preparation and quality assurance

Analytic keys were standardised using ISO3, Indicator, Year, Sex, Age, and UNICEF Region. The source data contain left-censored values encoded as character strings (e.g., <0.01) in the estimate and uncertainty bounds (Value, Lower, and Upper). For reproducibility and to permit log- and logit-scale transformations, any entry of the form *< t* was deterministically converted to *t/*2 (e.g., *<* 0.01 7→ 0.005) prior to analysis. Original raw entries were retained alongside the numeric values in the exported analytic dataset to enable full auditability.

Data quality checks were applied uniformly across all indicators. Implausible values were excluded, defined as negative incidence or mortality rates and mother-to-child transmission (MTCT) proportions outside the interval [0, 100]%. We further verified that uncertainty bounds satisfied Upper ≥ Lower. Where duplicate records occurred for the same analytic key (ISO3, Indicator, Age, Sex, Year), a single internally consistent record was retained. No additional temporal smoothing, interpolation, or model-based adjustment was applied beyond the UNAIDS Spectrum estimation process embedded in the source series.

### 2.5. Statistical analysis: pooling across places and quantifying between-place variation

Within each stratum defined by region × year × age × sex, we treated each country-year estimate as an effect size with an associated sampling variance. This approach is analogous to study-level meta-analysis, but here the unit is *place*: it summarises the regional distribution of country-specific levels while allowing for real between-place differences driven by health systems, policy contexts, and gendered vulnerability.

Incidence and mortality rates were analysed on the natural-log scale; MTCT percentages were analysed on the logit scale to map [0, 1] → ¡ and stabilise variance [21]. Standard errors were derived from 95% uncertainty bounds:

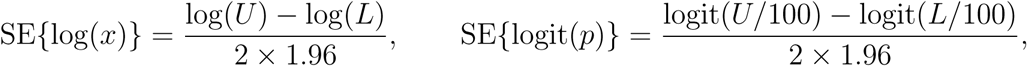

where [*L, U* ] are the bounds and *p* is a percentage. To ensure finite transformations, inputs were truncated away from boundaries: for log-scale outcomes, values were bounded below by 10^−6^; for logits, *p* was truncated to (10^−6^, 100 − 10^−6^).

Country effects were pooled using inverse-variance random-effects meta-analysis with restricted maximum likelihood (REML) estimation for *τ* ^2^ and Hartung–Knapp adjusted confidence intervals, implemented in `metafor (rma.uni, method=REML, test=knha)` [22, 23, 24]. We report back-transformed pooled estimates with 95% confidence intervals, along with *τ* ^2^ and *I*^2^ to summarise between-place heterogeneity [25]. If REML failed to converge, Paule–Mandel was used; if this failed, DerSimonian–Laird was applied. Strata with *k <* 2 countries were not pooled.

### 2.6. Adolescent sex disparity

For ages 15–19 in 2024, we quantified adolescent sex disparity by estimating country-specific female:male log-incidence ratios,

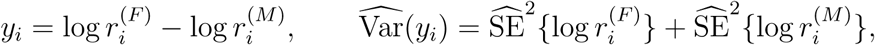

and pooled *y_i_* within ESA and WCA using the same REML+Hartung–Knapp model. Exponentiated results are presented as incidence rate ratios (IRRs) with 95% confidence intervals.

### 2.7. Progress metrics: achieved and required acceleration to 2030

We quantified progress from 2010 to 2024 using the achieved annualised rate of change (ARC),

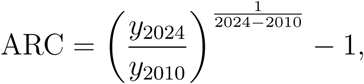

and the required ARC from 2024 to 2030 to reach a 90% reduction from the 2010 level,

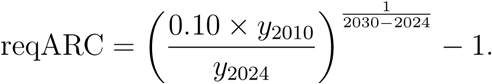

ARC and reqARC were computed using balanced panels (countries observed in both 2010 and 2024 within each stratum) to ensure consistent place composition.

### 2.8. Sensitivity analyses

To assess robustness to panel composition, influential countries, and heterogeneity estimation, we conducted prespecified checks: (i) balanced-panel re-estimation; (ii) leave-one-country-out refits; (iii) alternative *τ* ^2^ estimators (Paule–Mandel, DerSimonian–Laird); (iv) 95% prediction intervals for 2024 on the transformed scale to quantify expected between-place dispersion; and (v) parametric bootstrap intervals for ARC and reqARC (*B* = 5,000 draws) using normal approximations to pooled 2010 and 2024 effects on the meta-analytic scale, followed by back-transformation.

All analyses were conducted in R 4.4.1 using `readxl, dplyr, tidyr, ggplot2`, and `metafor` [26, 22]. The study used aggregated, publicly available estimates and did not involve individual-level human subjects data; therefore, ethics approval was not required. To ensure full reproducibility, the exact analytic dataset used in this study and the R scripts required to reproduce all analyses and figures are made available by the corresponding author.

## 3. Results

### 3.1. Overview: declines with persistent between-place inequality

Across 2000–2024, pooled regional trends indicate substantial declines in HIV incidence and AIDS-related mortality in both Eastern and Southern Africa (ESA) and West and Central Africa (WCA) (Figures 1 and 2). Using 2010 as the benchmark year and 2024 as the most recent observation, HIV incidence among children (0–14 years) declined by 72.4% in ESA and 65.7% in WCA, while adolescent incidence (15–19 years) declined by 53.9% in ESA and 61.4% in WCA (Table 1). AIDS-related mortality also fell markedly (Table 2), with larger proportional reductions among children than adolescents.

**Figure 1:**
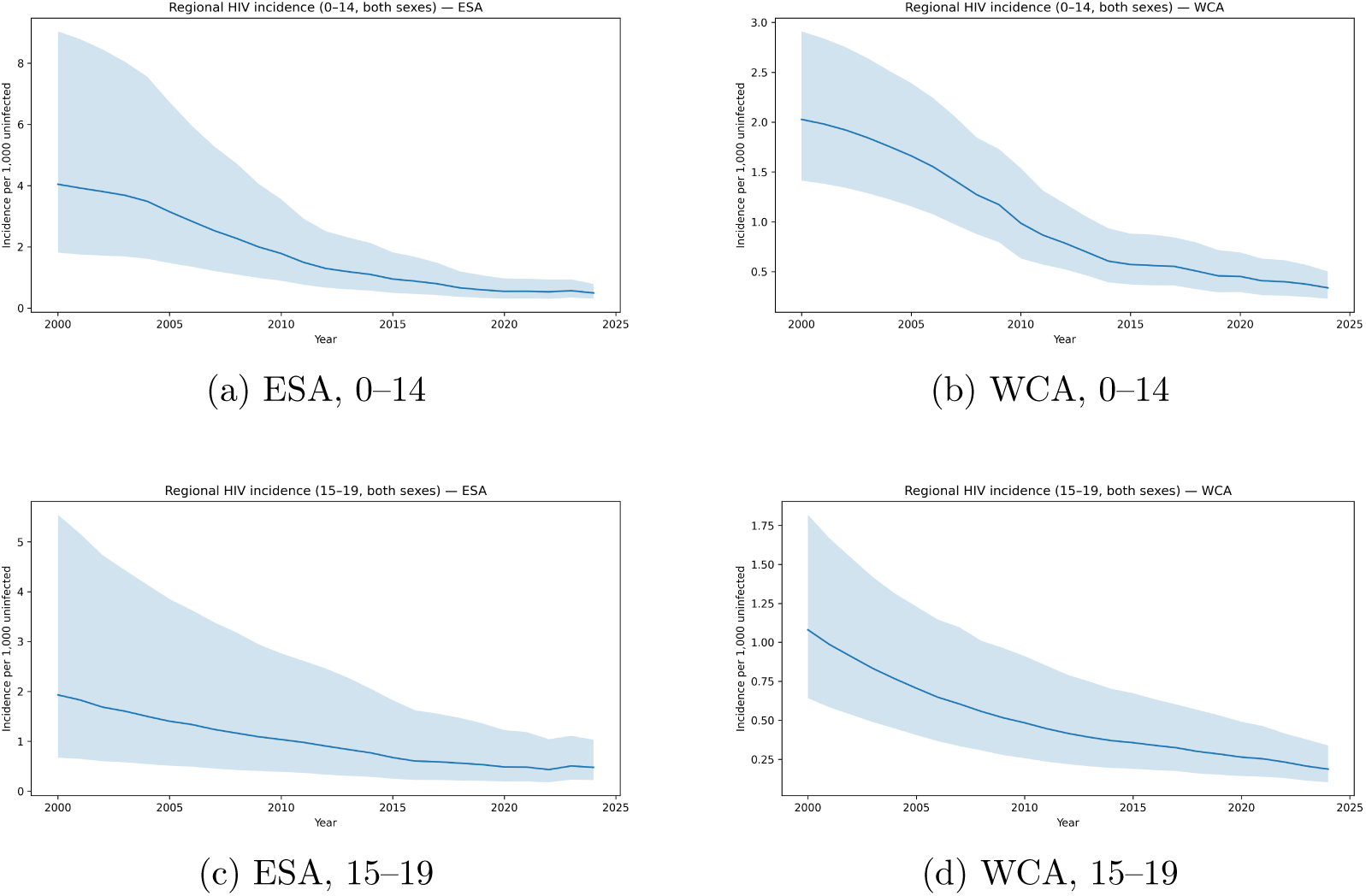
HIV incidence per 1,000 uninfected population: pooled regional trends with 95% confidence intervals, 2000–2024 (random-effects meta-analysis, REML with Hartung–Knapp).

**Figure 2:**
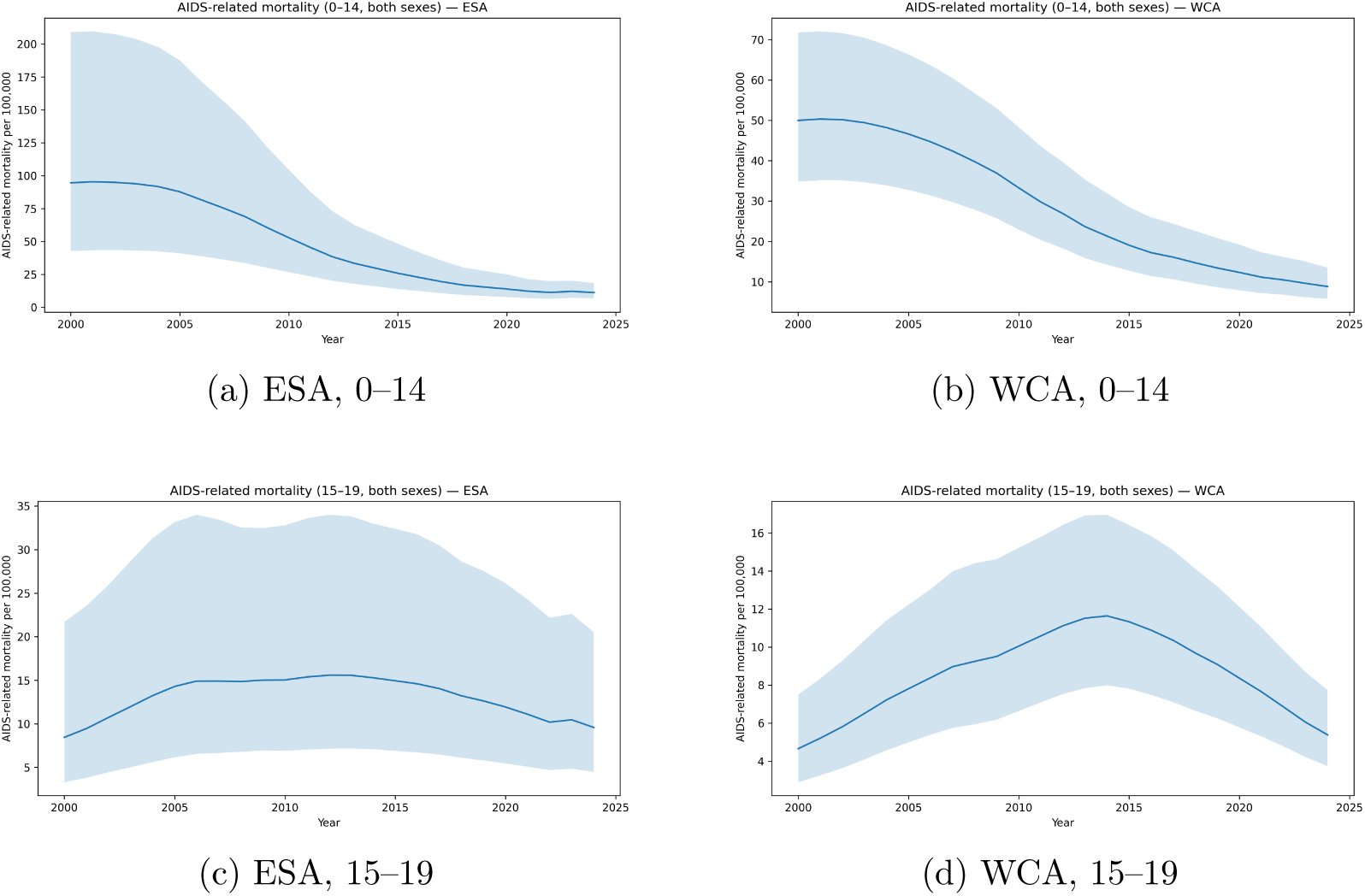
AIDS-related mortality per 100,000 population: pooled regional trends with 95% confidence intervals, 2000–2024 (random-effects meta-analysis, REML with Hartung–Knapp).

**Table 1:** Regional pooled HIV incidence per 1,000 uninfected population in Eastern and Southern Africa (ESA) and West and Central Africa (WCA), 2010 and 2024, with percent change and annualized rate of change (ARC).

| Region | Age group | 2010 | 95% CI | $\tau^2_{2010}$ | $I^2_{2010}$ (%) | 2024 | 95% CI | $\tau^2_{2024}$ | $I^2_{2024}$ (%) | Change (%) | ARC (%/year) | $k_{2010}$ | $k_{2024}$ |
| --- | --- | --- | --- | --- | --- | --- | --- | --- | --- | --- | --- | --- | --- |
| ESA | 0–14 years | 1.784 | [0.894, 3.560] | 2.407 | 99.5 | 0.493 | [0.308, 0.787] | 0.980 | 97.2 | -72.4 | -8.8 | 22 | 21 |
| ESA | 15–19 years | 1.035 | [0.388, 2.762] | 4.938 | 96.0 | 0.478 | [0.221, 1.032] | 2.804 | 93.5 | -53.9 | -5.4 | 23 | 22 |
| WCA | 0–14 years | 0.986 | [0.632, 1.539] | 0.882 | 98.8 | 0.338 | [0.228, 0.502] | 0.656 | 95.1 | -65.7 | -7.4 | 20 | 20 |
| WCA | 15–19 years | 0.484 | [0.257, 0.911] | 1.727 | 91.6 | 0.186 | [0.103, 0.339] | 1.525 | 89.0 | -61.4 | -6.6 | 21 | 21 |

*Notes:* Estimates are pooled using random-effects meta-analysis with restricted maximum likelihood (REML) and
Hartung–Knapp adjustment. $\tau^2$ denotes between-country variance on the log-incidence scale, and $I^2$ quantifies the proportion of total variability attributable to between-place heterogeneity rather than sampling error. $k$ indicates the number of contributing countries in each year.

However, large between-country heterogeneity persisted in every year and stratum (high *I*^2^ and non-zero *τ* ^2^), indicating that observed regional averages mask substantial *between-place* variation in epidemic burden and program performance. In both regions, adolescent girls experienced higher incidence than boys throughout the series (Figure 3), consistent with gendered and spatially uneven exposure, prevention access, and treatment continuity. Despite progress, observed declines remain insufficient to meet the 2030 target of reducing key indicators to 10% of their 2010 levels without accelerated and geographically targeted action (Section 3.7).

**Figure 3:**
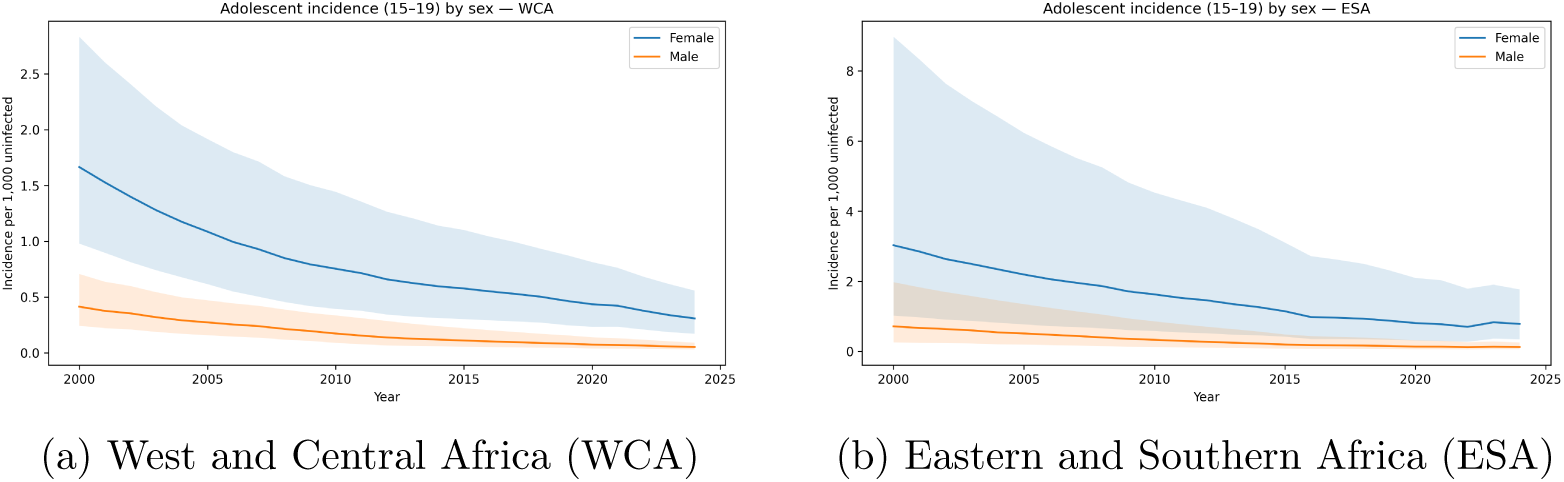
HIV incidence among adolescents aged 15–19 years by sex, 2000–2024. Solid lines show pooled regional estimates per 1,000 uninfected population; shaded bands indicate 95% uncertainty intervals derived from random-effects meta-analysis (REML with Hartung–Knapp adjustment). Across both regions and throughout the study period, incidence remained consistently higher among females than males, while substantial between-country heterogeneity persists within each region.

### 3.2. HIV incidence (0–14 and 15–19 years)

Regional pooled incidence declined steadily from 2000–2024 in both age groups and regions (Figure 1). In ESA, child incidence (0–14) decreased from 1.784 per 1,000 uninfected in 2010 to 0.493 in 2024 (72.4% reduction; ARC−8.8%/year), while adolescent incidence (15–19) declined from 1.035 to 0.478 (53.9% reduction; ARC −5.4%/year) (Table 1). In WCA, child incidence declined from 0.986 to 0.338 (65.7% reduction; ARC −7.4%/year) and adolescent incidence from 0.484 to 0.186 (61.4% reduction; ARC −6.6%/year).

While regional trajectories suggest broad progress, country-level distributions in 2024 reveal pronounced dispersion within each region (Figure 3). This heterogeneity indicates that *place* functions as a fundamental structuring context for adolescent HIV risk: a small subset of countries contributes disproportionately to the upper tail of incidence, while many countries cluster near comparatively low rates. These patterns underscore the need for place-differentiated prevention and treatment strategies rather than uniform regional programming.

### 3.3. Between-place heterogeneity in adolescent incidence (2024)

Forest plots for adolescent HIV incidence in 2024 (ages 15–19, both sexes) show long right tails in both Eastern and Southern Africa (ESA) and West and Central Africa (WCA) (Figure 4). In ESA, the upper tail is driven by a small group of countries with clearly elevated incidence relative to the pooled regional mean; in WCA, overall levels are lower, but between-place dispersion remains pronounced. These tail-driven distributions indicate that regional averages underestimate the degree of localized vulnerability and may obscure priority geographies for intervention.

**Figure 4:**
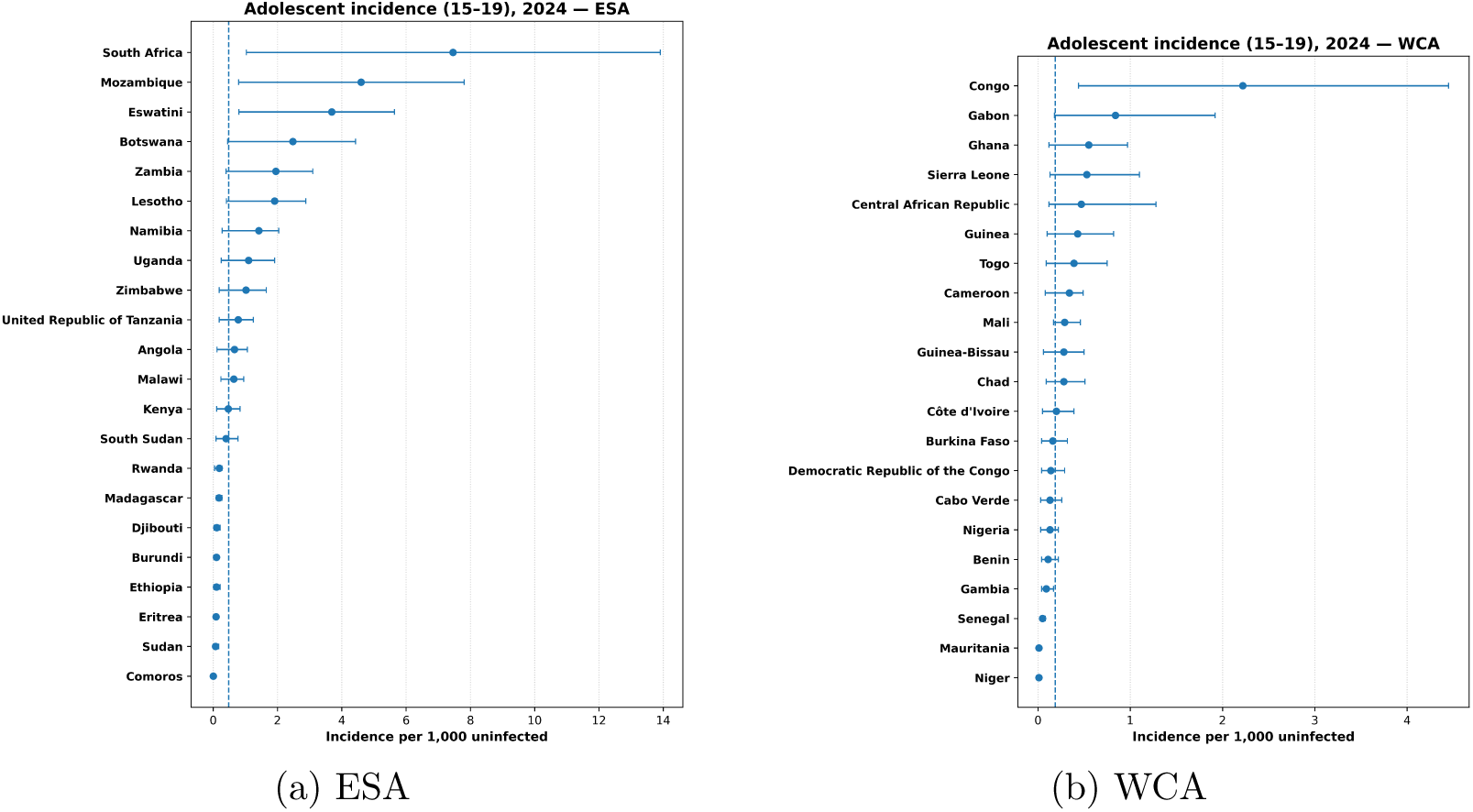
Country-level random-effects forest plots of adolescent HIV incidence (ages 15–19), 2024. Points represent country-specific estimates with 95% confidence intervals; the dashed vertical line indicates the pooled regional mean derived from random-effects meta-analysis (REML with Hartung–Knapp adjustment).

To make the place signal explicit, Figure 5 ranks the top 10 countries by adolescent incidence in each region in 2024. This ranking highlights the spatial concentration of risk and provides a policy-relevant lens for identifying where accelerated prevention and adolescent-friendly services are most urgently needed.

**Figure 5:**
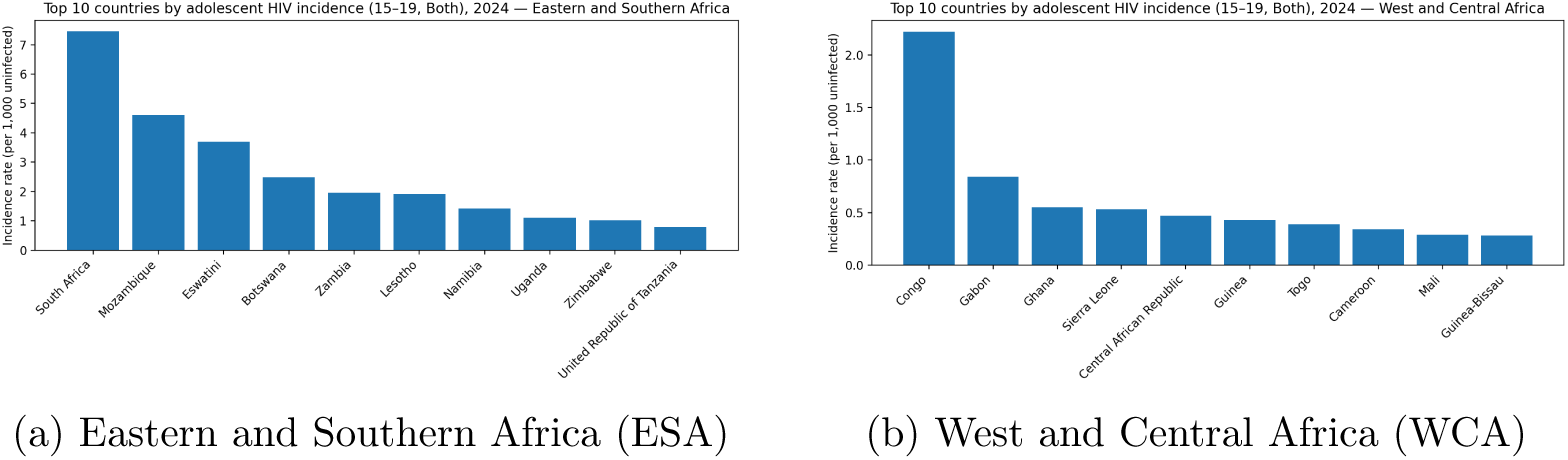
Top 10 countries by adolescent HIV incidence (ages 15–19, both sexes), 2024, by region. Bars show country-specific incidence rates per 1,000 uninfected population, derived from UNAIDS Spectrum estimates and ranked within each UNICEF region. The figure illustrates the spatial concentration of adolescent HIV risk, with a small number of countries accounting for the upper tail of incidence in each region.

Together with the forest meta-analyses (Figure 4), these rankings demonstrate that regional averages obscure substantial within-region inequality driven by a limited number of high-burden places.

### 3.4. AIDS-related mortality (0–14 and 15–19 years)

AIDS-related mortality declined substantially between 2000 and 2024, with faster improvements among children than adolescents (Figure 2). In ESA, mortality among children declined from 52.93 per 100,000 in 2010 to 11.30 in 2024 (78.7% reduction; ARC −10.4%/year), while adolescent mortality fell from 15.03 to 9.58 (36.3% reduction; ARC −3.2%/year) (Table 2). In WCA, child mortality declined from 33.26 to 8.82 (73.5% reduction; ARC −9.0%/year) and adolescent mortality from 10.05 to 5.39 (46.4% reduction; ARC −4.4%/year).

**Table 2:** AIDS-related mortality per 100,000: pooled 2010 and 2024 levels, percent change, and annualized rate of change (ARC).

| Region | Age | 2010 | 95% CI | $\tau_{2010}^2$ | $I_{2010}^2$ (%) | 2024 | 95% CI | $\tau_{2024}^2$ | $I_{2024}^2$ (%) | $\Delta\%$ | ARC (%/yr) | $k_{2010}$ | $k_{2024}$ |
| --- | --- | --- | --- | --- | --- | --- | --- | --- | --- | --- | --- | --- | --- |
| ESA | 0–14 | 52.93 | [26.80, 104.54] | 2.34 | 99.6 | 11.30 | [6.93, 18.43] | 1.08 | 96.2 | -78.7 | -10.4 | 22 | 21 |
| ESA | 15–19 | 15.03 | [6.89, 32.78] | 3.20 | 99.7 | 9.58 | [4.47, 20.51] | 2.81 | 99.5 | -36.3 | -3.2 | 23 | 22 |
| WCA | 0–14 | 33.26 | [22.90, 48.30] | 0.66 | 99.0 | 8.82 | [5.76, 13.49] | 0.83 | 95.9 | -73.5 | -9.0 | 21 | 21 |
| WCA | 15–19 | 10.05 | [6.64, 15.22] | 0.86 | 98.7 | 5.39 | [3.75, 7.74] | 0.64 | 96.1 | -46.4 | -4.4 | 22 | 22 |

Notes: Random-effects meta-analysis via REML with Hartung–Knapp adjustment. $\tau^2$ is reported on the log-mortality scale; larger values indicate greater between-country dispersion. $k$ denotes the number of contributing countries.

Despite overall progress, persistently high heterogeneity indicates that mortality reductions remain spatially uneven, consistent with between-place differences in timely diagnosis, pediatric ART access, and continuity of care across health systems.

### 3.5. Adolescent sex disparities (15–19 years): standardized pooled IRR (2024)

In 2024, adolescent girls had substantially higher HIV incidence than boys in both regions (Figure 3). In ESA, pooled incidence was 0.786 per 1,000 uninfected for females and 0.127 for males; in WCA, pooled incidence was 0.308 for females and 0.052 for males.

To quantify the female excess risk consistently, we estimated country-specific log incidence ratios (female vs. male) and pooled them by region using random-effects meta-analysis (REML with Hartung–Knapp). The resulting pooled female:male IRR was 4.13 (95% CI 3.32–5.13) in ESA and 4.92 (95% CI 4.04–5.99) in WCA (Figure 6). These results show that gendered risk is not confined to a few settings but is a robust, region-wide feature, operating through spatially uneven social, economic, and service-delivery contexts.

**Figure 6:**
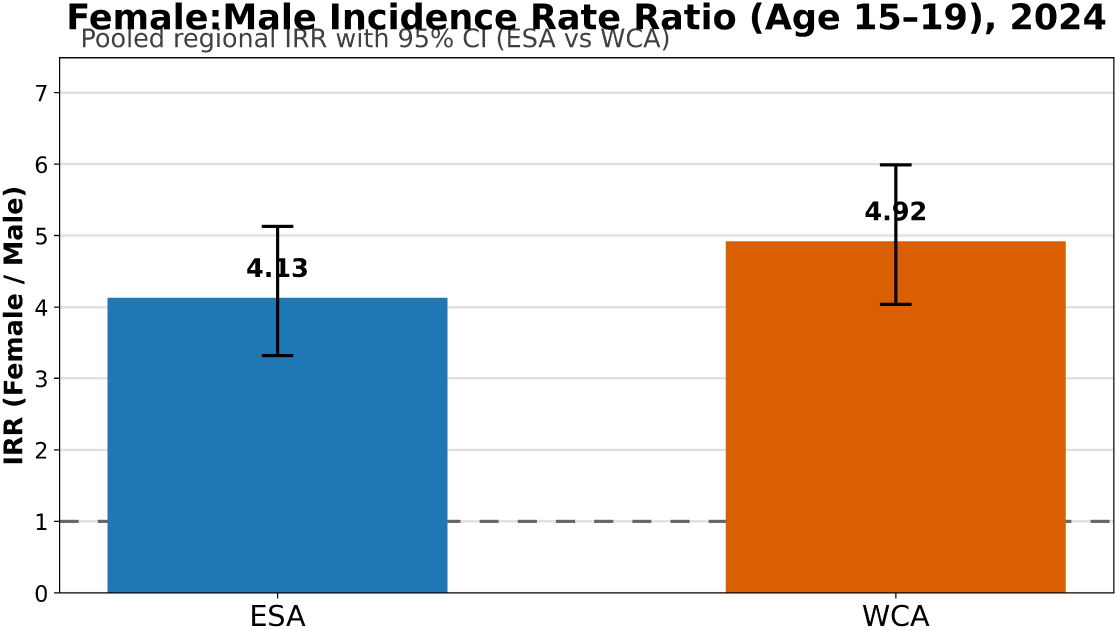
Pooled female-to-male HIV incidence rate ratios (IRR) among adolescents aged 15–19 years in 2024, by region. Estimates are derived from random-effects meta-analysis of country-specific log incidence rate ratios using restricted maximum likelihood with Hartung–Knapp adjustment. Error bars denote 95% confidence intervals; the dashed horizontal line marks IRR = 1 (no sex difference).

### 3.6. Mother-to-child transmission (MTCT)

Mother-to-child transmission (MTCT) declined markedly across both regions over the study period, with substantial improvements observed between the 2010 benchmark and 2024 (Figure 7; Table 3). In Eastern and Southern Africa (ESA), pooled MTCT decreased from 26.0% (95% CI: 20.3–32.6) in 2010 to 9.9% (6.1–15.6) in 2024, corresponding to a 61.9% reduction and an annualized rate of change (ARC) of −6.7% per year. In West and Central Africa (WCA), MTCT declined from 29.6% (26.3–33.1) to 17.6% (14.9–20.7), representing a smaller 40.5% reduction (ARC −3.6%/year), leaving MTCT levels in 2024 notably higher than in ESA.

**Figure 7:**
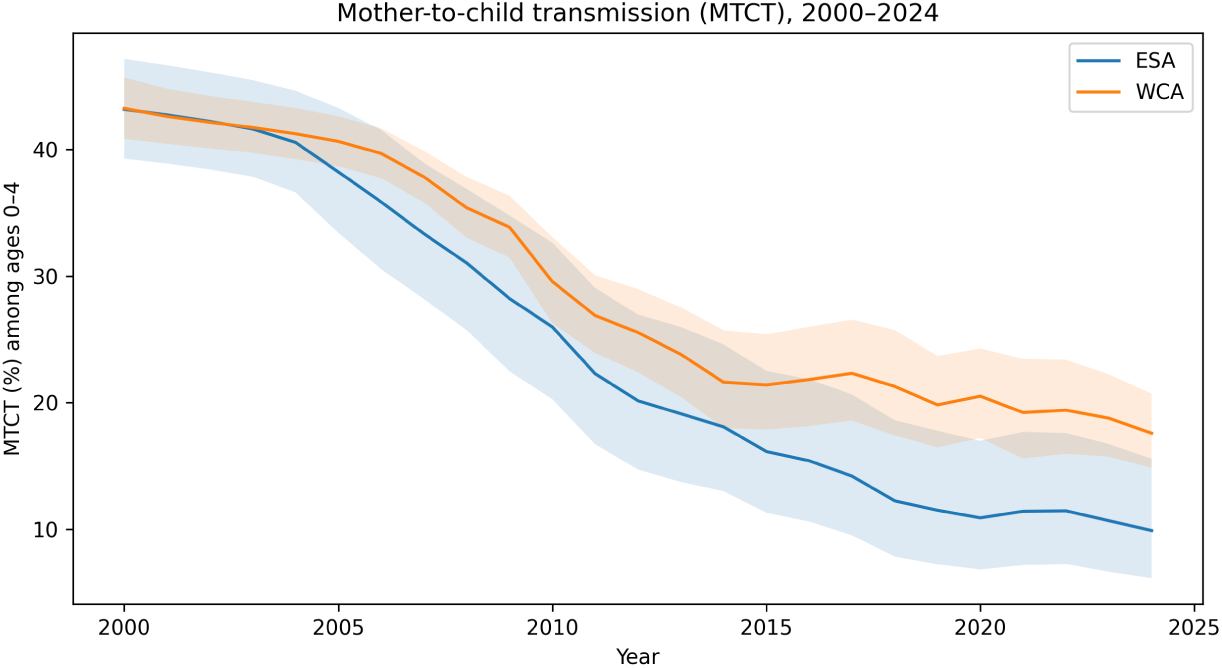
Mother-to-child transmission (MTCT) rate (%) among children aged 0–4 years, 2000–2024: pooled regional trends with 95% confidence intervals estimated using random-effects meta-analysis (REML with Hartung–Knapp adjustment).

**Table 3:** MTCT (%): pooled 2010 and 2024 levels (REML with Hartung–Knapp, logit scale), percent change, and annualized rate of change (ARC).

| Region | Age | 2010 | 95% CI | $\tau^2_{2010}$ | $I^2_{2010}$ (%) | 2024 | 95% CI | $\tau^2_{2024}$ | $I^2_{2024}$ (%) | $\Delta\%$ | ARC (%/yr) | $k_{2010}$ | $k_{2024}$ |
| --- | --- | --- | --- | --- | --- | --- | --- | --- | --- | --- | --- | --- | --- |
| ESA | Age 0–4 | 26.0 | [20.3, 32.6] | 0.487 | 99.5 | 9.9 | [6.1, 15.6] | 1.268 | 98.9 | -61.9 | -6.7 | 21 | 21 |
| WCA | Age 0–4 | 29.6 | [26.3, 33.1] | 0.114 | 98.0 | 17.6 | [14.9, 20.7] | 0.159 | 92.1 | -40.5 | -3.6 | 20 | 20 |

Notes: Random-effects meta-analysis via REML with Hartung–Knapp on the logit scale; values back-transformed to
percentages; $k$ = number of countries; $\tau^2$ on the logit scale.

Despite these gains, between-country heterogeneity remained substantial in both regions, as indicated by persistently high *τ* ^2^ and *I*^2^ values on the logit scale (Table 3). This heterogeneity demonstrates that progress toward MTCT elimination has been spatially uneven, with regional averages obscuring pronounced differences in prevention effectiveness, maternal ART coverage, and continuity of antenatal and postnatal care across countries.

*These findings suggest that further reductions in MTCT will require geographically targeted strengthening of antenatal testing, maternal treatment initiation, and postnatal follow-up in persistently high-burden countries, rather than uniform regional strategies*.

### 3.7. Acceleration required to meet the 2030 target

Across outcomes, the pace of improvement observed from 2010 to 2024 remains insufficient to meet the 2030 benchmark of reducing each indicator to 10% of its 2010 level. Supplementary (Table S1) summarises (i) the observed annualised rate of change (ARC) from 2010 to 2024 and (ii) the required annualised rate (reqARC) from 2024 to 2030, computed from the pooled regional levels in 2010 and 2024.

For HIV incidence, the required declines are substantially steeper than recent trends in both regions, with the largest shortfalls observed among adolescents aged 15–19 years (e.g., ESA ARC −5.4%/year versus reqARC −22.5%/year; WCA ARC −6.6%/year versus reqARC −20.1%/year; Table **3**). The largest acceleration gaps occur for adolescent AIDS-related mortality and for MTCT in West and Central Africa, reinforcing that the remaining burden is sustained by a limited number of high-burden places and that achieving elimination targets will depend on geographically prioritised strengthening of adolescent- and maternal-focused services rather than uniform regional approaches.

### 3.8. Sensitivity analyses

Robustness was assessed using prespecified sensitivity analyses addressing panel composition, influential countries, and heterogeneity estimation. Balanced-panel re-estimation restricting to countries observed in both 2010 and 2024 yielded pooled estimates and annualised rates of change that closely matched the primary analysis (Supplementary Table S2). Leave-one-country-out refits demonstrated limited sensitivity to any single country, with no dominant influence on regional pooled estimates (Supplementary Table S3). Alternative estimators of between-country variance (*τ* ^2^), including Paule–Mandel and DerSimonian–Laird, produced near-identical pooled estimates and uncertainty intervals compared with REML (Supplementary Table S4).

Across all outcomes, 95% prediction intervals for 2024 were wide relative to confidence intervals, indicating substantial between-place dispersion beyond sampling uncertainty (Supplementary Table S2). Parametric bootstrap intervals for observed and required annualised rates of change (ARC and reqARC; *B* = 5,000) closely aligned with point estimates and confirmed that observed rates of decline remain insufficient to meet 2030 targets (Supplementary Table S5). Collectively, these findings confirm that observed spatial disparities reflect substantive between-place differences in epidemic burden and program performance, rather than artifacts of model specification or uncertainty propagation.

## 4. Discussion

This regional meta-analytic assessment of UNICEF-harmonized indicators for children and adolescents (0–19 years) across Eastern and Southern Africa (ESA) and West and Central Africa (WCA) documents substantial progress since 2010, yet persistent shortfalls relative to the Sustainable Development Goal (SDG) 3.3 target to end AIDS as a public health threat by 2030 [27, 1]. Importantly, our findings demonstrate that progress is not uniform across space: trajectories differ systematically by region, age, and gender, reflecting place-specific configurations of health infrastructure, service accessibility, gender norms, and historical investment patterns rather than epidemiological dynamics alone.

Consistent with global reporting, the steepest gains were observed among younger children, while adolescents, particularly girls aged 15–19, remain a critical equity gap [6]. Among children aged 0–14 years, AIDS-related mortality declined sharply between 2010 and 2024 (ESA: ∼80%; WCA: ∼74%), reflecting earlier and more consistent scale-up of pediatric antiretroviral therapy and PMTCT services. In contrast, mortality declines among adolescents were substantially slower (ESA: ∼36%; WCA: ∼46%), underscoring how the adolescent life course intersects with spatially uneven access to testing, linkage, and retention services [28, 29]. These regional averages conceal substantial between-place heterogeneity, consistent with geospatial syntheses showing that national or regional improvements can mask localized plateaus or reversals among adolescents [30, 31, 32]. The pronounced right-tailed distributions observed in ESA are consistent with the presence of a small number of countries with mature, generalized epidemics that continue to shape regional patterns, most notably South Africa. Despite major gains in treatment coverage and viral suppression, South Africa remains characterized by high background prevalence, age- and gender-assortative sexual networks, entrenched socioeconomic inequality, and syndemic risk environments that disproportionately affect adolescents and young adults [33, 34]. These place-specific dynamics amplify adolescent vulnerability even as national indicators improve, underscoring how aggregate progress can coexist with persistent localized risk.

Recent Global Burden of Disease (GBD) analyses corroborate this concentration of adolescent HIV burden, identifying South Africa as a dominant contributor to new infections among adolescents and young adults at both regional and global scales [35]. Updated GBD projections further suggest that, without accelerated and geographically targeted interventions, declines in adolescent incidence will lag behind those required to meet SDG 3.3, particularly in settings with large population size and entrenched structural risk [36]. Estimates from the Institute for Health Metrics and Evaluation (IHME) similarly highlight that regional averages obscure substantial within-region inequality, reinforcing the need to interpret adolescent HIV risk through a place-based lens rather than through national or regional means alone.

Sex-disaggregated analyses further highlight how place and gender intersect to shape risk. In 2024, adolescent girls experienced markedly higher HIV incidence than boys in both regions, with pooled female-to-male incidence rate ratios exceeding four in ESA and WCA. These disparities were robust across countries and not driven by a small number of outliers, indicating that gendered vulnerability is a region-wide spatial pattern rather than a localized anomaly. Such gaps are consistent with evidence that adolescent girls and young women (AGYW) face layered biological susceptibility, constrained prevention autonomy, and place-specific social norms governing sexuality, schooling, and mobility [37, 34, 9, 38]. The faster declines observed among boys, particularly in ESA, suggest that general improvements in health systems are insufficient without interventions explicitly tailored to the gendered and spatial realities shaping AGYW risk.

Mother-to-child transmission (MTCT) declined substantially across both regions, yet 2024 levels remain above WHO validation thresholds for elimination [39, 11]. Progress has been uneven across space: ESA’s earlier and more sustained investments in maternal ART and PMTCT delivery have translated into faster declines, whereas lower antenatal coverage, postpartum retention, and pediatric treatment access in WCA continue to slow progress [40, 41]. Postpartum and breastfeeding periods remain critical spatial–temporal windows of risk, particularly in settings where distance to facilities, supply interruptions, and workforce shortages undermine continuity of care [42, 43]. The required annualized declines in MTCT from 2024 to 2030 exceed recent experience in many WCA settings, highlighting the need for geographically targeted strengthening rather than uniform regional scale-up.

### Implications for policy and practice

A central implication of this analysis is that achieving SDG 3.3 will depend less on expanding generic coverage targets and more on aligning interventions with the spatial distribution of risk and capacity. The right-tailed country distributions observed for adolescent incidence indicate that a limited number of places account for a disproportionate share of new infections. In these settings, accelerating progress will require intensified, place-responsive strategies that integrate adolescent-friendly services within local health system constraints.

For countries requiring annual incidence reductions exceeding 20–25% among adolescents, feasible pathways include concentrating comprehensive AGYW prevention packages in high-incidence districts, leveraging school-linked and community-based platforms where facility access is limited, and expanding differentiated service delivery models that reduce visit burden. Where procurement and supply chains permit, long-acting cabotegravir (CAB-LA) offers an important additional prevention option, but its impact will depend on place-specific delivery models that account for clinic density, workforce availability, and adolescent mobility patterns [44, 45, 46]. Without such geographic tailoring, the promise of long-acting prevention risks reinforcing existing spatial inequities.

Closing remaining MTCT gaps, particularly in WCA, requires strengthening the full PMTCT cascade within local care geographies. Priorities include sustaining maternal viral suppression during postpartum and breast-feeding periods, improving early infant diagnosis coverage in remote areas, and ensuring reliable infant prophylaxis where health system reach is weakest [42, 43, 47, 39]. These goals are unlikely to be achieved through national averages alone and instead require district-level performance monitoring linked to corrective action.

Finally, accountability mechanisms should be explicitly spatial. Tracking observed annualized rates of change against required acceleration (reqARC) at subnational levels would allow policymakers to identify where progress is sufficient and where intensified support is urgently needed. Reporting prediction intervals and heterogeneity measures alongside pooled means can further shift focus from average performance to the realistic range of outcomes expected across places.

## 5. Conclusion

Substantial gains in pediatric HIV outcomes have been achieved across ESA and WCA, yet adolescents, especially girls, remain markedly off-track for 2030, and MTCT continues to exceed elimination thresholds in many settings. These gaps are not merely epidemiological but spatial, reflecting place-specific configurations of health systems, gender norms, and service accessibility. Converting SDG 3.3 from ambition to attainment will require sustained PMTCT scale-up focused on postpartum viral suppression, rapid expansion of gender-responsive adolescent prevention and care, and place-responsive monitoring of required acceleration. Embedding prevention and care strategies within the geographic realities shaping risk, access, and continuity of services is essential if progress toward SDG 3.3 is to be both accelerated and equitably realized.

## CRediT authorship contribution statement

**Chigozie Louisa J. Ugwu:** Conceptualization; Methodology; Formal analysis; Investigation; Validation; Visualization; Software; Writing – original draft; Writing – review & editing.

**Woldegebriel Assefa Woldegerima:** Conceptualization; Resources; Project administration; Supervision; Funding acquisition; Investigation; Writing – review & editing.

## Ethics approval statement

Ethics approval was not required. This study analyzed publicly available, aggregated data and did not involve human participants, animals, or identifiable information.

## Declaration of generative AI and AI-assisted technologies in the writing process

During the preparation of this manuscript, the authors used ChatGPT and Grammarly only to improve the clarity, grammar, and readability of the text. After applying these tools, the authors carefully reviewed, revised, and edited the content, and take full responsibility for the integrity and accuracy of the published article.

## Data availability

All data used in this study are publicly available from UNICEF/UNAIDS [16, 17, 18].

## Data Availability

All data used in this study are publicly available from UNICEF and UNAIDS through their online data portals. No new data were generated. Processed data and analytic code are available from the authors upon reasonable request.

