## Supplementary Table S1 for "Place, gender, and uneven progress in pediatric and adolescent HIV across sub-Saharan Africa: a regional meta-analytic assessment (2000–2024)"

Table 1: Observed and required annualised change to 2030 (target = 10% of the 2010 level) for HIV incidence, AIDS-related mortality, and mother-to-child transmission (MTCT). Pooled regional levels (2010 and 2024) are from random-effects meta-analysis (REML with Hartung–Knapp; Sex = both). ARC and reqARC are computed from these pooled levels.

| Region | Indicator | Age | 2010 | | | 2024 | | | Δ% | ARC | reqARC |
|---|---|---|---|---|---|---|---|---|---|---|---|
| | | | Level | [L, U] | $k$ | Level | [L, U] | $k$ | (10→24) | (%/yr) | (%/yr) |
| ESA | Incidence (per 1,000) | 0–14 | 1.784 | [0.894, 3.560] | 22 | 0.493 | [0.308, 0.787] | 21 | -72.4 | -8.8 | -15.6 |
| ESA | Incidence (per 1,000) | 15–19 | 1.035 | [0.388, 2.762] | 23 | 0.478 | [0.221, 1.032] | 22 | -53.9 | -5.4 | -22.5 |
| WCA | Incidence (per 1,000) | 0–14 | 0.986 | [0.632, 1.539] | 20 | 0.338 | [0.228, 0.502] | 20 | -65.7 | -7.4 | -18.6 |
| WCA | Incidence (per 1,000) | 15–19 | 0.484 | [0.257, 0.911] | 21 | 0.186 | [0.103, 0.339] | 21 | -61.4 | -6.6 | -20.1 |
| ESA | Mortality (per 100,000) | 0–14 | 52.926 | [26.795, 104.541] | 22 | 11.298 | [6.925, 18.433] | 21 | -78.7 | -10.4 | -11.9 |
| ESA | Mortality (per 100,000) | 15–19 | 15.032 | [6.892, 32.784] | 23 | 9.580 | [4.474, 20.512] | 22 | -36.3 | -3.2 | -26.6 |
| WCA | Mortality (per 100,000) | 0–14 | 33.259 | [22.903, 48.298] | 21 | 8.816 | [5.762, 13.488] | 21 | -73.5 | -9.0 | -15.0 |
| WCA | Mortality (per 100,000) | 15–19 | 10.053 | [6.641, 15.220] | 22 | 5.387 | [3.748, 7.744] | 22 | -46.4 | -4.4 | -24.4 |
| ESA | MTCT (%) | 0–4 | 25.976 | [20.275, 32.625] | 21 | 9.905 | [6.147, 15.580] | 21 | -61.9 | -6.7 | -20.0 |
| WCA | MTCT (%) | 0–4 | 29.559 | [26.272, 33.072] | 20 | 17.592 | [14.856, 20.709] | 20 | -40.5 | -3.6 | -25.7 |

*Notes:* ARC = annualised rate of change from 2010 to 2024; reqARC = required annualised change from 2024 to 2030 to reach 10% of the 2010 level. Negative values indicate declines. A balanced-panel acceleration analysis (restricting to countries observed in both 2010 and 2024 within each stratum) should be reported as a sensitivity table if emphasised in the Methods.
