## Supplementary Table S2 for "Place, gender, and uneven progress in pediatric and adolescent HIV across sub-Saharan Africa: a regional meta-analytic assessment (2000–2024)"

Table 1: Balanced panel (countries with both 2010 and 2024): pooled regional levels with uncertainty and pacing. Random-effects meta-analysis (REML) with Hartung–Knapp; prediction intervals (PI) shown for 2024.

| Indicator | Region | Age | $k_{2010}$ | $\tau^2_{2010}$ | $I^2_{2010}$ (%) | 2010 [95% CI] | $k_{2024}$ | $\tau^2_{2024}$ | $I^2_{2024}$ (%) | 2024 [95% CI] | 2024 PI |
|---|---|---|---|---|---|---|---|---|---|---|---|
| Estimated incidence rate (new HIV infection per 1,000 uninfected population) | ESA | 0–14 | 21 | 2.116 | 99.4 | 2.040 [1.048, 3.972] | 21 | 0.980 | 97.2 | 0.493 [0.308, 0.787] | 0.493 [0.059, 4.095] |
| Estimated incidence rate (new HIV infection per 1,000 uninfected population) | ESA | 15–19 | 22 | 4.390 | 95.4 | 1.244 [0.480, 3.222] | 22 | 2.804 | 93.5 | 0.478 [0.221, 1.032] | 0.478 [0.013, 16.905] |
| Estimated incidence rate (new HIV infection per 1,000 uninfected population) | WCA | 0–14 | 20 | 0.882 | 98.8 | 0.986 [0.632, 1.539] | 20 | 0.656 | 95.1 | 0.338 [0.228, 0.502] | 0.338 [0.059, 1.928] |
| Estimated incidence rate (new HIV infection per 1,000 uninfected population) | WCA | 15–19 | 21 | 1.727 | 91.6 | 0.484 [0.257, 0.911] | 21 | 1.525 | 89.0 | 0.186 [0.103, 0.339] | 0.186 [0.013, 2.623] |
| Estimated mother-to-child transmission rate (%) | ESA | 0–4 | 20 | 0.485 | 99.5 | 25.284 [19.559, 32.019] | 20 | 1.306 | 99.0 | 9.584 [5.818, 15.391] | 9.584 [0.905, 55.170] |
| Estimated mother-to-child transmission rate (%) | WCA | 0–4 | 20 | 0.114 | 98.0 | 29.559 [26.272, 33.072] | 20 | 0.159 | 92.1 | 17.592 [14.856, 20.709] | 17.592 [8.294, 33.504] |
| Estimated rate of annual AIDS-related deaths (per 100,000 population) | ESA | 0–14 | 21 | 2.054 | 99.6 | 60.435 [31.379, 116.395] | 21 | 1.076 | 96.2 | 11.298 [6.925, 18.433] | 11.298 [1.229, 103.847 |
| Estimated rate of annual AIDS-related deaths (per 100,000 population) | ESA | 15–19 | 22 | 3.156 | 99.7 | 16.433 [7.421, 36.391] | 22 | 2.806 | 99.5 | 9.580 [4.474, 20.512] | 9.580 [0.271, 338.878] |
| Estimated rate of annual AIDS-related deaths (per 100,000 population) | WCA | 0–14 | 21 | 0.664 | 99.0 | 33.259 [22.903, 48.298] | 21 | 0.825 | 95.9 | 8.816 [5.762, 13.488] | 8.816 [1.265, 61.438] |
| Estimated rate of annual AIDS-related deaths (per 100,000 population) | WCA | 15–19 | 22 | 0.861 | 98.7 | 10.053 [6.641, 15.220] | 22 | 0.642 | 96.1 | 5.387 [3.748, 7.744] | 5.387 [0.979, 29.642] |

*Notes:* $k$ = number of contributing countries; larger $\tau^2$ indicates greater between-country dispersion. ARC computed as $(y_{2024}y_{2010})^{1/14} - 1$.
