## Supplementary Table S3 for "Place, gender, and uneven progress in pediatric and adolescent HIV across sub-Saharan Africa: a regional meta-analytic assessment (2000–2024)"

Table 1: Leave-one-country-out influence analysis. Pooled value with each country left out in turn; range shown relative to base pooled value.

| Indicator | Region | Age | Year | Base pooled | Min (drop–1) | Max (drop–1) | Range (% of base) |
|---|---|---|---|---|---|---|---|
| Estimated incidence rate (new HIV infection per 1,000 uninfected population) | ESA | 0–14 | 2010 | 1.784 | 1.637 | 2.070 | 24.3 |
| Estimated incidence rate (new HIV infection per 1,000 uninfected population) | ESA | 0–14 | 2024 | 0.493 | 0.452 | 0.547 | 19.4 |
| Estimated incidence rate (new HIV infection per 1,000 uninfected population) | ESA | 15–19 | 2010 | 1.035 | 0.917 | 1.333 | 40.2 |
| Estimated incidence rate (new HIV infection per 1,000 uninfected population) | ESA | 15–19 | 2024 | 0.478 | 0.421 | 0.604 | 38.1 |
| Estimated incidence rate (new HIV infection per 1,000 uninfected population) | WCA | 0–14 | 2010 | 0.986 | 0.928 | 1.103 | 17.7 |
| Estimated incidence rate (new HIV infection per 1,000 uninfected population) | WCA | 0–14 | 2024 | 0.338 | 0.308 | 0.367 | 17.4 |
| Estimated incidence rate (new HIV infection per 1,000 uninfected population) | WCA | 15–19 | 2010 | 0.484 | 0.449 | 0.564 | 23.7 |
| Estimated incidence rate (new HIV infection per 1,000 uninfected population) | WCA | 15–19 | 2024 | 0.186 | 0.165 | 0.216 | 27.0 |
| Estimated mother-to-child transmission rate (%) | ESA | 0–4 | 2010 | 0.351 | 0.330 | 0.376 | 13.1 |
| Estimated mother-to-child transmission rate (%) | ESA | 0–4 | 2024 | 0.110 | 0.099 | 0.122 | 21.4 |
| Estimated mother-to-child transmission rate (%) | WCA | 0–4 | 2010 | 0.420 | 0.408 | 0.437 | 7.1 |
| Estimated mother-to-child transmission rate (%) | WCA | 0–4 | 2024 | 0.213 | 0.205 | 0.222 | 7.8 |
| Estimated rate of annual AIDS-related deaths (per 100,000 population) | ESA | 0–14 | 2010 | 52.926 | 48.264 | 61.857 | 25.7 |
| Estimated rate of annual AIDS-related deaths (per 100,000 population) | ESA | 0–14 | 2024 | 11.298 | 10.346 | 12.661 | 20.5 |
| Estimated rate of annual AIDS-related deaths (per 100,000 population) | ESA | 15–19 | 2010 | 15.032 | 13.663 | 18.390 | 31.4 |
| Estimated rate of annual AIDS-related deaths (per 100,000 population) | ESA | 15–19 | 2024 | 9.580 | 8.736 | 11.565 | 29.5 |
| Estimated rate of annual AIDS-related deaths (per 100,000 population) | WCA | 0–14 | 2010 | 33.259 | 31.542 | 35.869 | 13.0 |
| Estimated rate of annual AIDS-related deaths (per 100,000 population) | WCA | 0–14 | 2024 | 8.816 | 8.039 | 9.573 | 17.4 |
| Estimated rate of annual AIDS-related deaths (per 100,000 population) | WCA | 15–19 | 2010 | 10.053 | 9.403 | 11.045 | 16.3 |
| Estimated rate of annual AIDS-related deaths (per 100,000 popu- | WCA | 15–19 | 2024 | 5.387 | 5.085 | 5.793 | 13.1 |
