## Supplementary Table S4 for "Place, gender, and uneven progress in pediatric and adolescent HIV across sub-Saharan Africa: a regional meta-analytic assessment (2000–2024)"

Table 1: Alternative random-effects estimators: REML, Paule–Mandel (PM), and DerSimonian–Laird (DL); Hartung–Knapp confidence intervals.

| Indicator | Region | Age group | Year | Method | $k$ | $\tau^2$ | $I^2$ (%) | Pooled estimate [95% |
|---|---|---|---|---|---|---|---|---|
| Estimated incidence rate (new HIV infection per 1,000 uninfected population) | ESA | Age 0-14 | 2000 | REML | 22 | 3.245 | 99.8 | 4.045 [1.810, 9.040] |
| Estimated incidence rate (new HIV infection per 1,000 uninfected population) | ESA | Age 0-14 | 2000 | PM | 22 | 3.267 | 99.8 | 4.045 [1.810, 9.040] |
| Estimated incidence rate (new HIV infection per 1,000 uninfected population) | ESA | Age 0-14 | 2000 | DL | 22 | 1.064 | 99.4 | 4.127 [1.857, 9.174] |
| Estimated incidence rate (new HIV infection per 1,000 uninfected population) | ESA | Age 0-14 | 2001 | REML | 22 | 3.286 | 99.8 | 3.920 [1.749, 8.788] |
| Estimated incidence rate (new HIV infection per 1,000 uninfected population) | ESA | Age 0-14 | 2001 | PM | 22 | 3.296 | 99.8 | 3.920 [1.749, 8.788] |
| Estimated incidence rate (new HIV infection per 1,000 uninfected population) | ESA | Age 0-14 | 2001 | DL | 22 | 1.244 | 99.5 | 3.963 [1.771, 8.868] |
| Estimated incidence rate (new HIV infection per 1,000 uninfected population) | ESA | Age 0-14 | 2002 | REML | 22 | 3.202 | 99.8 | 3.807 [1.715, 8.451] |
| Estimated incidence rate (new HIV infection per 1,000 uninfected population) | ESA | Age 0-14 | 2002 | PM | 22 | 3.214 | 99.8 | 3.807 [1.715, 8.451] |
| Estimated incidence rate (new HIV infection per 1,000 uninfected population) | ESA | Age 0-14 | 2002 | DL | 22 | 1.221 | 99.5 | 3.856 [1.741, 8.538] |
| Estimated incidence rate (new HIV infection per 1,000 uninfected population) | ESA | Age 0-14 | 2003 | REML | 22 | 3.084 | 99.8 | 3.684 [1.686, 8.049] |
| Estimated incidence rate (new HIV infection per 1,000 uninfected population) | ESA | Age 0-14 | 2003 | PM | 22 | 3.089 | 99.8 | 3.684 [1.686, 8.049] |
| Estimated incidence rate (new HIV infection per 1,000 uninfected population) | ESA | Age 0-14 | 2003 | DL | 22 | 1.372 | 99.6 | 3.712 [1.700, 8.103] |
| Estimated incidence rate (new HIV infection per 1,000 uninfected population) | ESA | Age 0-14 | 2004 | REML | 22 | 3.025 | 99.8 | 3.488 [1.609, 7.562] |
| Estimated incidence rate (new HIV infection per 1,000 uninfected population) | ESA | Age 0-14 | 2004 | PM | 22 | 3.028 | 99.8 | 3.488 [1.609, 7.562] |
| Estimated incidence rate (new HIV infection per 1,000 uninfected population) | ESA | Age 0-14 | 2004 | DL | 22 | 1.484 | 99.6 | 3.509 [1.619, 7.604] |
| Estimated incidence rate (new HIV infection per 1,000 uninfected population) | ESA | Age 0-14 | 2005 | REML | 22 | 2.922 | 99.8 | 3.147 [1.471, 6.732] |
| Estimated incidence rate (new HIV infection per 1,000 uninfected population) | ESA | Age 0-14 | 2005 | PM | 22 | 2.923 | 99.8 | 3.147 [1.471, 6.732] |
| Estimated incidence rate (new HIV infection per 1,000 uninfected population) | ESA | Age 0-14 | 2005 | DL | 22 | 1.572 | 99.6 | 3.161 [1.478, 6.763] |
| Estimated incidence rate (new HIV infection per 1,000 uninfected population) | ESA | Age 0-14 | 2006 | REML | 22 | 2.789 | 99.7 | 2.835 [1.349, 5.957] |
| Estimated incidence rate (new HIV infection per 1,000 uninfected population) | ESA | Age 0-14 | 2006 | PM | 22 | 2.786 | 99.7 | 2.835 [1.349, 5.957] |
| Estimated incidence rate (new HIV infection per 1,000 uninfected population) | ESA | Age 0-14 | 2006 | DL | 22 | 1.809 | 99.6 | 2.841 [1.352, 5.971] |
| Estimated incidence rate (new HIV infection per 1,000 uninfected population) | ESA | Age 0-14 | 2007 | REML | 22 | 2.741 | 99.7 | 2.530 [1.211, 5.285] |
| Estimated incidence rate (new HIV infection per 1,000 uninfected population) | ESA | Age 0-14 | 2007 | PM | 22 | 2.738 | 99.7 | 2.530 [1.211, 5.285] |
| Estimated incidence rate (new HIV infection per 1,000 uninfected population) | ESA | Age 0-14 | 2007 | DL | 22 | 2.113 | 99.6 | 2.534 [1.213, 5.293] |
| Estimated incidence rate (new HIV infection per 1,000 uninfected population) | ESA | Age 0-14 | 2008 | REML | 22 | 2.697 | 99.6 | 2.277 [1.096, 4.729] |
| Estimated incidence rate (new HIV infection per 1,000 uninfected population) | ESA | Age 0-14 | 2008 | PM | 22 | 2.694 | 99.6 | 2.277 [1.096, 4.729] |
| Estimated incidence rate (new HIV infection per 1,000 uninfected population) | ESA | Age 0-14 | 2008 | DL | 22 | 2.116 | 99.5 | 2.280 [1.097, 4.735] |

*Continued on next p*

**Supplementary Table S4 (continued)**

| Indicator | Region | Age group | Year | Method | $k$ | $\tau^2$ | $I^2$ (%) | Pooled estimate [95% |
|---|---|---|---|---|---|---|---|---|
| Estimated incidence rate (new HIV infection per 1,000 uninfected population) | ESA | Age 0-14 | 2009 | REML | 22 | 2.534 | 99.5 | 1.996 [0.983, 4.054] |
| Estimated incidence rate (new HIV infection per 1,000 uninfected population) | ESA | Age 0-14 | 2009 | PM | 22 | 2.530 | 99.5 | 1.996 [0.983, 4.054] |
| Estimated incidence rate (new HIV infection per 1,000 uninfected population) | ESA | Age 0-14 | 2009 | DL | 22 | 2.077 | 99.4 | 1.998 [0.983, 4.058] |
| Estimated incidence rate (new HIV infection per 1,000 uninfected population) | ESA | Age 0-14 | 2010 | REML | 22 | 2.407 | 99.5 | 1.784 [0.894, 3.560] |
| Estimated incidence rate (new HIV infection per 1,000 uninfected population) | ESA | Age 0-14 | 2010 | PM | 22 | 2.405 | 99.5 | 1.784 [0.894, 3.560] |
| Estimated incidence rate (new HIV infection per 1,000 uninfected population) | ESA | Age 0-14 | 2010 | DL | 22 | 2.105 | 99.5 | 1.785 [0.894, 3.562] |
| Estimated incidence rate (new HIV infection per 1,000 uninfected population) | ESA | Age 0-14 | 2011 | REML | 22 | 2.272 | 99.5 | 1.495 [0.764, 2.927] |
| Estimated incidence rate (new HIV infection per 1,000 uninfected population) | ESA | Age 0-14 | 2011 | PM | 22 | 2.271 | 99.5 | 1.495 [0.764, 2.927] |
| Estimated incidence rate (new HIV infection per 1,000 uninfected population) | ESA | Age 0-14 | 2011 | DL | 22 | 2.230 | 99.5 | 1.495 [0.764, 2.927] |
| Estimated incidence rate (new HIV infection per 1,000 uninfected population) | ESA | Age 0-14 | 2012 | REML | 22 | 2.211 | 99.5 | 1.293 [0.666, 2.512] |
| Estimated incidence rate (new HIV infection per 1,000 uninfected population) | ESA | Age 0-14 | 2012 | PM | 22 | 2.213 | 99.5 | 1.293 [0.666, 2.512] |
| Estimated incidence rate (new HIV infection per 1,000 uninfected population) | ESA | Age 0-14 | 2012 | DL | 22 | 1.858 | 99.4 | 1.295 [0.667, 2.515] |
| Estimated incidence rate (new HIV infection per 1,000 uninfected population) | ESA | Age 0-14 | 2013 | REML | 22 | 2.208 | 99.4 | 1.189 [0.612, 2.308] |
| Estimated incidence rate (new HIV infection per 1,000 uninfected population) | ESA | Age 0-14 | 2013 | PM | 22 | 2.211 | 99.5 | 1.189 [0.612, 2.308] |
| Estimated incidence rate (new HIV infection per 1,000 uninfected population) | ESA | Age 0-14 | 2013 | DL | 22 | 1.726 | 99.3 | 1.191 [0.613, 2.312] |
| Estimated incidence rate (new HIV infection per 1,000 uninfected population) | ESA | Age 0-14 | 2014 | REML | 22 | 2.182 | 99.4 | 1.098 [0.567, 2.128] |
| Estimated incidence rate (new HIV infection per 1,000 uninfected population) | ESA | Age 0-14 | 2014 | PM | 22 | 2.191 | 99.4 | 1.098 [0.567, 2.128] |
| Estimated incidence rate (new HIV infection per 1,000 uninfected population) | ESA | Age 0-14 | 2014 | DL | 22 | 1.595 | 99.2 | 1.102 [0.569, 2.134] |
| Estimated incidence rate (new HIV infection per 1,000 uninfected population) | ESA | Age 0-14 | 2015 | REML | 22 | 2.130 | 99.4 | 0.949 [0.494, 1.821] |
| Estimated incidence rate (new HIV infection per 1,000 uninfected population) | ESA | Age 0-14 | 2015 | PM | 22 | 2.131 | 99.4 | 0.949 [0.494, 1.821] |
| Estimated incidence rate (new HIV infection per 1,000 uninfected population) | ESA | Age 0-14 | 2015 | DL | 22 | 2.129 | 99.4 | 0.949 [0.494, 1.821] |
| Estimated incidence rate (new HIV infection per 1,000 uninfected population) | ESA | Age 0-14 | 2016 | REML | 22 | 2.072 | 99.3 | 0.879 [0.461, 1.676] |
| Estimated incidence rate (new HIV infection per 1,000 uninfected population) | ESA | Age 0-14 | 2016 | PM | 22 | 2.079 | 99.3 | 0.879 [0.461, 1.676] |
| Estimated incidence rate (new HIV infection per 1,000 uninfected population) | ESA | Age 0-14 | 2016 | DL | 22 | 1.879 | 99.2 | 0.880 [0.462, 1.677] |
| Estimated incidence rate (new HIV infection per 1,000 uninfected population) | ESA | Age 0-14 | 2017 | REML | 22 | 1.933 | 99.3 | 0.793 [0.425, 1.479] |
| Estimated incidence rate (new HIV infection per 1,000 uninfected population) | ESA | Age 0-14 | 2017 | PM | 22 | 1.940 | 99.3 | 0.793 [0.425, 1.479] |
| Estimated incidence rate (new HIV infection per 1,000 uninfected population) | ESA | Age 0-14 | 2017 | DL | 22 | 2.918 | 99.5 | 0.790 [0.423, 1.475] |
| Estimated incidence rate (new HIV infection per 1,000 uninfected population) | ESA | Age 0-14 | 2018 | REML | 22 | 1.748 | 99.1 | 0.662 [0.365, 1.199] |
| Estimated incidence rate (new HIV infection per 1,000 uninfected population) | ESA | Age 0-14 | 2018 | PM | 22 | 1.757 | 99.1 | 0.662 [0.365, 1.199] |
| Estimated incidence rate (new HIV infection per 1,000 uninfected population) | ESA | Age 0-14 | 2018 | DL | 22 | 1.474 | 98.9 | 0.663 [0.366, 1.201] |

*Continued on next p*

**Supplementary Table S4 (continued)**

| Indicator | Region | Age group | Year | Method | $k$ | $\tau^2$ | $I^2$ (%) | Pooled estimate [95% |
|---|---|---|---|---|---|---|---|---|
| Estimated incidence rate (new HIV infection per 1,000 uninfected population) | ESA | Age 0-14 | 2019 | REML | 22 | 1.708 | 99.2 | 0.596 [0.332, 1.070] |
| Estimated incidence rate (new HIV infection per 1,000 uninfected population) | ESA | Age 0-14 | 2019 | PM | 22 | 1.709 | 99.2 | 0.596 [0.332, 1.070] |
| Estimated incidence rate (new HIV infection per 1,000 uninfected population) | ESA | Age 0-14 | 2019 | DL | 22 | 2.127 | 99.4 | 0.595 [0.331, 1.069] |
| Estimated incidence rate (new HIV infection per 1,000 uninfected population) | ESA | Age 0-14 | 2020 | REML | 22 | 1.618 | 99.1 | 0.545 [0.307, 0.967] |
| Estimated incidence rate (new HIV infection per 1,000 uninfected population) | ESA | Age 0-14 | 2020 | PM | 22 | 1.631 | 99.1 | 0.545 [0.307, 0.967] |
| Estimated incidence rate (new HIV infection per 1,000 uninfected population) | ESA | Age 0-14 | 2020 | DL | 22 | 0.989 | 98.5 | 0.549 [0.310, 0.972] |
| Estimated incidence rate (new HIV infection per 1,000 uninfected population) | ESA | Age 0-14 | 2021 | REML | 22 | 1.536 | 98.7 | 0.547 [0.313, 0.955] |
| Estimated incidence rate (new HIV infection per 1,000 uninfected population) | ESA | Age 0-14 | 2021 | PM | 22 | 1.541 | 98.7 | 0.547 [0.313, 0.955] |
| Estimated incidence rate (new HIV infection per 1,000 uninfected population) | ESA | Age 0-14 | 2021 | DL | 22 | 1.239 | 98.4 | 0.548 [0.314, 0.956] |
| Estimated incidence rate (new HIV infection per 1,000 uninfected population) | ESA | Age 0-14 | 2022 | REML | 22 | 1.593 | 98.2 | 0.530 [0.301, 0.935] |
| Estimated incidence rate (new HIV infection per 1,000 uninfected population) | ESA | Age 0-14 | 2022 | PM | 22 | 1.594 | 98.2 | 0.530 [0.301, 0.935] |
| Estimated incidence rate (new HIV infection per 1,000 uninfected population) | ESA | Age 0-14 | 2022 | DL | 22 | 1.999 | 98.6 | 0.530 [0.300, 0.934] |
| Estimated incidence rate (new HIV infection per 1,000 uninfected population) | ESA | Age 0-14 | 2023 | REML | 21 | 1.150 | 97.2 | 0.567 [0.343, 0.938] |
| Estimated incidence rate (new HIV infection per 1,000 uninfected population) | ESA | Age 0-14 | 2023 | PM | 21 | 1.174 | 97.2 | 0.567 [0.343, 0.938] |
| Estimated incidence rate (new HIV infection per 1,000 uninfected population) | ESA | Age 0-14 | 2023 | DL | 21 | 0.847 | 96.2 | 0.570 [0.345, 0.942] |
| Estimated incidence rate (new HIV infection per 1,000 uninfected population) | ESA | Age 0-14 | 2024 | REML | 21 | 0.980 | 97.2 | 0.493 [0.308, 0.787] |
| Estimated incidence rate (new HIV infection per 1,000 uninfected population) | ESA | Age 0-14 | 2024 | PM | 21 | 1.009 | 97.3 | 0.493 [0.308, 0.787] |
| Estimated incidence rate (new HIV infection per 1,000 uninfected population) | ESA | Age 0-14 | 2024 | DL | 21 | 0.773 | 96.5 | 0.496 [0.311, 0.791] |
| Estimated incidence rate (new HIV infection per 1,000 uninfected population) | ESA | Age 15-19 | 2000 | REML | 23 | 5.764 | 96.6 | 1.930 [0.672, 5.541] |
| Estimated incidence rate (new HIV infection per 1,000 uninfected population) | ESA | Age 15-19 | 2000 | PM | 23 | 5.654 | 96.5 | 1.929 [0.672, 5.540] |
| Estimated incidence rate (new HIV infection per 1,000 uninfected population) | ESA | Age 15-19 | 2000 | DL | 23 | 11.289 | 98.2 | 1.952 [0.683, 5.579] |
| Estimated incidence rate (new HIV infection per 1,000 uninfected population) | ESA | Age 15-19 | 2001 | REML | 23 | 5.555 | 96.6 | 1.827 [0.647, 5.159] |
| Estimated incidence rate (new HIV infection per 1,000 uninfected population) | ESA | Age 15-19 | 2001 | PM | 23 | 5.457 | 96.6 | 1.826 [0.647, 5.158] |
| Estimated incidence rate (new HIV infection per 1,000 uninfected population) | ESA | Age 15-19 | 2001 | DL | 23 | 10.793 | 98.2 | 1.843 [0.655, 5.184] |
| Estimated incidence rate (new HIV infection per 1,000 uninfected population) | ESA | Age 15-19 | 2002 | REML | 23 | 5.510 | 96.5 | 1.685 [0.600, 4.732] |
| Estimated incidence rate (new HIV infection per 1,000 uninfected population) | ESA | Age 15-19 | 2002 | PM | 23 | 5.405 | 96.5 | 1.684 [0.599, 4.731] |
| Estimated incidence rate (new HIV infection per 1,000 uninfected population) | ESA | Age 15-19 | 2002 | DL | 23 | 10.635 | 98.2 | 1.703 [0.609, 4.763] |
| Estimated incidence rate (new HIV infection per 1,000 uninfected population) | ESA | Age 15-19 | 2003 | REML | 23 | 5.328 | 96.3 | 1.604 [0.580, 4.433] |
| Estimated incidence rate (new HIV infection per 1,000 uninfected population) | ESA | Age 15-19 | 2003 | PM | 23 | 5.227 | 96.3 | 1.603 [0.580, 4.431] |
| Estimated incidence rate (new HIV infection per 1,000 uninfected population) | ESA | Age 15-19 | 2003 | DL | 23 | 10.475 | 98.1 | 1.621 [0.589, 4.461] |

*Continued on next p*

**Supplementary Table S4 (continued)**

| Indicator | Region | Age group | Year | Method | $k$ | $\tau^2$ | $I^2$ (%) | Pooled estimate [95% |
|---|---|---|---|---|---|---|---|---|
| Estimated incidence rate (new HIV infection per 1,000 uninfected population) | ESA | Age 15-19 | 2004 | REML | 23 | 5.332 | 96.8 | 1.499 [0.543, 4.141] |
| Estimated incidence rate (new HIV infection per 1,000 uninfected population) | ESA | Age 15-19 | 2004 | PM | 23 | 5.231 | 96.7 | 1.498 [0.542, 4.140] |
| Estimated incidence rate (new HIV infection per 1,000 uninfected population) | ESA | Age 15-19 | 2004 | DL | 23 | 9.541 | 98.2 | 1.514 [0.550, 4.166] |
| Estimated incidence rate (new HIV infection per 1,000 uninfected population) | ESA | Age 15-19 | 2005 | REML | 23 | 5.263 | 96.6 | 1.402 [0.510, 3.851] |
| Estimated incidence rate (new HIV infection per 1,000 uninfected population) | ESA | Age 15-19 | 2005 | PM | 23 | 5.159 | 96.6 | 1.401 [0.510, 3.850] |
| Estimated incidence rate (new HIV infection per 1,000 uninfected population) | ESA | Age 15-19 | 2005 | DL | 23 | 9.540 | 98.1 | 1.416 [0.518, 3.875] |
| Estimated incidence rate (new HIV infection per 1,000 uninfected population) | ESA | Age 15-19 | 2006 | REML | 23 | 5.133 | 96.5 | 1.337 [0.492, 3.629] |
| Estimated incidence rate (new HIV infection per 1,000 uninfected population) | ESA | Age 15-19 | 2006 | PM | 23 | 5.030 | 96.5 | 1.336 [0.492, 3.628] |
| Estimated incidence rate (new HIV infection per 1,000 uninfected population) | ESA | Age 15-19 | 2006 | DL | 23 | 9.258 | 98.0 | 1.351 [0.500, 3.654] |
| Estimated incidence rate (new HIV infection per 1,000 uninfected population) | ESA | Age 15-19 | 2007 | REML | 23 | 5.205 | 96.0 | 1.238 [0.453, 3.386] |
| Estimated incidence rate (new HIV infection per 1,000 uninfected population) | ESA | Age 15-19 | 2007 | PM | 23 | 5.107 | 96.0 | 1.237 [0.452, 3.385] |
| Estimated incidence rate (new HIV infection per 1,000 uninfected population) | ESA | Age 15-19 | 2007 | DL | 23 | 10.138 | 97.9 | 1.251 [0.459, 3.409] |
| Estimated incidence rate (new HIV infection per 1,000 uninfected population) | ESA | Age 15-19 | 2008 | REML | 23 | 5.197 | 96.1 | 1.164 [0.426, 3.181] |
| Estimated incidence rate (new HIV infection per 1,000 uninfected population) | ESA | Age 15-19 | 2008 | PM | 23 | 5.105 | 96.0 | 1.163 [0.426, 3.180] |
| Estimated incidence rate (new HIV infection per 1,000 uninfected population) | ESA | Age 15-19 | 2008 | DL | 23 | 9.943 | 97.9 | 1.176 [0.432, 3.203] |
| Estimated incidence rate (new HIV infection per 1,000 uninfected population) | ESA | Age 15-19 | 2009 | REML | 23 | 5.059 | 96.2 | 1.091 [0.404, 2.943] |
| Estimated incidence rate (new HIV infection per 1,000 uninfected population) | ESA | Age 15-19 | 2009 | PM | 23 | 4.970 | 96.2 | 1.090 [0.404, 2.942] |
| Estimated incidence rate (new HIV infection per 1,000 uninfected population) | ESA | Age 15-19 | 2009 | DL | 23 | 9.053 | 97.8 | 1.102 [0.410, 2.964] |
| Estimated incidence rate (new HIV infection per 1,000 uninfected population) | ESA | Age 15-19 | 2010 | REML | 23 | 4.938 | 96.0 | 1.035 [0.388, 2.762] |
| Estimated incidence rate (new HIV infection per 1,000 uninfected population) | ESA | Age 15-19 | 2010 | PM | 23 | 4.855 | 96.0 | 1.035 [0.388, 2.761] |
| Estimated incidence rate (new HIV infection per 1,000 uninfected population) | ESA | Age 15-19 | 2010 | DL | 23 | 9.033 | 97.8 | 1.045 [0.393, 2.779] |
| Estimated incidence rate (new HIV infection per 1,000 uninfected population) | ESA | Age 15-19 | 2011 | REML | 23 | 4.946 | 96.1 | 0.979 [0.367, 2.614] |
| Estimated incidence rate (new HIV infection per 1,000 uninfected population) | ESA | Age 15-19 | 2011 | PM | 23 | 4.867 | 96.0 | 0.978 [0.366, 2.613] |
| Estimated incidence rate (new HIV infection per 1,000 uninfected population) | ESA | Age 15-19 | 2011 | DL | 23 | 8.803 | 97.7 | 0.989 [0.371, 2.632] |
| Estimated incidence rate (new HIV infection per 1,000 uninfected population) | ESA | Age 15-19 | 2012 | REML | 23 | 5.136 | 96.2 | 0.905 [0.333, 2.459] |
| Estimated incidence rate (new HIV infection per 1,000 uninfected population) | ESA | Age 15-19 | 2012 | PM | 23 | 5.058 | 96.2 | 0.904 [0.333, 2.458] |
| Estimated incidence rate (new HIV infection per 1,000 uninfected population) | ESA | Age 15-19 | 2012 | DL | 23 | 8.850 | 97.8 | 0.913 [0.337, 2.476] |
| Estimated incidence rate (new HIV infection per 1,000 uninfected population) | ESA | Age 15-19 | 2013 | REML | 23 | 5.135 | 96.1 | 0.836 [0.308, 2.275] |
| Estimated incidence rate (new HIV infection per 1,000 uninfected population) | ESA | Age 15-19 | 2013 | PM | 23 | 5.060 | 96.0 | 0.836 [0.307, 2.274] |
| Estimated incidence rate (new HIV infection per 1,000 uninfected population) | ESA | Age 15-19 | 2013 | DL | 23 | 8.969 | 97.7 | 0.844 [0.311, 2.288] |

*Continued on next*

**Supplementary Table S4 (continued)**

| Indicator | Region | Age group | Year | Method | $k$ | $\tau^2$ | $I^2$ (%) | Pooled estimate [95% |
|---|---|---|---|---|---|---|---|---|
| Estimated incidence rate (new HIV infection per 1,000 uninfected population) | ESA | Age 15-19 | 2014 | REML | 23 | 4.954 | 96.0 | 0.769 [0.288, 2.057] |
| Estimated incidence rate (new HIV infection per 1,000 uninfected population) | ESA | Age 15-19 | 2014 | PM | 23 | 4.879 | 95.9 | 0.769 [0.288, 2.057] |
| Estimated incidence rate (new HIV infection per 1,000 uninfected population) | ESA | Age 15-19 | 2014 | DL | 23 | 8.530 | 97.6 | 0.777 [0.291, 2.073] |
| Estimated incidence rate (new HIV infection per 1,000 uninfected population) | ESA | Age 15-19 | 2015 | REML | 23 | 5.082 | 96.0 | 0.674 [0.249, 1.825] |
| Estimated incidence rate (new HIV infection per 1,000 uninfected population) | ESA | Age 15-19 | 2015 | PM | 23 | 5.004 | 96.0 | 0.674 [0.249, 1.825] |
| Estimated incidence rate (new HIV infection per 1,000 uninfected population) | ESA | Age 15-19 | 2015 | DL | 23 | 8.641 | 97.6 | 0.680 [0.252, 1.836] |
| Estimated incidence rate (new HIV infection per 1,000 uninfected population) | ESA | Age 15-19 | 2016 | REML | 22 | 4.050 | 95.0 | 0.758 [0.303, 1.896] |
| Estimated incidence rate (new HIV infection per 1,000 uninfected population) | ESA | Age 15-19 | 2016 | PM | 22 | 3.986 | 95.0 | 0.758 [0.303, 1.896] |
| Estimated incidence rate (new HIV infection per 1,000 uninfected population) | ESA | Age 15-19 | 2016 | DL | 22 | 8.031 | 97.4 | 0.765 [0.307, 1.907] |
| Estimated incidence rate (new HIV infection per 1,000 uninfected population) | ESA | Age 15-19 | 2017 | REML | 22 | 3.878 | 94.8 | 0.738 [0.301, 1.811] |
| Estimated incidence rate (new HIV infection per 1,000 uninfected population) | ESA | Age 15-19 | 2017 | PM | 22 | 3.812 | 94.8 | 0.738 [0.300, 1.811] |
| Estimated incidence rate (new HIV infection per 1,000 uninfected population) | ESA | Age 15-19 | 2017 | DL | 22 | 7.784 | 97.4 | 0.746 [0.305, 1.824] |
| Estimated incidence rate (new HIV infection per 1,000 uninfected population) | ESA | Age 15-19 | 2018 | REML | 23 | 4.745 | 96.6 | 0.562 [0.215, 1.470] |
| Estimated incidence rate (new HIV infection per 1,000 uninfected population) | ESA | Age 15-19 | 2018 | PM | 23 | 4.665 | 96.5 | 0.562 [0.215, 1.470] |
| Estimated incidence rate (new HIV infection per 1,000 uninfected population) | ESA | Age 15-19 | 2018 | DL | 23 | 8.121 | 98.0 | 0.569 [0.218, 1.482] |
| Estimated incidence rate (new HIV infection per 1,000 uninfected population) | ESA | Age 15-19 | 2019 | REML | 23 | 4.535 | 96.5 | 0.533 [0.208, 1.363] |
| Estimated incidence rate (new HIV infection per 1,000 uninfected population) | ESA | Age 15-19 | 2019 | PM | 23 | 4.457 | 96.4 | 0.532 [0.208, 1.363] |
| Estimated incidence rate (new HIV infection per 1,000 uninfected population) | ESA | Age 15-19 | 2019 | DL | 23 | 7.965 | 98.0 | 0.539 [0.211, 1.375] |
| Estimated incidence rate (new HIV infection per 1,000 uninfected population) | ESA | Age 15-19 | 2020 | REML | 23 | 4.378 | 96.3 | 0.486 [0.193, 1.225] |
| Estimated incidence rate (new HIV infection per 1,000 uninfected population) | ESA | Age 15-19 | 2020 | PM | 23 | 4.303 | 96.2 | 0.486 [0.193, 1.224] |
| Estimated incidence rate (new HIV infection per 1,000 uninfected population) | ESA | Age 15-19 | 2020 | DL | 23 | 7.776 | 97.9 | 0.492 [0.196, 1.235] |
| Estimated incidence rate (new HIV infection per 1,000 uninfected population) | ESA | Age 15-19 | 2021 | REML | 23 | 4.154 | 96.1 | 0.482 [0.196, 1.186] |
| Estimated incidence rate (new HIV infection per 1,000 uninfected population) | ESA | Age 15-19 | 2021 | PM | 23 | 4.073 | 96.0 | 0.482 [0.196, 1.185] |
| Estimated incidence rate (new HIV infection per 1,000 uninfected population) | ESA | Age 15-19 | 2021 | DL | 23 | 7.542 | 97.8 | 0.489 [0.200, 1.199] |
| Estimated incidence rate (new HIV infection per 1,000 uninfected population) | ESA | Age 15-19 | 2022 | REML | 23 | 3.953 | 95.9 | 0.433 [0.180, 1.042] |
| Estimated incidence rate (new HIV infection per 1,000 uninfected population) | ESA | Age 15-19 | 2022 | PM | 23 | 3.874 | 95.9 | 0.432 [0.179, 1.042] |
| Estimated incidence rate (new HIV infection per 1,000 uninfected population) | ESA | Age 15-19 | 2022 | DL | 23 | 7.310 | 97.8 | 0.438 [0.183, 1.052] |
| Estimated incidence rate (new HIV infection per 1,000 uninfected population) | ESA | Age 15-19 | 2023 | REML | 22 | 2.926 | 93.5 | 0.506 [0.230, 1.110] |
| Estimated incidence rate (new HIV infection per 1,000 uninfected population) | ESA | Age 15-19 | 2023 | PM | 22 | 2.871 | 93.4 | 0.505 [0.230, 1.110] |
| Estimated incidence rate (new HIV infection per 1,000 uninfected population) | ESA | Age 15-19 | 2023 | DL | 22 | 6.149 | 96.8 | 0.512 [0.234, 1.119] |

*Continued on next [illegible]*

**Supplementary Table S4 (continued)**

| Indicator | Region | Age group | Year | Method | $k$ | $\tau^2$ | $I^2$ (%) | Pooled estimate [95% |
|---|---|---|---|---|---|---|---|---|
| Estimated incidence rate (new HIV infection per 1,000 uninfected population) | ESA | Age 15-19 | 2024 | REML | 22 | 2.804 | 93.5 | 0.478 [0.221, 1.032] |
| Estimated incidence rate (new HIV infection per 1,000 uninfected population) | ESA | Age 15-19 | 2024 | PM | 22 | 2.752 | 93.4 | 0.477 [0.221, 1.032] |
| Estimated incidence rate (new HIV infection per 1,000 uninfected population) | ESA | Age 15-19 | 2024 | DL | 22 | 5.812 | 96.8 | 0.483 [0.224, 1.040] |
| Estimated incidence rate (new HIV infection per 1,000 uninfected population) | WCA | Age 0-14 | 2000 | REML | 20 | 0.585 | 98.5 | 2.028 [1.413, 2.910] |
| Estimated incidence rate (new HIV infection per 1,000 uninfected population) | WCA | Age 0-14 | 2000 | PM | 20 | 0.584 | 98.5 | 2.028 [1.413, 2.910] |
| Estimated incidence rate (new HIV infection per 1,000 uninfected population) | WCA | Age 0-14 | 2000 | DL | 20 | 0.742 | 98.8 | 2.028 [1.413, 2.910] |
| Estimated incidence rate (new HIV infection per 1,000 uninfected population) | WCA | Age 0-14 | 2001 | REML | 20 | 0.582 | 98.7 | 1.982 [1.383, 2.840] |
| Estimated incidence rate (new HIV infection per 1,000 uninfected population) | WCA | Age 0-14 | 2001 | PM | 20 | 0.581 | 98.7 | 1.982 [1.383, 2.840] |
| Estimated incidence rate (new HIV infection per 1,000 uninfected population) | WCA | Age 0-14 | 2001 | DL | 20 | 0.698 | 98.9 | 1.982 [1.383, 2.841] |
| Estimated incidence rate (new HIV infection per 1,000 uninfected population) | WCA | Age 0-14 | 2002 | REML | 20 | 0.581 | 98.6 | 1.922 [1.341, 2.753] |
| Estimated incidence rate (new HIV infection per 1,000 uninfected population) | WCA | Age 0-14 | 2002 | PM | 20 | 0.579 | 98.6 | 1.922 [1.341, 2.753] |
| Estimated incidence rate (new HIV infection per 1,000 uninfected population) | WCA | Age 0-14 | 2002 | DL | 20 | 0.721 | 98.9 | 1.922 [1.342, 2.754] |
| Estimated incidence rate (new HIV infection per 1,000 uninfected population) | WCA | Age 0-14 | 2003 | REML | 20 | 0.582 | 98.7 | 1.845 [1.288, 2.643] |
| Estimated incidence rate (new HIV infection per 1,000 uninfected population) | WCA | Age 0-14 | 2003 | PM | 20 | 0.580 | 98.7 | 1.845 [1.288, 2.643] |
| Estimated incidence rate (new HIV infection per 1,000 uninfected population) | WCA | Age 0-14 | 2003 | DL | 20 | 0.699 | 98.9 | 1.845 [1.288, 2.643] |
| Estimated incidence rate (new HIV infection per 1,000 uninfected population) | WCA | Age 0-14 | 2004 | REML | 20 | 0.582 | 98.6 | 1.756 [1.226, 2.515] |
| Estimated incidence rate (new HIV infection per 1,000 uninfected population) | WCA | Age 0-14 | 2004 | PM | 20 | 0.580 | 98.6 | 1.756 [1.226, 2.515] |
| Estimated incidence rate (new HIV infection per 1,000 uninfected population) | WCA | Age 0-14 | 2004 | DL | 20 | 0.714 | 98.9 | 1.756 [1.226, 2.516] |
| Estimated incidence rate (new HIV infection per 1,000 uninfected population) | WCA | Age 0-14 | 2005 | REML | 20 | 0.597 | 98.8 | 1.662 [1.155, 2.392] |
| Estimated incidence rate (new HIV infection per 1,000 uninfected population) | WCA | Age 0-14 | 2005 | PM | 20 | 0.596 | 98.8 | 1.662 [1.155, 2.392] |
| Estimated incidence rate (new HIV infection per 1,000 uninfected population) | WCA | Age 0-14 | 2005 | DL | 20 | 0.718 | 99.0 | 1.663 [1.155, 2.392] |
| Estimated incidence rate (new HIV infection per 1,000 uninfected population) | WCA | Age 0-14 | 2006 | REML | 20 | 0.609 | 98.8 | 1.553 [1.075, 2.242] |
| Estimated incidence rate (new HIV infection per 1,000 uninfected population) | WCA | Age 0-14 | 2006 | PM | 20 | 0.608 | 98.8 | 1.553 [1.075, 2.242] |
| Estimated incidence rate (new HIV infection per 1,000 uninfected population) | WCA | Age 0-14 | 2006 | DL | 20 | 0.757 | 99.1 | 1.553 [1.076, 2.243] |
| Estimated incidence rate (new HIV infection per 1,000 uninfected population) | WCA | Age 0-14 | 2007 | REML | 20 | 0.631 | 98.7 | 1.413 [0.972, 2.055] |
| Estimated incidence rate (new HIV infection per 1,000 uninfected population) | WCA | Age 0-14 | 2007 | PM | 20 | 0.629 | 98.7 | 1.413 [0.972, 2.055] |
| Estimated incidence rate (new HIV infection per 1,000 uninfected population) | WCA | Age 0-14 | 2007 | DL | 20 | 0.829 | 99.0 | 1.414 [0.973, 2.055] |
| Estimated incidence rate (new HIV infection per 1,000 uninfected population) | WCA | Age 0-14 | 2008 | REML | 20 | 0.624 | 98.5 | 1.271 [0.876, 1.844] |
| Estimated incidence rate (new HIV infection per 1,000 uninfected population) | WCA | Age 0-14 | 2008 | PM | 20 | 0.621 | 98.5 | 1.271 [0.876, 1.844] |
| Estimated incidence rate (new HIV infection per 1,000 uninfected population) | WCA | Age 0-14 | 2008 | DL | 20 | 0.884 | 98.9 | 1.272 [0.877, 1.845] |

*Continued on next p*

**Supplementary Table S4 (continued)**

| Indicator | Region | Age group | Year | Method | $k$ | $\tau^2$ | $I^2$ (%) | Pooled estimate [95% |
|---|---|---|---|---|---|---|---|---|
| Estimated incidence rate (new HIV infection per 1,000 uninfected population) | WCA | Age 0-14 | 2009 | REML | 20 | 0.680 | 98.5 | 1.173 [0.795, 1.730] |
| Estimated incidence rate (new HIV infection per 1,000 uninfected population) | WCA | Age 0-14 | 2009 | PM | 20 | 0.678 | 98.5 | 1.173 [0.795, 1.730] |
| Estimated incidence rate (new HIV infection per 1,000 uninfected population) | WCA | Age 0-14 | 2009 | DL | 20 | 0.910 | 98.9 | 1.173 [0.796, 1.730] |
| Estimated incidence rate (new HIV infection per 1,000 uninfected population) | WCA | Age 0-14 | 2010 | REML | 20 | 0.882 | 98.8 | 0.986 [0.632, 1.539] |
| Estimated incidence rate (new HIV infection per 1,000 uninfected population) | WCA | Age 0-14 | 2010 | PM | 20 | 0.887 | 98.8 | 0.986 [0.632, 1.539] |
| Estimated incidence rate (new HIV infection per 1,000 uninfected population) | WCA | Age 0-14 | 2010 | DL | 20 | 0.968 | 98.9 | 0.986 [0.632, 1.538] |
| Estimated incidence rate (new HIV infection per 1,000 uninfected population) | WCA | Age 0-14 | 2011 | REML | 20 | 0.778 | 98.3 | 0.868 [0.572, 1.316] |
| Estimated incidence rate (new HIV infection per 1,000 uninfected population) | WCA | Age 0-14 | 2011 | PM | 20 | 0.775 | 98.3 | 0.868 [0.572, 1.316] |
| Estimated incidence rate (new HIV infection per 1,000 uninfected population) | WCA | Age 0-14 | 2011 | DL | 20 | 0.909 | 98.5 | 0.868 [0.572, 1.316] |
| Estimated incidence rate (new HIV infection per 1,000 uninfected population) | WCA | Age 0-14 | 2012 | REML | 20 | 0.744 | 98.3 | 0.787 [0.524, 1.183] |
| Estimated incidence rate (new HIV infection per 1,000 uninfected population) | WCA | Age 0-14 | 2012 | PM | 20 | 0.739 | 98.3 | 0.787 [0.524, 1.183] |
| Estimated incidence rate (new HIV infection per 1,000 uninfected population) | WCA | Age 0-14 | 2012 | DL | 20 | 0.927 | 98.6 | 0.787 [0.524, 1.183] |
| Estimated incidence rate (new HIV infection per 1,000 uninfected population) | WCA | Age 0-14 | 2013 | REML | 20 | 0.756 | 98.1 | 0.697 [0.462, 1.051] |
| Estimated incidence rate (new HIV infection per 1,000 uninfected population) | WCA | Age 0-14 | 2013 | PM | 20 | 0.749 | 98.1 | 0.697 [0.462, 1.051] |
| Estimated incidence rate (new HIV infection per 1,000 uninfected population) | WCA | Age 0-14 | 2013 | DL | 20 | 1.034 | 98.6 | 0.698 [0.463, 1.051] |
| Estimated incidence rate (new HIV infection per 1,000 uninfected population) | WCA | Age 0-14 | 2014 | REML | 20 | 0.839 | 98.1 | 0.606 [0.392, 0.935] |
| Estimated incidence rate (new HIV infection per 1,000 uninfected population) | WCA | Age 0-14 | 2014 | PM | 20 | 0.837 | 98.1 | 0.606 [0.392, 0.935] |
| Estimated incidence rate (new HIV infection per 1,000 uninfected population) | WCA | Age 0-14 | 2014 | DL | 20 | 1.070 | 98.5 | 0.605 [0.392, 0.935] |
| Estimated incidence rate (new HIV infection per 1,000 uninfected population) | WCA | Age 0-14 | 2015 | REML | 20 | 0.832 | 98.2 | 0.572 [0.372, 0.882] |
| Estimated incidence rate (new HIV infection per 1,000 uninfected population) | WCA | Age 0-14 | 2015 | PM | 20 | 0.827 | 98.2 | 0.572 [0.372, 0.882] |
| Estimated incidence rate (new HIV infection per 1,000 uninfected population) | WCA | Age 0-14 | 2015 | DL | 20 | 1.159 | 98.7 | 0.573 [0.372, 0.882] |
| Estimated incidence rate (new HIV infection per 1,000 uninfected population) | WCA | Age 0-14 | 2016 | REML | 20 | 0.858 | 98.2 | 0.562 [0.363, 0.872] |
| Estimated incidence rate (new HIV infection per 1,000 uninfected population) | WCA | Age 0-14 | 2016 | PM | 20 | 0.855 | 98.2 | 0.562 [0.363, 0.872] |
| Estimated incidence rate (new HIV infection per 1,000 uninfected population) | WCA | Age 0-14 | 2016 | DL | 20 | 1.128 | 98.6 | 0.563 [0.363, 0.872] |
| Estimated incidence rate (new HIV infection per 1,000 uninfected population) | WCA | Age 0-14 | 2017 | REML | 20 | 0.800 | 98.1 | 0.553 [0.362, 0.844] |
| Estimated incidence rate (new HIV infection per 1,000 uninfected population) | WCA | Age 0-14 | 2017 | PM | 20 | 0.793 | 98.1 | 0.553 [0.362, 0.844] |
| Estimated incidence rate (new HIV infection per 1,000 uninfected population) | WCA | Age 0-14 | 2017 | DL | 20 | 1.272 | 98.8 | 0.554 [0.363, 0.845] |
| Estimated incidence rate (new HIV infection per 1,000 uninfected population) | WCA | Age 0-14 | 2018 | REML | 20 | 0.898 | 98.0 | 0.507 [0.324, 0.794] |
| Estimated incidence rate (new HIV infection per 1,000 uninfected population) | WCA | Age 0-14 | 2018 | PM | 20 | 0.891 | 98.0 | 0.507 [0.324, 0.794] |
| Estimated incidence rate (new HIV infection per 1,000 uninfected population) | WCA | Age 0-14 | 2018 | DL | 20 | 1.361 | 98.7 | 0.508 [0.325, 0.795] |

*Continued on next p*

**Supplementary Table S4 (continued)**

| Indicator | Region | Age group | Year | Method | $k$ | $\tau^2$ | $I^2$ (%) | Pooled estimate [95% |
|---|---|---|---|---|---|---|---|---|
| Estimated incidence rate (new HIV infection per 1,000 uninfected population) | WCA | Age 0-14 | 2019 | REML | 20 | 0.889 | 97.5 | 0.458 [0.293, 0.717] |
| Estimated incidence rate (new HIV infection per 1,000 uninfected population) | WCA | Age 0-14 | 2019 | PM | 20 | 0.880 | 97.4 | 0.458 [0.293, 0.717] |
| Estimated incidence rate (new HIV infection per 1,000 uninfected population) | WCA | Age 0-14 | 2019 | DL | 20 | 1.308 | 98.3 | 0.459 [0.294, 0.717] |
| Estimated incidence rate (new HIV infection per 1,000 uninfected population) | WCA | Age 0-14 | 2020 | REML | 20 | 0.806 | 96.9 | 0.453 [0.295, 0.694] |
| Estimated incidence rate (new HIV infection per 1,000 uninfected population) | WCA | Age 0-14 | 2020 | PM | 20 | 0.797 | 96.9 | 0.453 [0.295, 0.694] |
| Estimated incidence rate (new HIV infection per 1,000 uninfected population) | WCA | Age 0-14 | 2020 | DL | 20 | 1.261 | 98.0 | 0.454 [0.296, 0.695] |
| Estimated incidence rate (new HIV infection per 1,000 uninfected population) | WCA | Age 0-14 | 2021 | REML | 20 | 0.824 | 97.0 | 0.409 [0.265, 0.631] |
| Estimated incidence rate (new HIV infection per 1,000 uninfected population) | WCA | Age 0-14 | 2021 | PM | 20 | 0.818 | 96.9 | 0.409 [0.265, 0.631] |
| Estimated incidence rate (new HIV infection per 1,000 uninfected population) | WCA | Age 0-14 | 2021 | DL | 20 | 1.183 | 97.9 | 0.409 [0.265, 0.631] |
| Estimated incidence rate (new HIV infection per 1,000 uninfected population) | WCA | Age 0-14 | 2022 | REML | 20 | 0.812 | 97.5 | 0.400 [0.260, 0.616] |
| Estimated incidence rate (new HIV infection per 1,000 uninfected population) | WCA | Age 0-14 | 2022 | PM | 20 | 0.807 | 97.4 | 0.400 [0.260, 0.616] |
| Estimated incidence rate (new HIV infection per 1,000 uninfected population) | WCA | Age 0-14 | 2022 | DL | 20 | 1.368 | 98.5 | 0.401 [0.260, 0.617] |
| Estimated incidence rate (new HIV infection per 1,000 uninfected population) | WCA | Age 0-14 | 2023 | REML | 20 | 0.750 | 96.3 | 0.375 [0.247, 0.570] |
| Estimated incidence rate (new HIV infection per 1,000 uninfected population) | WCA | Age 0-14 | 2023 | PM | 20 | 0.744 | 96.2 | 0.375 [0.247, 0.570] |
| Estimated incidence rate (new HIV infection per 1,000 uninfected population) | WCA | Age 0-14 | 2023 | DL | 20 | 1.017 | 97.2 | 0.376 [0.248, 0.571] |
| Estimated incidence rate (new HIV infection per 1,000 uninfected population) | WCA | Age 0-14 | 2024 | REML | 20 | 0.656 | 95.1 | 0.338 [0.228, 0.502] |
| Estimated incidence rate (new HIV infection per 1,000 uninfected population) | WCA | Age 0-14 | 2024 | PM | 20 | 0.652 | 95.1 | 0.338 [0.228, 0.502] |
| Estimated incidence rate (new HIV infection per 1,000 uninfected population) | WCA | Age 0-14 | 2024 | DL | 20 | 0.883 | 96.3 | 0.340 [0.229, 0.504] |
| Estimated incidence rate (new HIV infection per 1,000 uninfected population) | WCA | Age 15-19 | 2000 | REML | 21 | 1.132 | 88.1 | 1.081 [0.642, 1.818] |
| Estimated incidence rate (new HIV infection per 1,000 uninfected population) | WCA | Age 15-19 | 2000 | PM | 21 | 1.051 | 87.3 | 1.075 [0.638, 1.811] |
| Estimated incidence rate (new HIV infection per 1,000 uninfected population) | WCA | Age 15-19 | 2000 | DL | 21 | 1.421 | 90.3 | 1.095 [0.652, 1.838] |
| Estimated incidence rate (new HIV infection per 1,000 uninfected population) | WCA | Age 15-19 | 2001 | REML | 21 | 1.160 | 88.2 | 0.986 [0.583, 1.667] |
| Estimated incidence rate (new HIV infection per 1,000 uninfected population) | WCA | Age 15-19 | 2001 | PM | 21 | 1.077 | 87.4 | 0.981 [0.580, 1.661] |
| Estimated incidence rate (new HIV infection per 1,000 uninfected population) | WCA | Age 15-19 | 2001 | DL | 21 | 1.456 | 90.4 | 0.999 [0.592, 1.685] |
| Estimated incidence rate (new HIV infection per 1,000 uninfected population) | WCA | Age 15-19 | 2002 | REML | 21 | 1.175 | 88.0 | 0.909 [0.536, 1.541] |
| Estimated incidence rate (new HIV infection per 1,000 uninfected population) | WCA | Age 15-19 | 2002 | PM | 21 | 1.091 | 87.2 | 0.904 [0.533, 1.536] |
| Estimated incidence rate (new HIV infection per 1,000 uninfected population) | WCA | Age 15-19 | 2002 | DL | 21 | 1.490 | 90.3 | 0.921 [0.544, 1.558] |
| Estimated incidence rate (new HIV infection per 1,000 uninfected population) | WCA | Age 15-19 | 2003 | REML | 21 | 1.203 | 88.8 | 0.832 [0.488, 1.419] |
| Estimated incidence rate (new HIV infection per 1,000 uninfected population) | WCA | Age 15-19 | 2003 | PM | 21 | 1.114 | 88.0 | 0.828 [0.486, 1.413] |
| Estimated incidence rate (new HIV infection per 1,000 uninfected population) | WCA | Age 15-19 | 2003 | DL | 21 | 1.648 | 91.6 | 0.846 [0.498, 1.437] |

*Continued on next*

**Supplementary Table S4 (continued)**

| Indicator | Region | Age group | Year | Method | $k$ | $\tau^2$ | $I^2$ (%) | Pooled estimate [95% |
|---|---|---|---|---|---|---|---|---|
| Estimated incidence rate (new HIV infection per 1,000 uninfected population) | WCA | Age 15-19 | 2004 | REML | 21 | 1.232 | 88.4 | 0.767 [0.447, 1.315] |
| Estimated incidence rate (new HIV infection per 1,000 uninfected population) | WCA | Age 15-19 | 2004 | PM | 21 | 1.147 | 87.6 | 0.764 [0.445, 1.311] |
| Estimated incidence rate (new HIV infection per 1,000 uninfected population) | WCA | Age 15-19 | 2004 | DL | 21 | 1.605 | 90.8 | 0.777 [0.455, 1.328] |
| Estimated incidence rate (new HIV infection per 1,000 uninfected population) | WCA | Age 15-19 | 2005 | REML | 21 | 1.316 | 88.6 | 0.706 [0.405, 1.230] |
| Estimated incidence rate (new HIV infection per 1,000 uninfected population) | WCA | Age 15-19 | 2005 | PM | 21 | 1.231 | 87.9 | 0.703 [0.403, 1.227] |
| Estimated incidence rate (new HIV infection per 1,000 uninfected population) | WCA | Age 15-19 | 2005 | DL | 21 | 1.738 | 91.1 | 0.715 [0.412, 1.242] |
| Estimated incidence rate (new HIV infection per 1,000 uninfected population) | WCA | Age 15-19 | 2006 | REML | 21 | 1.410 | 90.1 | 0.648 [0.366, 1.146] |
| Estimated incidence rate (new HIV infection per 1,000 uninfected population) | WCA | Age 15-19 | 2006 | PM | 21 | 1.318 | 89.5 | 0.645 [0.364, 1.143] |
| Estimated incidence rate (new HIV infection per 1,000 uninfected population) | WCA | Age 15-19 | 2006 | DL | 21 | 1.993 | 92.8 | 0.659 [0.374, 1.161] |
| Estimated incidence rate (new HIV infection per 1,000 uninfected population) | WCA | Age 15-19 | 2007 | REML | 21 | 1.536 | 90.0 | 0.604 [0.333, 1.096] |
| Estimated incidence rate (new HIV infection per 1,000 uninfected population) | WCA | Age 15-19 | 2007 | PM | 21 | 1.448 | 89.5 | 0.602 [0.332, 1.093] |
| Estimated incidence rate (new HIV infection per 1,000 uninfected population) | WCA | Age 15-19 | 2007 | DL | 21 | 2.034 | 92.3 | 0.612 [0.339, 1.106] |
| Estimated incidence rate (new HIV infection per 1,000 uninfected population) | WCA | Age 15-19 | 2008 | REML | 21 | 1.533 | 89.8 | 0.557 [0.307, 1.010] |
| Estimated incidence rate (new HIV infection per 1,000 uninfected population) | WCA | Age 15-19 | 2008 | PM | 21 | 1.450 | 89.3 | 0.555 [0.306, 1.008] |
| Estimated incidence rate (new HIV infection per 1,000 uninfected population) | WCA | Age 15-19 | 2008 | DL | 21 | 1.995 | 92.0 | 0.563 [0.312, 1.019] |
| Estimated incidence rate (new HIV infection per 1,000 uninfected population) | WCA | Age 15-19 | 2009 | REML | 21 | 1.664 | 90.4 | 0.516 [0.277, 0.964] |
| Estimated incidence rate (new HIV infection per 1,000 uninfected population) | WCA | Age 15-19 | 2009 | PM | 21 | 1.609 | 90.1 | 0.516 [0.276, 0.963] |
| Estimated incidence rate (new HIV infection per 1,000 uninfected population) | WCA | Age 15-19 | 2009 | DL | 21 | 2.315 | 92.9 | 0.521 [0.280, 0.971] |
| Estimated incidence rate (new HIV infection per 1,000 uninfected population) | WCA | Age 15-19 | 2010 | REML | 21 | 1.727 | 91.6 | 0.484 [0.257, 0.911] |
| Estimated incidence rate (new HIV infection per 1,000 uninfected population) | WCA | Age 15-19 | 2010 | PM | 21 | 1.675 | 91.3 | 0.483 [0.256, 0.910] |
| Estimated incidence rate (new HIV infection per 1,000 uninfected population) | WCA | Age 15-19 | 2010 | DL | 21 | 2.603 | 94.2 | 0.489 [0.260, 0.920] |
| Estimated incidence rate (new HIV infection per 1,000 uninfected population) | WCA | Age 15-19 | 2011 | REML | 21 | 1.809 | 92.1 | 0.447 [0.234, 0.851] |
| Estimated incidence rate (new HIV infection per 1,000 uninfected population) | WCA | Age 15-19 | 2011 | PM | 21 | 1.745 | 91.9 | 0.446 [0.234, 0.850] |
| Estimated incidence rate (new HIV infection per 1,000 uninfected population) | WCA | Age 15-19 | 2011 | DL | 21 | 2.926 | 95.0 | 0.453 [0.238, 0.860] |
| Estimated incidence rate (new HIV infection per 1,000 uninfected population) | WCA | Age 15-19 | 2012 | REML | 21 | 1.800 | 91.7 | 0.416 [0.219, 0.791] |
| Estimated incidence rate (new HIV infection per 1,000 uninfected population) | WCA | Age 15-19 | 2012 | PM | 21 | 1.737 | 91.4 | 0.415 [0.218, 0.790] |
| Estimated incidence rate (new HIV infection per 1,000 uninfected population) | WCA | Age 15-19 | 2012 | DL | 21 | 2.482 | 93.8 | 0.420 [0.221, 0.797] |
| Estimated incidence rate (new HIV infection per 1,000 uninfected population) | WCA | Age 15-19 | 2013 | REML | 21 | 1.835 | 91.2 | 0.391 [0.204, 0.748] |
| Estimated incidence rate (new HIV infection per 1,000 uninfected population) | WCA | Age 15-19 | 2013 | PM | 21 | 1.773 | 90.9 | 0.390 [0.204, 0.747] |
| Estimated incidence rate (new HIV infection per 1,000 uninfected population) | WCA | Age 15-19 | 2013 | DL | 21 | 2.438 | 93.2 | 0.394 [0.207, 0.753] |

*Continued on next p*

**Supplementary Table S4 (continued)**

| Indicator | Region | Age group | Year | Method | $k$ | $\tau^2$ | $I^2$ (%) | Pooled estimate [95% |
|---|---|---|---|---|---|---|---|---|
| Estimated incidence rate (new HIV infection per 1,000 uninfected population) | WCA | Age 15-19 | 2014 | REML | 21 | 1.788 | 90.8 | 0.370 [0.195, 0.702] |
| Estimated incidence rate (new HIV infection per 1,000 uninfected population) | WCA | Age 15-19 | 2014 | PM | 21 | 1.727 | 90.6 | 0.369 [0.194, 0.701] |
| Estimated incidence rate (new HIV infection per 1,000 uninfected population) | WCA | Age 15-19 | 2014 | DL | 21 | 2.383 | 93.0 | 0.373 [0.197, 0.706] |
| Estimated incidence rate (new HIV infection per 1,000 uninfected population) | WCA | Age 15-19 | 2015 | REML | 21 | 1.759 | 90.7 | 0.356 [0.189, 0.674] |
| Estimated incidence rate (new HIV infection per 1,000 uninfected population) | WCA | Age 15-19 | 2015 | PM | 21 | 1.699 | 90.4 | 0.356 [0.188, 0.673] |
| Estimated incidence rate (new HIV infection per 1,000 uninfected population) | WCA | Age 15-19 | 2015 | DL | 21 | 2.326 | 92.8 | 0.360 [0.191, 0.678] |
| Estimated incidence rate (new HIV infection per 1,000 uninfected population) | WCA | Age 15-19 | 2016 | REML | 21 | 1.720 | 90.5 | 0.339 [0.181, 0.635] |
| Estimated incidence rate (new HIV infection per 1,000 uninfected population) | WCA | Age 15-19 | 2016 | PM | 21 | 1.655 | 90.2 | 0.338 [0.181, 0.634] |
| Estimated incidence rate (new HIV infection per 1,000 uninfected population) | WCA | Age 15-19 | 2016 | DL | 21 | 2.297 | 92.7 | 0.342 [0.183, 0.640] |
| Estimated incidence rate (new HIV infection per 1,000 uninfected population) | WCA | Age 15-19 | 2017 | REML | 21 | 1.650 | 90.1 | 0.324 [0.174, 0.601] |
| Estimated incidence rate (new HIV infection per 1,000 uninfected population) | WCA | Age 15-19 | 2017 | PM | 21 | 1.599 | 89.8 | 0.323 [0.174, 0.601] |
| Estimated incidence rate (new HIV infection per 1,000 uninfected population) | WCA | Age 15-19 | 2017 | DL | 21 | 2.157 | 92.2 | 0.327 [0.176, 0.606] |
| Estimated incidence rate (new HIV infection per 1,000 uninfected population) | WCA | Age 15-19 | 2018 | REML | 21 | 1.753 | 90.3 | 0.300 [0.158, 0.566] |
| Estimated incidence rate (new HIV infection per 1,000 uninfected population) | WCA | Age 15-19 | 2018 | PM | 21 | 1.704 | 90.0 | 0.299 [0.158, 0.566] |
| Estimated incidence rate (new HIV infection per 1,000 uninfected population) | WCA | Age 15-19 | 2018 | DL | 21 | 2.067 | 91.6 | 0.301 [0.160, 0.569] |
| Estimated incidence rate (new HIV infection per 1,000 uninfected population) | WCA | Age 15-19 | 2019 | REML | 21 | 1.705 | 89.9 | 0.283 [0.151, 0.531] |
| Estimated incidence rate (new HIV infection per 1,000 uninfected population) | WCA | Age 15-19 | 2019 | PM | 21 | 1.663 | 89.7 | 0.283 [0.151, 0.530] |
| Estimated incidence rate (new HIV infection per 1,000 uninfected population) | WCA | Age 15-19 | 2019 | DL | 21 | 1.999 | 91.2 | 0.284 [0.152, 0.533] |
| Estimated incidence rate (new HIV infection per 1,000 uninfected population) | WCA | Age 15-19 | 2020 | REML | 21 | 1.655 | 89.7 | 0.264 [0.142, 0.491] |
| Estimated incidence rate (new HIV infection per 1,000 uninfected population) | WCA | Age 15-19 | 2020 | PM | 21 | 1.617 | 89.5 | 0.263 [0.141, 0.490] |
| Estimated incidence rate (new HIV infection per 1,000 uninfected population) | WCA | Age 15-19 | 2020 | DL | 21 | 1.890 | 90.8 | 0.265 [0.142, 0.493] |
| Estimated incidence rate (new HIV infection per 1,000 uninfected population) | WCA | Age 15-19 | 2021 | REML | 21 | 1.541 | 89.1 | 0.253 [0.138, 0.463] |
| Estimated incidence rate (new HIV infection per 1,000 uninfected population) | WCA | Age 15-19 | 2021 | PM | 21 | 1.515 | 89.0 | 0.253 [0.138, 0.462] |
| Estimated incidence rate (new HIV infection per 1,000 uninfected population) | WCA | Age 15-19 | 2021 | DL | 21 | 1.637 | 89.7 | 0.253 [0.139, 0.463] |
| Estimated incidence rate (new HIV infection per 1,000 uninfected population) | WCA | Age 15-19 | 2022 | REML | 21 | 1.429 | 87.4 | 0.232 [0.129, 0.415] |
| Estimated incidence rate (new HIV infection per 1,000 uninfected population) | WCA | Age 15-19 | 2022 | PM | 21 | 1.396 | 87.2 | 0.231 [0.129, 0.415] |
| Estimated incidence rate (new HIV infection per 1,000 uninfected population) | WCA | Age 15-19 | 2022 | DL | 21 | 1.571 | 88.4 | 0.232 [0.130, 0.416] |
| Estimated incidence rate (new HIV infection per 1,000 uninfected population) | WCA | Age 15-19 | 2023 | REML | 21 | 1.573 | 89.1 | 0.206 [0.113, 0.377] |
| Estimated incidence rate (new HIV infection per 1,000 uninfected population) | WCA | Age 15-19 | 2023 | PM | 21 | 1.528 | 88.8 | 0.206 [0.112, 0.377] |
| Estimated incidence rate (new HIV infection per 1,000 uninfected population) | WCA | Age 15-19 | 2023 | DL | 21 | 1.778 | 90.3 | 0.207 [0.113, 0.379] |

*Continued on next p*

**Supplementary Table S4 (continued)**

| Indicator | Region | Age group | Year | Method | $k$ | $\tau^2$ | $I^2$ (%) | Pooled estimate [95% |
|---|---|---|---|---|---|---|---|---|
| Estimated incidence rate (new HIV infection per 1,000 uninfected population) | WCA | Age 15-19 | 2024 | REML | 21 | 1.525 | 89.0 | 0.186 [0.103, 0.339] |
| Estimated incidence rate (new HIV infection per 1,000 uninfected population) | WCA | Age 15-19 | 2024 | PM | 21 | 1.484 | 88.7 | 0.186 [0.103, 0.338] |
| Estimated incidence rate (new HIV infection per 1,000 uninfected population) | WCA | Age 15-19 | 2024 | DL | 21 | 1.715 | 90.1 | 0.187 [0.103, 0.340] |
| Estimated mother-to-child transmission rate (%) | ESA | Age 0-4 | 2000 | REML | 21 | 0.103 | 97.6 | 0.759 [0.646, 0.891] |
| Estimated mother-to-child transmission rate (%) | ESA | Age 0-4 | 2000 | PM | 21 | 0.114 | 97.8 | 0.761 [0.647, 0.894] |
| Estimated mother-to-child transmission rate (%) | ESA | Age 0-4 | 2000 | DL | 21 | 0.056 | 95.7 | 0.749 [0.642, 0.874] |
| Estimated mother-to-child transmission rate (%) | ESA | Age 0-4 | 2001 | REML | 21 | 0.104 | 97.8 | 0.746 [0.636, 0.874] |
| Estimated mother-to-child transmission rate (%) | ESA | Age 0-4 | 2001 | PM | 21 | 0.111 | 97.9 | 0.747 [0.636, 0.876] |
| Estimated mother-to-child transmission rate (%) | ESA | Age 0-4 | 2001 | DL | 21 | 0.068 | 96.6 | 0.739 [0.633, 0.864] |
| Estimated mother-to-child transmission rate (%) | ESA | Age 0-4 | 2002 | REML | 21 | 0.106 | 97.9 | 0.730 [0.624, 0.855] |
| Estimated mother-to-child transmission rate (%) | ESA | Age 0-4 | 2002 | PM | 21 | 0.109 | 97.9 | 0.730 [0.624, 0.855] |
| Estimated mother-to-child transmission rate (%) | ESA | Age 0-4 | 2002 | DL | 21 | 0.089 | 97.5 | 0.728 [0.623, 0.851] |
| Estimated mother-to-child transmission rate (%) | ESA | Age 0-4 | 2003 | REML | 21 | 0.108 | 98.0 | 0.712 [0.609, 0.833] |
| Estimated mother-to-child transmission rate (%) | ESA | Age 0-4 | 2003 | PM | 21 | 0.108 | 98.0 | 0.712 [0.609, 0.833] |
| Estimated mother-to-child transmission rate (%) | ESA | Age 0-4 | 2003 | DL | 21 | 0.098 | 97.8 | 0.711 [0.608, 0.832] |
| Estimated mother-to-child transmission rate (%) | ESA | Age 0-4 | 2004 | REML | 21 | 0.126 | 98.3 | 0.682 [0.577, 0.806] |
| Estimated mother-to-child transmission rate (%) | ESA | Age 0-4 | 2004 | PM | 21 | 0.124 | 98.3 | 0.682 [0.577, 0.806] |
| Estimated mother-to-child transmission rate (%) | ESA | Age 0-4 | 2004 | DL | 21 | 0.099 | 97.8 | 0.680 [0.576, 0.803] |
| Estimated mother-to-child transmission rate (%) | ESA | Age 0-4 | 2005 | REML | 21 | 0.201 | 99.1 | 0.619 [0.502, 0.762] |
| Estimated mother-to-child transmission rate (%) | ESA | Age 0-4 | 2005 | PM | 21 | 0.199 | 99.0 | 0.618 [0.502, 0.762] |
| Estimated mother-to-child transmission rate (%) | ESA | Age 0-4 | 2005 | DL | 21 | 0.108 | 98.3 | 0.614 [0.499, 0.757] |
| Estimated mother-to-child transmission rate (%) | ESA | Age 0-4 | 2006 | REML | 21 | 0.266 | 99.4 | 0.559 [0.440, 0.710] |
| Estimated mother-to-child transmission rate (%) | ESA | Age 0-4 | 2006 | PM | 21 | 0.265 | 99.4 | 0.559 [0.440, 0.710] |
| Estimated mother-to-child transmission rate (%) | ESA | Age 0-4 | 2006 | DL | 21 | 0.132 | 98.7 | 0.558 [0.439, 0.707] |
| Estimated mother-to-child transmission rate (%) | ESA | Age 0-4 | 2007 | REML | 21 | 0.273 | 99.4 | 0.500 [0.392, 0.637] |
| Estimated mother-to-child transmission rate (%) | ESA | Age 0-4 | 2007 | PM | 21 | 0.271 | 99.4 | 0.500 [0.392, 0.637] |
| Estimated mother-to-child transmission rate (%) | ESA | Age 0-4 | 2007 | DL | 21 | 0.126 | 98.7 | 0.498 [0.392, 0.634] |
| Estimated mother-to-child transmission rate (%) | ESA | Age 0-4 | 2008 | REML | 21 | 0.316 | 99.4 | 0.450 [0.346, 0.584] |
| Estimated mother-to-child transmission rate (%) | ESA | Age 0-4 | 2008 | PM | 21 | 0.317 | 99.4 | 0.450 [0.346, 0.584] |
| Estimated mother-to-child transmission rate (%) | ESA | Age 0-4 | 2008 | DL | 21 | 0.111 | 98.3 | 0.451 [0.349, 0.584] |

*Continued on next p*

**Supplementary Table S4 (continued)**

| Indicator | Region | Age group | Year | Method | $k$ | $\tau^2$ | $I^2$ (%) | Pooled estimate [95% |
|---|---|---|---|---|---|---|---|---|
| Estimated mother-to-child transmission rate (%) | ESA | Age 0-4 | 2009 | REML | 21 | 0.433 | 99.6 | 0.393 [0.290, 0.533] |
| Estimated mother-to-child transmission rate (%) | ESA | Age 0-4 | 2009 | PM | 21 | 0.435 | 99.6 | 0.393 [0.290, 0.533] |
| Estimated mother-to-child transmission rate (%) | ESA | Age 0-4 | 2009 | DL | 21 | 0.174 | 98.9 | 0.395 [0.292, 0.534] |
| Estimated mother-to-child transmission rate (%) | ESA | Age 0-4 | 2010 | REML | 21 | 0.487 | 99.5 | 0.351 [0.254, 0.484] |
| Estimated mother-to-child transmission rate (%) | ESA | Age 0-4 | 2010 | PM | 21 | 0.487 | 99.5 | 0.351 [0.254, 0.484] |
| Estimated mother-to-child transmission rate (%) | ESA | Age 0-4 | 2010 | DL | 21 | 0.239 | 99.0 | 0.351 [0.255, 0.485] |
| Estimated mother-to-child transmission rate (%) | ESA | Age 0-4 | 2011 | REML | 21 | 0.606 | 99.5 | 0.287 [0.201, 0.410] |
| Estimated mother-to-child transmission rate (%) | ESA | Age 0-4 | 2011 | PM | 21 | 0.602 | 99.5 | 0.287 [0.201, 0.410] |
| Estimated mother-to-child transmission rate (%) | ESA | Age 0-4 | 2011 | DL | 21 | 0.417 | 99.3 | 0.287 [0.201, 0.410] |
| Estimated mother-to-child transmission rate (%) | ESA | Age 0-4 | 2012 | REML | 21 | 0.686 | 99.5 | 0.252 [0.173, 0.369] |
| Estimated mother-to-child transmission rate (%) | ESA | Age 0-4 | 2012 | PM | 21 | 0.683 | 99.5 | 0.252 [0.173, 0.369] |
| Estimated mother-to-child transmission rate (%) | ESA | Age 0-4 | 2012 | DL | 21 | 0.548 | 99.4 | 0.253 [0.173, 0.369] |
| Estimated mother-to-child transmission rate (%) | ESA | Age 0-4 | 2013 | REML | 21 | 0.740 | 99.5 | 0.236 [0.159, 0.351] |
| Estimated mother-to-child transmission rate (%) | ESA | Age 0-4 | 2013 | PM | 21 | 0.739 | 99.5 | 0.236 [0.159, 0.351] |
| Estimated mother-to-child transmission rate (%) | ESA | Age 0-4 | 2013 | DL | 21 | 0.572 | 99.3 | 0.237 [0.160, 0.352] |
| Estimated mother-to-child transmission rate (%) | ESA | Age 0-4 | 2014 | REML | 21 | 0.722 | 99.4 | 0.221 [0.150, 0.326] |
| Estimated mother-to-child transmission rate (%) | ESA | Age 0-4 | 2014 | PM | 21 | 0.719 | 99.4 | 0.221 [0.150, 0.326] |
| Estimated mother-to-child transmission rate (%) | ESA | Age 0-4 | 2014 | DL | 21 | 0.553 | 99.3 | 0.221 [0.150, 0.327] |
| Estimated mother-to-child transmission rate (%) | ESA | Age 0-4 | 2015 | REML | 21 | 0.801 | 99.4 | 0.193 [0.128, 0.291] |
| Estimated mother-to-child transmission rate (%) | ESA | Age 0-4 | 2015 | PM | 21 | 0.801 | 99.4 | 0.193 [0.128, 0.291] |
| Estimated mother-to-child transmission rate (%) | ESA | Age 0-4 | 2015 | DL | 21 | 0.520 | 99.1 | 0.193 [0.128, 0.292] |
| Estimated mother-to-child transmission rate (%) | ESA | Age 0-4 | 2016 | REML | 21 | 0.866 | 99.5 | 0.182 [0.119, 0.280] |
| Estimated mother-to-child transmission rate (%) | ESA | Age 0-4 | 2016 | PM | 21 | 0.866 | 99.5 | 0.182 [0.119, 0.280] |
| Estimated mother-to-child transmission rate (%) | ESA | Age 0-4 | 2016 | DL | 21 | 0.562 | 99.2 | 0.183 [0.119, 0.281] |
| Estimated mother-to-child transmission rate (%) | ESA | Age 0-4 | 2017 | REML | 21 | 0.964 | 99.4 | 0.166 [0.105, 0.260] |
| Estimated mother-to-child transmission rate (%) | ESA | Age 0-4 | 2017 | PM | 21 | 0.968 | 99.4 | 0.166 [0.105, 0.260] |
| Estimated mother-to-child transmission rate (%) | ESA | Age 0-4 | 2017 | DL | 21 | 0.669 | 99.2 | 0.166 [0.106, 0.261] |
| Estimated mother-to-child transmission rate (%) | ESA | Age 0-4 | 2018 | REML | 21 | 1.164 | 99.5 | 0.140 [0.085, 0.229] |
| Estimated mother-to-child transmission rate (%) | ESA | Age 0-4 | 2018 | PM | 21 | 1.159 | 99.5 | 0.140 [0.085, 0.229] |
| Estimated mother-to-child transmission rate (%) | ESA | Age 0-4 | 2018 | DL | 21 | 1.029 | 99.4 | 0.140 [0.085, 0.229] |

*Continued on next p*

**Supplementary Table S4 (continued)**

| Indicator | Region | Age group | Year | Method | $k$ | $\tau^2$ | $I^2$ (%) | Pooled estimate [95% |
|---|---|---|---|---|---|---|---|---|
| Estimated mother-to-child transmission rate (%) | ESA | Age 0-4 | 2019 | REML | 21 | 1.239 | 99.4 | 0.130 [0.078, 0.216] |
| Estimated mother-to-child transmission rate (%) | ESA | Age 0-4 | 2019 | PM | 21 | 1.234 | 99.4 | 0.130 [0.078, 0.216] |
| Estimated mother-to-child transmission rate (%) | ESA | Age 0-4 | 2019 | DL | 21 | 1.301 | 99.5 | 0.130 [0.078, 0.216] |
| Estimated mother-to-child transmission rate (%) | ESA | Age 0-4 | 2020 | REML | 21 | 1.260 | 99.4 | 0.122 [0.073, 0.205] |
| Estimated mother-to-child transmission rate (%) | ESA | Age 0-4 | 2020 | PM | 21 | 1.254 | 99.4 | 0.122 [0.073, 0.205] |
| Estimated mother-to-child transmission rate (%) | ESA | Age 0-4 | 2020 | DL | 21 | 1.582 | 99.5 | 0.122 [0.073, 0.204] |
| Estimated mother-to-child transmission rate (%) | ESA | Age 0-4 | 2021 | REML | 21 | 1.248 | 99.3 | 0.129 [0.077, 0.215] |
| Estimated mother-to-child transmission rate (%) | ESA | Age 0-4 | 2021 | PM | 21 | 1.244 | 99.3 | 0.129 [0.077, 0.215] |
| Estimated mother-to-child transmission rate (%) | ESA | Age 0-4 | 2021 | DL | 21 | 1.380 | 99.4 | 0.129 [0.077, 0.215] |
| Estimated mother-to-child transmission rate (%) | ESA | Age 0-4 | 2022 | REML | 21 | 1.197 | 99.0 | 0.129 [0.078, 0.214] |
| Estimated mother-to-child transmission rate (%) | ESA | Age 0-4 | 2022 | PM | 21 | 1.192 | 99.0 | 0.129 [0.078, 0.214] |
| Estimated mother-to-child transmission rate (%) | ESA | Age 0-4 | 2022 | DL | 21 | 1.312 | 99.1 | 0.129 [0.078, 0.214] |
| Estimated mother-to-child transmission rate (%) | ESA | Age 0-4 | 2023 | REML | 20 | 1.207 | 98.8 | 0.120 [0.071, 0.201] |
| Estimated mother-to-child transmission rate (%) | ESA | Age 0-4 | 2023 | PM | 20 | 1.206 | 98.8 | 0.120 [0.071, 0.201] |
| Estimated mother-to-child transmission rate (%) | ESA | Age 0-4 | 2023 | DL | 20 | 1.401 | 99.0 | 0.119 [0.071, 0.201] |
| Estimated mother-to-child transmission rate (%) | ESA | Age 0-4 | 2024 | REML | 21 | 1.268 | 98.9 | 0.110 [0.065, 0.185] |
| Estimated mother-to-child transmission rate (%) | ESA | Age 0-4 | 2024 | PM | 21 | 1.269 | 98.9 | 0.110 [0.065, 0.185] |
| Estimated mother-to-child transmission rate (%) | ESA | Age 0-4 | 2024 | DL | 21 | 1.487 | 99.1 | 0.110 [0.065, 0.184] |
| Estimated mother-to-child transmission rate (%) | WCA | Age 0-4 | 2000 | REML | 20 | 0.041 | 94.5 | 0.761 [0.690, 0.841] |
| Estimated mother-to-child transmission rate (%) | WCA | Age 0-4 | 2000 | PM | 20 | 0.042 | 94.6 | 0.762 [0.690, 0.841] |
| Estimated mother-to-child transmission rate (%) | WCA | Age 0-4 | 2000 | DL | 20 | 0.040 | 94.4 | 0.761 [0.690, 0.841] |
| Estimated mother-to-child transmission rate (%) | WCA | Age 0-4 | 2001 | REML | 20 | 0.034 | 94.2 | 0.742 [0.679, 0.811] |
| Estimated mother-to-child transmission rate (%) | WCA | Age 0-4 | 2001 | PM | 20 | 0.034 | 94.2 | 0.742 [0.679, 0.811] |
| Estimated mother-to-child transmission rate (%) | WCA | Age 0-4 | 2001 | DL | 20 | 0.034 | 94.3 | 0.742 [0.679, 0.811] |
| Estimated mother-to-child transmission rate (%) | WCA | Age 0-4 | 2002 | REML | 20 | 0.031 | 93.9 | 0.728 [0.669, 0.793] |
| Estimated mother-to-child transmission rate (%) | WCA | Age 0-4 | 2002 | PM | 20 | 0.031 | 93.9 | 0.728 [0.669, 0.793] |
| Estimated mother-to-child transmission rate (%) | WCA | Age 0-4 | 2002 | DL | 20 | 0.031 | 94.0 | 0.728 [0.669, 0.793] |
| Estimated mother-to-child transmission rate (%) | WCA | Age 0-4 | 2003 | REML | 20 | 0.029 | 94.0 | 0.716 [0.659, 0.777] |
| Estimated mother-to-child transmission rate (%) | WCA | Age 0-4 | 2003 | PM | 20 | 0.029 | 94.0 | 0.716 [0.659, 0.777] |
| Estimated mother-to-child transmission rate (%) | WCA | Age 0-4 | 2003 | DL | 20 | 0.029 | 94.1 | 0.716 [0.659, 0.778] |

*Continued on next*

**Supplementary Table S4 (continued)**

| Indicator | Region | Age group | Year | Method | $k$ | $\tau^2$ | $I^2$ (%) | Pooled estimate [95% |
|---|---|---|---|---|---|---|---|---|
| Estimated mother-to-child transmission rate (%) | WCA | Age 0-4 | 2004 | REML | 20 | 0.030 | 94.4 | 0.701 [0.645, 0.762] |
| Estimated mother-to-child transmission rate (%) | WCA | Age 0-4 | 2004 | PM | 20 | 0.030 | 94.4 | 0.701 [0.645, 0.762] |
| Estimated mother-to-child transmission rate (%) | WCA | Age 0-4 | 2004 | DL | 20 | 0.032 | 94.7 | 0.701 [0.645, 0.762] |
| Estimated mother-to-child transmission rate (%) | WCA | Age 0-4 | 2005 | REML | 20 | 0.029 | 94.6 | 0.684 [0.630, 0.743] |
| Estimated mother-to-child transmission rate (%) | WCA | Age 0-4 | 2005 | PM | 20 | 0.029 | 94.6 | 0.684 [0.630, 0.743] |
| Estimated mother-to-child transmission rate (%) | WCA | Age 0-4 | 2005 | DL | 20 | 0.029 | 94.7 | 0.684 [0.630, 0.743] |
| Estimated mother-to-child transmission rate (%) | WCA | Age 0-4 | 2006 | REML | 20 | 0.029 | 95.4 | 0.658 [0.606, 0.714] |
| Estimated mother-to-child transmission rate (%) | WCA | Age 0-4 | 2006 | PM | 20 | 0.029 | 95.4 | 0.658 [0.606, 0.714] |
| Estimated mother-to-child transmission rate (%) | WCA | Age 0-4 | 2006 | DL | 20 | 0.030 | 95.5 | 0.658 [0.606, 0.714] |
| Estimated mother-to-child transmission rate (%) | WCA | Age 0-4 | 2007 | REML | 20 | 0.033 | 95.7 | 0.608 [0.558, 0.663] |
| Estimated mother-to-child transmission rate (%) | WCA | Age 0-4 | 2007 | PM | 20 | 0.032 | 95.7 | 0.608 [0.558, 0.663] |
| Estimated mother-to-child transmission rate (%) | WCA | Age 0-4 | 2007 | DL | 20 | 0.035 | 95.9 | 0.608 [0.558, 0.663] |
| Estimated mother-to-child transmission rate (%) | WCA | Age 0-4 | 2008 | REML | 20 | 0.049 | 96.9 | 0.547 [0.493, 0.608] |
| Estimated mother-to-child transmission rate (%) | WCA | Age 0-4 | 2008 | PM | 20 | 0.049 | 96.9 | 0.547 [0.493, 0.608] |
| Estimated mother-to-child transmission rate (%) | WCA | Age 0-4 | 2008 | DL | 20 | 0.046 | 96.7 | 0.547 [0.493, 0.608] |
| Estimated mother-to-child transmission rate (%) | WCA | Age 0-4 | 2009 | REML | 20 | 0.052 | 96.8 | 0.512 [0.459, 0.571] |
| Estimated mother-to-child transmission rate (%) | WCA | Age 0-4 | 2009 | PM | 20 | 0.052 | 96.8 | 0.512 [0.459, 0.571] |
| Estimated mother-to-child transmission rate (%) | WCA | Age 0-4 | 2009 | DL | 20 | 0.051 | 96.7 | 0.512 [0.459, 0.571] |
| Estimated mother-to-child transmission rate (%) | WCA | Age 0-4 | 2010 | REML | 20 | 0.114 | 98.0 | 0.420 [0.356, 0.494] |
| Estimated mother-to-child transmission rate (%) | WCA | Age 0-4 | 2010 | PM | 20 | 0.118 | 98.1 | 0.420 [0.356, 0.494] |
| Estimated mother-to-child transmission rate (%) | WCA | Age 0-4 | 2010 | DL | 20 | 0.077 | 97.1 | 0.421 [0.358, 0.495] |
| Estimated mother-to-child transmission rate (%) | WCA | Age 0-4 | 2011 | REML | 20 | 0.107 | 97.3 | 0.368 [0.314, 0.430] |
| Estimated mother-to-child transmission rate (%) | WCA | Age 0-4 | 2011 | PM | 20 | 0.107 | 97.3 | 0.368 [0.314, 0.430] |
| Estimated mother-to-child transmission rate (%) | WCA | Age 0-4 | 2011 | DL | 20 | 0.095 | 97.0 | 0.368 [0.315, 0.430] |
| Estimated mother-to-child transmission rate (%) | WCA | Age 0-4 | 2012 | REML | 20 | 0.131 | 97.6 | 0.343 [0.289, 0.408] |
| Estimated mother-to-child transmission rate (%) | WCA | Age 0-4 | 2012 | PM | 20 | 0.131 | 97.6 | 0.343 [0.289, 0.408] |
| Estimated mother-to-child transmission rate (%) | WCA | Age 0-4 | 2012 | DL | 20 | 0.139 | 97.7 | 0.343 [0.288, 0.408] |
| Estimated mother-to-child transmission rate (%) | WCA | Age 0-4 | 2013 | REML | 20 | 0.167 | 97.7 | 0.313 [0.257, 0.380] |
| Estimated mother-to-child transmission rate (%) | WCA | Age 0-4 | 2013 | PM | 20 | 0.167 | 97.7 | 0.313 [0.257, 0.380] |
| Estimated mother-to-child transmission rate (%) | WCA | Age 0-4 | 2013 | DL | 20 | 0.173 | 97.8 | 0.313 [0.257, 0.380] |

*Continued on next p*

**Supplementary Table S4 (continued)**

| Indicator | Region | Age group | Year | Method | $k$ | $\tau^2$ | $I^2$ (%) | Pooled estimate [95% |
|---|---|---|---|---|---|---|---|---|
| Estimated mother-to-child transmission rate (%) | WCA | Age 0-4 | 2014 | REML | 20 | 0.228 | 97.7 | 0.276 [0.219, 0.346] |
| Estimated mother-to-child transmission rate (%) | WCA | Age 0-4 | 2014 | PM | 20 | 0.228 | 97.7 | 0.276 [0.219, 0.346] |
| Estimated mother-to-child transmission rate (%) | WCA | Age 0-4 | 2014 | DL | 20 | 0.197 | 97.4 | 0.276 [0.220, 0.347] |
| Estimated mother-to-child transmission rate (%) | WCA | Age 0-4 | 2015 | REML | 20 | 0.219 | 97.5 | 0.272 [0.218, 0.340] |
| Estimated mother-to-child transmission rate (%) | WCA | Age 0-4 | 2015 | PM | 20 | 0.218 | 97.5 | 0.272 [0.218, 0.340] |
| Estimated mother-to-child transmission rate (%) | WCA | Age 0-4 | 2015 | DL | 20 | 0.224 | 97.6 | 0.272 [0.218, 0.340] |
| Estimated mother-to-child transmission rate (%) | WCA | Age 0-4 | 2016 | REML | 20 | 0.236 | 97.8 | 0.279 [0.222, 0.351] |
| Estimated mother-to-child transmission rate (%) | WCA | Age 0-4 | 2016 | PM | 20 | 0.233 | 97.8 | 0.279 [0.222, 0.351] |
| Estimated mother-to-child transmission rate (%) | WCA | Age 0-4 | 2016 | DL | 20 | 0.290 | 98.2 | 0.279 [0.222, 0.351] |
| Estimated mother-to-child transmission rate (%) | WCA | Age 0-4 | 2017 | REML | 20 | 0.234 | 97.7 | 0.287 [0.228, 0.362] |
| Estimated mother-to-child transmission rate (%) | WCA | Age 0-4 | 2017 | PM | 20 | 0.233 | 97.7 | 0.287 [0.228, 0.362] |
| Estimated mother-to-child transmission rate (%) | WCA | Age 0-4 | 2017 | DL | 20 | 0.270 | 98.0 | 0.287 [0.228, 0.361] |
| Estimated mother-to-child transmission rate (%) | WCA | Age 0-4 | 2018 | REML | 20 | 0.276 | 97.9 | 0.270 [0.211, 0.347] |
| Estimated mother-to-child transmission rate (%) | WCA | Age 0-4 | 2018 | PM | 20 | 0.273 | 97.9 | 0.270 [0.211, 0.347] |
| Estimated mother-to-child transmission rate (%) | WCA | Age 0-4 | 2018 | DL | 20 | 0.376 | 98.4 | 0.270 [0.211, 0.346] |
| Estimated mother-to-child transmission rate (%) | WCA | Age 0-4 | 2019 | REML | 20 | 0.223 | 97.0 | 0.247 [0.197, 0.310] |
| Estimated mother-to-child transmission rate (%) | WCA | Age 0-4 | 2019 | PM | 20 | 0.222 | 96.9 | 0.247 [0.197, 0.310] |
| Estimated mother-to-child transmission rate (%) | WCA | Age 0-4 | 2019 | DL | 20 | 0.353 | 98.1 | 0.246 [0.196, 0.309] |
| Estimated mother-to-child transmission rate (%) | WCA | Age 0-4 | 2020 | REML | 20 | 0.201 | 96.2 | 0.258 [0.208, 0.321] |
| Estimated mother-to-child transmission rate (%) | WCA | Age 0-4 | 2020 | PM | 20 | 0.201 | 96.2 | 0.258 [0.208, 0.321] |
| Estimated mother-to-child transmission rate (%) | WCA | Age 0-4 | 2020 | DL | 20 | 0.270 | 97.1 | 0.257 [0.207, 0.319] |
| Estimated mother-to-child transmission rate (%) | WCA | Age 0-4 | 2021 | REML | 20 | 0.271 | 97.2 | 0.238 [0.185, 0.306] |
| Estimated mother-to-child transmission rate (%) | WCA | Age 0-4 | 2021 | PM | 20 | 0.276 | 97.3 | 0.238 [0.185, 0.306] |
| Estimated mother-to-child transmission rate (%) | WCA | Age 0-4 | 2021 | DL | 20 | 0.335 | 97.7 | 0.237 [0.184, 0.306] |
| Estimated mother-to-child transmission rate (%) | WCA | Age 0-4 | 2022 | REML | 20 | 0.231 | 96.1 | 0.241 [0.190, 0.305] |
| Estimated mother-to-child transmission rate (%) | WCA | Age 0-4 | 2022 | PM | 20 | 0.238 | 96.2 | 0.241 [0.190, 0.305] |
| Estimated mother-to-child transmission rate (%) | WCA | Age 0-4 | 2022 | DL | 20 | 0.206 | 95.7 | 0.241 [0.191, 0.306] |
| Estimated mother-to-child transmission rate (%) | WCA | Age 0-4 | 2023 | REML | 20 | 0.181 | 94.3 | 0.231 [0.187, 0.286] |
| Estimated mother-to-child transmission rate (%) | WCA | Age 0-4 | 2023 | PM | 20 | 0.186 | 94.5 | 0.231 [0.187, 0.286] |
| Estimated mother-to-child transmission rate (%) | WCA | Age 0-4 | 2023 | DL | 20 | 0.153 | 93.4 | 0.232 [0.188, 0.287] |

*Continued on next p*

**Supplementary Table S4 (continued)**

| Indicator | Region | Age group | Year | Method | $k$ | $\tau^2$ | $I^2$ (%) | Pooled estimate [95% |
|---|---|---|---|---|---|---|---|---|
| Estimated mother-to-child transmission rate (%) | WCA | Age 0-4 | 2024 | REML | 20 | 0.159 | 92.1 | 0.213 [0.174, 0.261] |
| Estimated mother-to-child transmission rate (%) | WCA | Age 0-4 | 2024 | PM | 20 | 0.159 | 92.1 | 0.213 [0.174, 0.261] |
| Estimated mother-to-child transmission rate (%) | WCA | Age 0-4 | 2024 | DL | 20 | 0.146 | 91.5 | 0.214 [0.175, 0.262] |
| Estimated rate of annual AIDS-related deaths (per 100,000 population) | ESA | Age 0-14 | 2000 | REML | 22 | 3.147 | 99.8 | 94.615 [42.815, 209.085 |
| Estimated rate of annual AIDS-related deaths (per 100,000 population) | ESA | Age 0-14 | 2000 | PM | 22 | 3.175 | 99.8 | 94.606 [42.810, 209.072 |
| Estimated rate of annual AIDS-related deaths (per 100,000 population) | ESA | Age 0-14 | 2000 | DL | 22 | 0.749 | 99.3 | 97.707 [44.685, 213.645 |
| Estimated rate of annual AIDS-related deaths (per 100,000 population) | ESA | Age 0-14 | 2001 | REML | 22 | 3.111 | 99.8 | 95.343 [43.385, 209.526 |
| Estimated rate of annual AIDS-related deaths (per 100,000 population) | ESA | Age 0-14 | 2001 | PM | 22 | 3.132 | 99.8 | 95.337 [43.381, 209.517 |
| Estimated rate of annual AIDS-related deaths (per 100,000 population) | ESA | Age 0-14 | 2001 | DL | 22 | 0.815 | 99.4 | 97.734 [44.798, 213.221 |
| Estimated rate of annual AIDS-related deaths (per 100,000 population) | ESA | Age 0-14 | 2002 | REML | 22 | 3.072 | 99.8 | 94.984 [43.477, 207.508 |
| Estimated rate of annual AIDS-related deaths (per 100,000 population) | ESA | Age 0-14 | 2002 | PM | 22 | 3.086 | 99.8 | 94.980 [43.475, 207.502 |
| Estimated rate of annual AIDS-related deaths (per 100,000 population) | ESA | Age 0-14 | 2002 | DL | 22 | 0.872 | 99.4 | 96.816 [44.523, 210.528 |
| Estimated rate of annual AIDS-related deaths (per 100,000 population) | ESA | Age 0-14 | 2003 | REML | 22 | 3.022 | 99.8 | 93.875 [43.260, 203.712 |
| Estimated rate of annual AIDS-related deaths (per 100,000 population) | ESA | Age 0-14 | 2003 | PM | 22 | 3.034 | 99.8 | 93.873 [43.258, 203.708 |
| Estimated rate of annual AIDS-related deaths (per 100,000 population) | ESA | Age 0-14 | 2003 | DL | 22 | 0.928 | 99.5 | 95.374 [44.103, 206.251 |
| Estimated rate of annual AIDS-related deaths (per 100,000 population) | ESA | Age 0-14 | 2004 | REML | 22 | 2.975 | 99.8 | 91.758 [42.568, 197.789 |
| Estimated rate of annual AIDS-related deaths (per 100,000 population) | ESA | Age 0-14 | 2004 | PM | 22 | 2.982 | 99.8 | 91.756 [42.567, 197.786 |
| Estimated rate of annual AIDS-related deaths (per 100,000 population) | ESA | Age 0-14 | 2004 | DL | 22 | 1.001 | 99.5 | 92.861 [43.165, 199.773 |
| Estimated rate of annual AIDS-related deaths (per 100,000 population) | ESA | Age 0-14 | 2005 | REML | 22 | 2.908 | 99.8 | 87.802 [41.104, 187.555 |
| Estimated rate of annual AIDS-related deaths (per 100,000 population) | ESA | Age 0-14 | 2005 | PM | 22 | 2.913 | 99.8 | 87.802 [41.103, 187.554 |
| Estimated rate of annual AIDS-related deaths (per 100,000 population) | ESA | Age 0-14 | 2005 | DL | 22 | 1.133 | 99.6 | 88.543 [41.496, 188.931 |
| Estimated rate of annual AIDS-related deaths (per 100,000 population) | ESA | Age 0-14 | 2006 | REML | 22 | 2.782 | 99.8 | 81.604 [38.851, 171.405 |
| Estimated rate of annual AIDS-related deaths (per 100,000 population) | ESA | Age 0-14 | 2006 | PM | 22 | 2.784 | 99.8 | 81.604 [38.851, 171.404 |
| Estimated rate of annual AIDS-related deaths (per 100,000 population) | ESA | Age 0-14 | 2006 | DL | 22 | 1.181 | 99.5 | 82.096 [39.102, 172.364 |
| Estimated rate of annual AIDS-related deaths (per 100,000 population) | ESA | Age 0-14 | 2007 | REML | 22 | 2.698 | 99.8 | 75.438 [36.326, 156.663 |
| Estimated rate of annual AIDS-related deaths (per 100,000 population) | ESA | Age 0-14 | 2007 | PM | 22 | 2.699 | 99.8 | 75.438 [36.326, 156.663 |
| Estimated rate of annual AIDS-related deaths (per 100,000 population) | ESA | Age 0-14 | 2007 | DL | 22 | 1.260 | 99.5 | 75.791 [36.501, 157.370 |
| Estimated rate of annual AIDS-related deaths (per 100,000 population) | ESA | Age 0-14 | 2008 | REML | 22 | 2.601 | 99.8 | 68.898 [33.624, 141.177 |
| Estimated rate of annual AIDS-related deaths (per 100,000 population) | ESA | Age 0-14 | 2008 | PM | 22 | 2.600 | 99.8 | 68.898 [33.624, 141.178 |
| Estimated rate of annual AIDS-related deaths (per 100,000 population) | ESA | Age 0-14 | 2008 | DL | 22 | 1.427 | 99.5 | 69.107 [33.722, 141.620 |

*Continued on next p*

**Supplementary Table S4 (continued)**

| Indicator | Region | Age group | Year | Method | $k$ | $\tau^2$ | $I^2$ (%) | Pooled estimate [95% |
|---|---|---|---|---|---|---|---|---|
| Estimated rate of annual AIDS-related deaths (per 100,000 population) | ESA | Age 0-14 | 2009 | REML | 22 | 2.458 | 99.7 | 60.649 [30.193, 121.827 |
| Estimated rate of annual AIDS-related deaths (per 100,000 population) | ESA | Age 0-14 | 2009 | PM | 22 | 2.456 | 99.7 | 60.649 [30.193, 121.827 |
| Estimated rate of annual AIDS-related deaths (per 100,000 population) | ESA | Age 0-14 | 2009 | DL | 22 | 1.569 | 99.5 | 60.761 [30.244, 122.070 |
| Estimated rate of annual AIDS-related deaths (per 100,000 population) | ESA | Age 0-14 | 2010 | REML | 22 | 2.339 | 99.6 | 52.926 [26.795, 104.541 |
| Estimated rate of annual AIDS-related deaths (per 100,000 population) | ESA | Age 0-14 | 2010 | PM | 22 | 2.336 | 99.6 | 52.926 [26.795, 104.541 |
| Estimated rate of annual AIDS-related deaths (per 100,000 population) | ESA | Age 0-14 | 2010 | DL | 22 | 1.653 | 99.5 | 53.002 [26.829, 104.707 |
| Estimated rate of annual AIDS-related deaths (per 100,000 population) | ESA | Age 0-14 | 2011 | REML | 22 | 2.176 | 99.5 | 45.489 [23.588, 87.723] |
| Estimated rate of annual AIDS-related deaths (per 100,000 population) | ESA | Age 0-14 | 2011 | PM | 22 | 2.173 | 99.5 | 45.489 [23.588, 87.724] |
| Estimated rate of annual AIDS-related deaths (per 100,000 population) | ESA | Age 0-14 | 2011 | DL | 22 | 1.633 | 99.4 | 45.545 [23.614, 87.843] |
| Estimated rate of annual AIDS-related deaths (per 100,000 population) | ESA | Age 0-14 | 2012 | REML | 22 | 2.078 | 99.5 | 38.475 [20.244, 73.125] |
| Estimated rate of annual AIDS-related deaths (per 100,000 population) | ESA | Age 0-14 | 2012 | PM | 22 | 2.075 | 99.5 | 38.476 [20.244, 73.126] |
| Estimated rate of annual AIDS-related deaths (per 100,000 population) | ESA | Age 0-14 | 2012 | DL | 22 | 1.661 | 99.4 | 38.513 [20.262, 73.205] |
| Estimated rate of annual AIDS-related deaths (per 100,000 population) | ESA | Age 0-14 | 2013 | REML | 22 | 1.975 | 99.4 | 33.515 [17.910, 62.716] |
| Estimated rate of annual AIDS-related deaths (per 100,000 population) | ESA | Age 0-14 | 2013 | PM | 22 | 1.973 | 99.4 | 33.515 [17.910, 62.716] |
| Estimated rate of annual AIDS-related deaths (per 100,000 population) | ESA | Age 0-14 | 2013 | DL | 22 | 1.640 | 99.3 | 33.543 [17.924, 62.772] |
| Estimated rate of annual AIDS-related deaths (per 100,000 population) | ESA | Age 0-14 | 2014 | REML | 22 | 1.943 | 99.4 | 29.725 [15.955, 55.379] |
| Estimated rate of annual AIDS-related deaths (per 100,000 population) | ESA | Age 0-14 | 2014 | PM | 22 | 1.942 | 99.4 | 29.725 [15.955, 55.379] |
| Estimated rate of annual AIDS-related deaths (per 100,000 population) | ESA | Age 0-14 | 2014 | DL | 22 | 1.722 | 99.3 | 29.742 [15.964, 55.411] |
| Estimated rate of annual AIDS-related deaths (per 100,000 population) | ESA | Age 0-14 | 2015 | REML | 22 | 1.940 | 99.3 | 25.957 [13.930, 48.368] |
| Estimated rate of annual AIDS-related deaths (per 100,000 population) | ESA | Age 0-14 | 2015 | PM | 22 | 1.940 | 99.3 | 25.957 [13.930, 48.368] |
| Estimated rate of annual AIDS-related deaths (per 100,000 population) | ESA | Age 0-14 | 2015 | DL | 22 | 1.787 | 99.2 | 25.969 [13.936, 48.389] |
| Estimated rate of annual AIDS-related deaths (per 100,000 population) | ESA | Age 0-14 | 2016 | REML | 22 | 1.842 | 99.2 | 22.722 [12.394, 41.659] |
| Estimated rate of annual AIDS-related deaths (per 100,000 population) | ESA | Age 0-14 | 2016 | PM | 22 | 1.837 | 99.2 | 22.722 [12.394, 41.659] |
| Estimated rate of annual AIDS-related deaths (per 100,000 population) | ESA | Age 0-14 | 2016 | DL | 22 | 1.922 | 99.2 | 22.719 [12.392, 41.651] |
| Estimated rate of annual AIDS-related deaths (per 100,000 population) | ESA | Age 0-14 | 2017 | REML | 22 | 1.773 | 98.9 | 19.582 [10.780, 35.571] |
| Estimated rate of annual AIDS-related deaths (per 100,000 population) | ESA | Age 0-14 | 2017 | PM | 22 | 1.774 | 98.9 | 19.582 [10.779, 35.571] |
| Estimated rate of annual AIDS-related deaths (per 100,000 population) | ESA | Age 0-14 | 2017 | DL | 22 | 1.917 | 99.0 | 19.571 [10.774, 35.553] |
| Estimated rate of annual AIDS-related deaths (per 100,000 population) | ESA | Age 0-14 | 2018 | REML | 22 | 1.682 | 98.8 | 16.950 [9.460, 30.369] |
| Estimated rate of annual AIDS-related deaths (per 100,000 population) | ESA | Age 0-14 | 2018 | PM | 22 | 1.686 | 98.8 | 16.950 [9.460, 30.369] |
| Estimated rate of annual AIDS-related deaths (per 100,000 population) | ESA | Age 0-14 | 2018 | DL | 22 | 1.857 | 98.9 | 16.937 [9.452, 30.349] |

*Continued on next*

**Supplementary Table S4 (continued)**

| Indicator | Region | Age group | Year | Method | $k$ | $\tau^2$ | $I^2$ (%) | Pooled estimate [95% |
|---|---|---|---|---|---|---|---|---|
| Estimated rate of annual AIDS-related deaths (per 100,000 population) | ESA | Age 0-14 | 2019 | REML | 22 | 1.677 | 98.7 | 15.451 [8.623, 27.686] |
| Estimated rate of annual AIDS-related deaths (per 100,000 population) | ESA | Age 0-14 | 2019 | PM | 22 | 1.682 | 98.7 | 15.451 [8.623, 27.685] |
| Estimated rate of annual AIDS-related deaths (per 100,000 population) | ESA | Age 0-14 | 2019 | DL | 22 | 1.873 | 98.8 | 15.439 [8.615, 27.666] |
| Estimated rate of annual AIDS-related deaths (per 100,000 population) | ESA | Age 0-14 | 2020 | REML | 22 | 1.692 | 98.6 | 13.991 [7.790, 25.127] |
| Estimated rate of annual AIDS-related deaths (per 100,000 population) | ESA | Age 0-14 | 2020 | PM | 22 | 1.698 | 98.6 | 13.990 [7.790, 25.127] |
| Estimated rate of annual AIDS-related deaths (per 100,000 population) | ESA | Age 0-14 | 2020 | DL | 22 | 1.931 | 98.8 | 13.977 [7.781, 25.106] |
| Estimated rate of annual AIDS-related deaths (per 100,000 population) | ESA | Age 0-14 | 2021 | REML | 22 | 1.515 | 98.3 | 12.281 [7.043, 21.415] |
| Estimated rate of annual AIDS-related deaths (per 100,000 population) | ESA | Age 0-14 | 2021 | PM | 22 | 1.522 | 98.3 | 12.281 [7.043, 21.415] |
| Estimated rate of annual AIDS-related deaths (per 100,000 population) | ESA | Age 0-14 | 2021 | DL | 22 | 1.841 | 98.6 | 12.263 [7.031, 21.389] |
| Estimated rate of annual AIDS-related deaths (per 100,000 population) | ESA | Age 0-14 | 2022 | REML | 22 | 1.534 | 98.1 | 11.435 [6.520, 20.057] |
| Estimated rate of annual AIDS-related deaths (per 100,000 population) | ESA | Age 0-14 | 2022 | PM | 22 | 1.550 | 98.1 | 11.434 [6.519, 20.055] |
| Estimated rate of annual AIDS-related deaths (per 100,000 population) | ESA | Age 0-14 | 2022 | DL | 22 | 1.757 | 98.3 | 11.419 [6.508, 20.036] |
| Estimated rate of annual AIDS-related deaths (per 100,000 population) | ESA | Age 0-14 | 2023 | REML | 21 | 1.160 | 96.8 | 12.201 [7.334, 20.299] |
| Estimated rate of annual AIDS-related deaths (per 100,000 population) | ESA | Age 0-14 | 2023 | PM | 21 | 1.190 | 96.9 | 12.196 [7.330, 20.293] |
| Estimated rate of annual AIDS-related deaths (per 100,000 population) | ESA | Age 0-14 | 2023 | DL | 21 | 0.842 | 95.7 | 12.280 [7.397, 20.384] |
| Estimated rate of annual AIDS-related deaths (per 100,000 population) | ESA | Age 0-14 | 2024 | REML | 21 | 1.076 | 96.2 | 11.298 [6.925, 18.433] |
| Estimated rate of annual AIDS-related deaths (per 100,000 population) | ESA | Age 0-14 | 2024 | PM | 21 | 1.095 | 96.3 | 11.294 [6.922, 18.428] |
| Estimated rate of annual AIDS-related deaths (per 100,000 population) | ESA | Age 0-14 | 2024 | DL | 21 | 0.823 | 95.1 | 11.363 [6.973, 18.515] |
| Estimated rate of annual AIDS-related deaths (per 100,000 population) | ESA | Age 15-19 | 2000 | REML | 23 | 4.659 | 99.9 | 8.445 [3.287, 21.695] |
| Estimated rate of annual AIDS-related deaths (per 100,000 population) | ESA | Age 15-19 | 2000 | PM | 23 | 4.727 | 99.9 | 8.443 [3.286, 21.691] |
| Estimated rate of annual AIDS-related deaths (per 100,000 population) | ESA | Age 15-19 | 2000 | DL | 23 | 0.523 | 99.3 | 9.603 [3.909, 23.592] |
| Estimated rate of annual AIDS-related deaths (per 100,000 population) | ESA | Age 15-19 | 2001 | REML | 23 | 4.355 | 99.9 | 9.446 [3.788, 23.554] |
| Estimated rate of annual AIDS-related deaths (per 100,000 population) | ESA | Age 15-19 | 2001 | PM | 23 | 4.433 | 99.9 | 9.443 [3.786, 23.549] |
| Estimated rate of annual AIDS-related deaths (per 100,000 population) | ESA | Age 15-19 | 2001 | DL | 23 | 0.463 | 99.4 | 10.866 [4.594, 25.701] |
| Estimated rate of annual AIDS-related deaths (per 100,000 population) | ESA | Age 15-19 | 2002 | REML | 23 | 4.087 | 99.9 | 10.724 [4.433, 25.942] |
| Estimated rate of annual AIDS-related deaths (per 100,000 population) | ESA | Age 15-19 | 2002 | PM | 23 | 4.145 | 99.9 | 10.722 [4.432, 25.938] |
| Estimated rate of annual AIDS-related deaths (per 100,000 population) | ESA | Age 15-19 | 2002 | DL | 23 | 0.447 | 99.4 | 12.050 [5.192, 27.967] |
| Estimated rate of annual AIDS-related deaths (per 100,000 population) | ESA | Age 15-19 | 2003 | REML | 23 | 4.016 | 99.9 | 11.963 [4.990, 28.679] |
| Estimated rate of annual AIDS-related deaths (per 100,000 population) | ESA | Age 15-19 | 2003 | PM | 23 | 4.063 | 99.9 | 11.961 [4.989, 28.675] |
| Estimated rate of annual AIDS-related deaths (per 100,000 population) | ESA | Age 15-19 | 2003 | DL | 23 | 0.443 | 99.4 | 13.233 [5.716, 30.636] |

*Continued on next p*

**Supplementary Table S4 (continued)**

| Indicator | Region | Age group | Year | Method | $k$ | $\tau^2$ | $I^2$ (%) | Pooled estimate [95% |
|---|---|---|---|---|---|---|---|---|
| Estimated rate of annual AIDS-related deaths (per 100,000 population) | ESA | Age 15-19 | 2004 | REML | 23 | 3.903 | 99.9 | 13.231 [5.590, 31.319] |
| Estimated rate of annual AIDS-related deaths (per 100,000 population) | ESA | Age 15-19 | 2004 | PM | 23 | 3.947 | 99.9 | 13.229 [5.588, 31.316] |
| Estimated rate of annual AIDS-related deaths (per 100,000 population) | ESA | Age 15-19 | 2004 | DL | 23 | 0.481 | 99.5 | 14.392 [6.261, 33.082] |
| Estimated rate of annual AIDS-related deaths (per 100,000 population) | ESA | Age 15-19 | 2005 | REML | 23 | 3.732 | 99.9 | 14.285 [6.153, 33.166] |
| Estimated rate of annual AIDS-related deaths (per 100,000 population) | ESA | Age 15-19 | 2005 | PM | 23 | 3.772 | 99.9 | 14.283 [6.151, 33.164] |
| Estimated rate of annual AIDS-related deaths (per 100,000 population) | ESA | Age 15-19 | 2005 | DL | 23 | 0.580 | 99.5 | 15.184 [6.683, 34.501] |
| Estimated rate of annual AIDS-related deaths (per 100,000 population) | ESA | Age 15-19 | 2006 | REML | 23 | 3.585 | 99.9 | 14.905 [6.534, 34.000] |
| Estimated rate of annual AIDS-related deaths (per 100,000 population) | ESA | Age 15-19 | 2006 | PM | 23 | 3.616 | 99.9 | 14.904 [6.534, 33.998] |
| Estimated rate of annual AIDS-related deaths (per 100,000 population) | ESA | Age 15-19 | 2006 | DL | 23 | 0.724 | 99.5 | 15.486 [6.878, 34.870] |
| Estimated rate of annual AIDS-related deaths (per 100,000 population) | ESA | Age 15-19 | 2007 | REML | 23 | 3.440 | 99.9 | 14.909 [6.646, 33.444] |
| Estimated rate of annual AIDS-related deaths (per 100,000 population) | ESA | Age 15-19 | 2007 | PM | 23 | 3.470 | 99.9 | 14.908 [6.645, 33.442] |
| Estimated rate of annual AIDS-related deaths (per 100,000 population) | ESA | Age 15-19 | 2007 | DL | 23 | 0.867 | 99.4 | 15.336 [6.901, 34.078] |
| Estimated rate of annual AIDS-related deaths (per 100,000 population) | ESA | Age 15-19 | 2008 | REML | 23 | 3.245 | 99.8 | 14.854 [6.779, 32.544] |
| Estimated rate of annual AIDS-related deaths (per 100,000 population) | ESA | Age 15-19 | 2008 | PM | 23 | 3.269 | 99.8 | 14.853 [6.779, 32.543] |
| Estimated rate of annual AIDS-related deaths (per 100,000 population) | ESA | Age 15-19 | 2008 | DL | 23 | 1.087 | 99.4 | 15.112 [6.936, 32.927] |
| Estimated rate of annual AIDS-related deaths (per 100,000 population) | ESA | Age 15-19 | 2009 | REML | 23 | 3.146 | 99.7 | 15.009 [6.935, 32.487] |
| Estimated rate of annual AIDS-related deaths (per 100,000 population) | ESA | Age 15-19 | 2009 | PM | 23 | 3.167 | 99.7 | 15.009 [6.934, 32.486] |
| Estimated rate of annual AIDS-related deaths (per 100,000 population) | ESA | Age 15-19 | 2009 | DL | 23 | 1.238 | 99.3 | 15.201 [7.049, 32.778] |
| Estimated rate of annual AIDS-related deaths (per 100,000 population) | ESA | Age 15-19 | 2010 | REML | 23 | 3.197 | 99.7 | 15.032 [6.892, 32.784] |
| Estimated rate of annual AIDS-related deaths (per 100,000 population) | ESA | Age 15-19 | 2010 | PM | 23 | 3.228 | 99.7 | 15.030 [6.891, 32.782] |
| Estimated rate of annual AIDS-related deaths (per 100,000 population) | ESA | Age 15-19 | 2010 | DL | 23 | 1.267 | 99.3 | 15.275 [7.042, 33.135] |
| Estimated rate of annual AIDS-related deaths (per 100,000 population) | ESA | Age 15-19 | 2011 | REML | 23 | 3.193 | 99.7 | 15.373 [7.034, 33.595] |
| Estimated rate of annual AIDS-related deaths (per 100,000 population) | ESA | Age 15-19 | 2011 | PM | 23 | 3.241 | 99.7 | 15.369 [7.032, 33.590] |
| Estimated rate of annual AIDS-related deaths (per 100,000 population) | ESA | Age 15-19 | 2011 | DL | 23 | 1.374 | 99.2 | 15.666 [7.220, 33.994] |
| Estimated rate of annual AIDS-related deaths (per 100,000 population) | ESA | Age 15-19 | 2012 | REML | 23 | 3.180 | 99.7 | 15.578 [7.139, 33.995] |
| Estimated rate of annual AIDS-related deaths (per 100,000 population) | ESA | Age 15-19 | 2012 | PM | 23 | 3.229 | 99.7 | 15.574 [7.136, 33.990] |
| Estimated rate of annual AIDS-related deaths (per 100,000 population) | ESA | Age 15-19 | 2012 | DL | 23 | 1.538 | 99.3 | 15.824 [7.294, 34.332] |
| Estimated rate of annual AIDS-related deaths (per 100,000 population) | ESA | Age 15-19 | 2013 | REML | 23 | 3.132 | 99.6 | 15.565 [7.163, 33.822] |
| Estimated rate of annual AIDS-related deaths (per 100,000 population) | ESA | Age 15-19 | 2013 | PM | 23 | 3.191 | 99.6 | 15.559 [7.159, 33.815] |
| Estimated rate of annual AIDS-related deaths (per 100,000 population) | ESA | Age 15-19 | 2013 | DL | 23 | 1.723 | 99.3 | 15.789 [7.305, 34.127] |

*Continued on next*

**Supplementary Table S4 (continued)**

| Indicator | Region | Age group | Year | Method | $k$ | $\tau^2$ | $I^2$ (%) | Pooled estimate [95% |
|---|---|---|---|---|---|---|---|---|
| Estimated rate of annual AIDS-related deaths (per 100,000 population) | ESA | Age 15-19 | 2014 | REML | 23 | 3.082 | 99.6 | 15.280 [7.080, 32.978] |
| Estimated rate of annual AIDS-related deaths (per 100,000 population) | ESA | Age 15-19 | 2014 | PM | 23 | 3.134 | 99.6 | 15.276 [7.077, 32.972] |
| Estimated rate of annual AIDS-related deaths (per 100,000 population) | ESA | Age 15-19 | 2014 | DL | 23 | 1.832 | 99.4 | 15.458 [7.192, 33.226] |
| Estimated rate of annual AIDS-related deaths (per 100,000 population) | ESA | Age 15-19 | 2015 | REML | 23 | 3.112 | 99.6 | 14.937 [6.889, 32.389] |
| Estimated rate of annual AIDS-related deaths (per 100,000 population) | ESA | Age 15-19 | 2015 | PM | 23 | 3.171 | 99.6 | 14.932 [6.885, 32.382] |
| Estimated rate of annual AIDS-related deaths (per 100,000 population) | ESA | Age 15-19 | 2015 | DL | 23 | 1.858 | 99.4 | 15.128 [7.008, 32.654] |
| Estimated rate of annual AIDS-related deaths (per 100,000 population) | ESA | Age 15-19 | 2016 | REML | 23 | 3.145 | 99.6 | 14.587 [6.700, 31.758] |
| Estimated rate of annual AIDS-related deaths (per 100,000 population) | ESA | Age 15-19 | 2016 | PM | 23 | 3.204 | 99.6 | 14.581 [6.696, 31.751] |
| Estimated rate of annual AIDS-related deaths (per 100,000 population) | ESA | Age 15-19 | 2016 | DL | 23 | 1.910 | 99.4 | 14.768 [6.813, 32.015] |
| Estimated rate of annual AIDS-related deaths (per 100,000 population) | ESA | Age 15-19 | 2017 | REML | 23 | 3.129 | 99.6 | 14.041 [6.467, 30.485] |
| Estimated rate of annual AIDS-related deaths (per 100,000 population) | ESA | Age 15-19 | 2017 | PM | 23 | 3.182 | 99.6 | 14.036 [6.464, 30.479] |
| Estimated rate of annual AIDS-related deaths (per 100,000 population) | ESA | Age 15-19 | 2017 | DL | 23 | 1.881 | 99.4 | 14.212 [6.572, 30.735] |
| Estimated rate of annual AIDS-related deaths (per 100,000 population) | ESA | Age 15-19 | 2018 | REML | 23 | 3.112 | 99.6 | 13.212 [6.095, 28.637] |
| Estimated rate of annual AIDS-related deaths (per 100,000 population) | ESA | Age 15-19 | 2018 | PM | 23 | 3.167 | 99.6 | 13.207 [6.092, 28.630] |
| Estimated rate of annual AIDS-related deaths (per 100,000 population) | ESA | Age 15-19 | 2018 | DL | 23 | 1.802 | 99.3 | 13.397 [6.209, 28.906] |
| Estimated rate of annual AIDS-related deaths (per 100,000 population) | ESA | Age 15-19 | 2019 | REML | 23 | 3.162 | 99.6 | 12.623 [5.783, 27.555] |
| Estimated rate of annual AIDS-related deaths (per 100,000 population) | ESA | Age 15-19 | 2019 | PM | 23 | 3.224 | 99.6 | 12.618 [5.780, 27.548] |
| Estimated rate of annual AIDS-related deaths (per 100,000 population) | ESA | Age 15-19 | 2019 | DL | 23 | 1.744 | 99.3 | 12.834 [5.913, 27.857] |
| Estimated rate of annual AIDS-related deaths (per 100,000 population) | ESA | Age 15-19 | 2020 | REML | 23 | 3.187 | 99.6 | 11.924 [5.441, 26.132] |
| Estimated rate of annual AIDS-related deaths (per 100,000 population) | ESA | Age 15-19 | 2020 | PM | 23 | 3.256 | 99.6 | 11.918 [5.438, 26.124] |
| Estimated rate of annual AIDS-related deaths (per 100,000 population) | ESA | Age 15-19 | 2020 | DL | 23 | 1.702 | 99.2 | 12.150 [5.581, 26.452] |
| Estimated rate of annual AIDS-related deaths (per 100,000 population) | ESA | Age 15-19 | 2021 | REML | 23 | 3.169 | 99.5 | 11.093 [5.068, 24.279] |
| Estimated rate of annual AIDS-related deaths (per 100,000 population) | ESA | Age 15-19 | 2021 | PM | 23 | 3.243 | 99.5 | 11.086 [5.064, 24.270] |
| Estimated rate of annual AIDS-related deaths (per 100,000 population) | ESA | Age 15-19 | 2021 | DL | 23 | 1.661 | 99.1 | 11.326 [5.213, 24.610] |
| Estimated rate of annual AIDS-related deaths (per 100,000 population) | ESA | Age 15-19 | 2022 | REML | 23 | 3.126 | 99.5 | 10.182 [4.675, 22.179] |
| Estimated rate of annual AIDS-related deaths (per 100,000 population) | ESA | Age 15-19 | 2022 | PM | 23 | 3.201 | 99.5 | 10.176 [4.671, 22.170] |
| Estimated rate of annual AIDS-related deaths (per 100,000 population) | ESA | Age 15-19 | 2022 | DL | 23 | 1.543 | 99.0 | 10.432 [4.830, 22.534] |
| Estimated rate of annual AIDS-related deaths (per 100,000 population) | ESA | Age 15-19 | 2023 | REML | 22 | 2.898 | 99.4 | 10.455 [4.835, 22.610] |
| Estimated rate of annual AIDS-related deaths (per 100,000 population) | ESA | Age 15-19 | 2023 | PM | 22 | 2.986 | 99.5 | 10.447 [4.829, 22.598] |
| Estimated rate of annual AIDS-related deaths (per 100,000 population) | ESA | Age 15-19 | 2023 | DL | 22 | 1.254 | 98.7 | 10.811 [5.064, 23.077] |

*Continued on next*

**Supplementary Table S4 (continued)**

| Indicator | Region | Age group | Year | Method | $k$ | $\tau^2$ | $I^2$ (%) | Pooled estimate [95% |
|---|---|---|---|---|---|---|---|---|
| Estimated rate of annual AIDS-related deaths (per 100,000 population) | ESA | Age 15-19 | 2024 | REML | 22 | 2.806 | 99.5 | 9.580 [4.474, 20.512] |
| Estimated rate of annual AIDS-related deaths (per 100,000 population) | ESA | Age 15-19 | 2024 | PM | 22 | 2.906 | 99.5 | 9.570 [4.468, 20.499] |
| Estimated rate of annual AIDS-related deaths (per 100,000 population) | ESA | Age 15-19 | 2024 | DL | 22 | 1.222 | 98.8 | 9.936 [4.708, 20.970] |
| Estimated rate of annual AIDS-related deaths (per 100,000 population) | WCA | Age 0-14 | 2000 | REML | 21 | 0.622 | 98.8 | 49.970 [34.797, 71.757] |
| Estimated rate of annual AIDS-related deaths (per 100,000 population) | WCA | Age 0-14 | 2000 | PM | 21 | 0.620 | 98.8 | 49.970 [34.797, 71.757] |
| Estimated rate of annual AIDS-related deaths (per 100,000 population) | WCA | Age 0-14 | 2000 | DL | 21 | 0.959 | 99.2 | 49.987 [34.817, 71.767] |
| Estimated rate of annual AIDS-related deaths (per 100,000 population) | WCA | Age 0-14 | 2001 | REML | 21 | 0.612 | 98.9 | 50.325 [35.159, 72.034] |
| Estimated rate of annual AIDS-related deaths (per 100,000 population) | WCA | Age 0-14 | 2001 | PM | 21 | 0.610 | 98.9 | 50.325 [35.158, 72.034] |
| Estimated rate of annual AIDS-related deaths (per 100,000 population) | WCA | Age 0-14 | 2001 | DL | 21 | 0.893 | 99.2 | 50.342 [35.176, 72.046] |
| Estimated rate of annual AIDS-related deaths (per 100,000 population) | WCA | Age 0-14 | 2002 | REML | 21 | 0.604 | 98.8 | 50.159 [35.127, 71.624] |
| Estimated rate of annual AIDS-related deaths (per 100,000 population) | WCA | Age 0-14 | 2002 | PM | 21 | 0.602 | 98.8 | 50.159 [35.127, 71.624] |
| Estimated rate of annual AIDS-related deaths (per 100,000 population) | WCA | Age 0-14 | 2002 | DL | 21 | 0.837 | 99.2 | 50.174 [35.142, 71.635] |
| Estimated rate of annual AIDS-related deaths (per 100,000 population) | WCA | Age 0-14 | 2003 | REML | 21 | 0.600 | 98.9 | 49.423 [34.655, 70.483] |
| Estimated rate of annual AIDS-related deaths (per 100,000 population) | WCA | Age 0-14 | 2003 | PM | 21 | 0.599 | 98.9 | 49.423 [34.655, 70.483] |
| Estimated rate of annual AIDS-related deaths (per 100,000 population) | WCA | Age 0-14 | 2003 | DL | 21 | 0.812 | 99.2 | 49.436 [34.669, 70.495] |
| Estimated rate of annual AIDS-related deaths (per 100,000 population) | WCA | Age 0-14 | 2004 | REML | 21 | 0.594 | 99.0 | 48.201 [33.864, 68.608] |
| Estimated rate of annual AIDS-related deaths (per 100,000 population) | WCA | Age 0-14 | 2004 | PM | 21 | 0.593 | 99.0 | 48.201 [33.864, 68.608] |
| Estimated rate of annual AIDS-related deaths (per 100,000 population) | WCA | Age 0-14 | 2004 | DL | 21 | 0.813 | 99.2 | 48.213 [33.876, 68.617] |
| Estimated rate of annual AIDS-related deaths (per 100,000 population) | WCA | Age 0-14 | 2005 | REML | 21 | 0.594 | 99.0 | 46.611 [32.753, 66.333] |
| Estimated rate of annual AIDS-related deaths (per 100,000 population) | WCA | Age 0-14 | 2005 | PM | 21 | 0.593 | 99.0 | 46.611 [32.753, 66.333] |
| Estimated rate of annual AIDS-related deaths (per 100,000 population) | WCA | Age 0-14 | 2005 | DL | 21 | 0.789 | 99.2 | 46.623 [32.765, 66.342] |
| Estimated rate of annual AIDS-related deaths (per 100,000 population) | WCA | Age 0-14 | 2006 | REML | 21 | 0.598 | 99.0 | 44.683 [31.364, 63.657] |
| Estimated rate of annual AIDS-related deaths (per 100,000 population) | WCA | Age 0-14 | 2006 | PM | 21 | 0.597 | 99.0 | 44.683 [31.364, 63.657] |
| Estimated rate of annual AIDS-related deaths (per 100,000 population) | WCA | Age 0-14 | 2006 | DL | 21 | 0.814 | 99.3 | 44.695 [31.376, 63.668] |
| Estimated rate of annual AIDS-related deaths (per 100,000 population) | WCA | Age 0-14 | 2007 | REML | 21 | 0.603 | 99.0 | 42.384 [29.712, 60.459] |
| Estimated rate of annual AIDS-related deaths (per 100,000 population) | WCA | Age 0-14 | 2007 | PM | 21 | 0.601 | 99.0 | 42.383 [29.712, 60.459] |
| Estimated rate of annual AIDS-related deaths (per 100,000 population) | WCA | Age 0-14 | 2007 | DL | 21 | 0.843 | 99.3 | 42.400 [29.727, 60.474] |
| Estimated rate of annual AIDS-related deaths (per 100,000 population) | WCA | Age 0-14 | 2008 | REML | 21 | 0.602 | 99.0 | 39.760 [27.872, 56.719] |
| Estimated rate of annual AIDS-related deaths (per 100,000 population) | WCA | Age 0-14 | 2008 | PM | 21 | 0.601 | 99.0 | 39.760 [27.872, 56.719] |
| Estimated rate of annual AIDS-related deaths (per 100,000 population) | WCA | Age 0-14 | 2008 | DL | 21 | 0.863 | 99.3 | 39.782 [27.891, 56.741] |

*Continued on next p*

**Supplementary Table S4 (continued)**

| Indicator | Region | Age group | Year | Method | $k$ | $\tau^2$ | $I^2$ (%) | Pooled estimate [95% |
|---|---|---|---|---|---|---|---|---|
| Estimated rate of annual AIDS-related deaths (per 100,000 population) | WCA | Age 0-14 | 2009 | REML | 21 | 0.625 | 99.0 | 36.905 [25.705, 52.987] |
| Estimated rate of annual AIDS-related deaths (per 100,000 population) | WCA | Age 0-14 | 2009 | PM | 21 | 0.623 | 99.0 | 36.905 [25.704, 52.987] |
| Estimated rate of annual AIDS-related deaths (per 100,000 population) | WCA | Age 0-14 | 2009 | DL | 21 | 0.936 | 99.3 | 36.924 [25.723, 53.004] |
| Estimated rate of annual AIDS-related deaths (per 100,000 population) | WCA | Age 0-14 | 2010 | REML | 21 | 0.664 | 99.0 | 33.259 [22.903, 48.298] |
| Estimated rate of annual AIDS-related deaths (per 100,000 population) | WCA | Age 0-14 | 2010 | PM | 21 | 0.662 | 98.9 | 33.259 [22.903, 48.298] |
| Estimated rate of annual AIDS-related deaths (per 100,000 population) | WCA | Age 0-14 | 2010 | DL | 21 | 0.964 | 99.3 | 33.271 [22.916, 48.306] |
| Estimated rate of annual AIDS-related deaths (per 100,000 population) | WCA | Age 0-14 | 2011 | REML | 21 | 0.691 | 98.9 | 29.753 [20.332, 43.540] |
| Estimated rate of annual AIDS-related deaths (per 100,000 population) | WCA | Age 0-14 | 2011 | PM | 21 | 0.688 | 98.8 | 29.753 [20.332, 43.540] |
| Estimated rate of annual AIDS-related deaths (per 100,000 population) | WCA | Age 0-14 | 2011 | DL | 21 | 0.953 | 99.2 | 29.760 [20.341, 43.543] |
| Estimated rate of annual AIDS-related deaths (per 100,000 population) | WCA | Age 0-14 | 2012 | REML | 21 | 0.715 | 98.8 | 26.897 [18.254, 39.632] |
| Estimated rate of annual AIDS-related deaths (per 100,000 population) | WCA | Age 0-14 | 2012 | PM | 21 | 0.713 | 98.8 | 26.897 [18.253, 39.632] |
| Estimated rate of annual AIDS-related deaths (per 100,000 population) | WCA | Age 0-14 | 2012 | DL | 21 | 0.968 | 99.1 | 26.902 [18.260, 39.634] |
| Estimated rate of annual AIDS-related deaths (per 100,000 population) | WCA | Age 0-14 | 2013 | REML | 21 | 0.764 | 98.7 | 23.662 [15.848, 35.327] |
| Estimated rate of annual AIDS-related deaths (per 100,000 population) | WCA | Age 0-14 | 2013 | PM | 21 | 0.761 | 98.7 | 23.662 [15.848, 35.327] |
| Estimated rate of annual AIDS-related deaths (per 100,000 population) | WCA | Age 0-14 | 2013 | DL | 21 | 1.021 | 99.1 | 23.667 [15.855, 35.329] |
| Estimated rate of annual AIDS-related deaths (per 100,000 population) | WCA | Age 0-14 | 2014 | REML | 21 | 0.776 | 98.7 | 21.317 [14.231, 31.931] |
| Estimated rate of annual AIDS-related deaths (per 100,000 population) | WCA | Age 0-14 | 2014 | PM | 21 | 0.773 | 98.7 | 21.317 [14.230, 31.931] |
| Estimated rate of annual AIDS-related deaths (per 100,000 population) | WCA | Age 0-14 | 2014 | DL | 21 | 1.005 | 99.0 | 21.322 [14.237, 31.935] |
| Estimated rate of annual AIDS-related deaths (per 100,000 population) | WCA | Age 0-14 | 2015 | REML | 21 | 0.770 | 98.5 | 19.050 [12.733, 28.500] |
| Estimated rate of annual AIDS-related deaths (per 100,000 population) | WCA | Age 0-14 | 2015 | PM | 21 | 0.767 | 98.5 | 19.050 [12.733, 28.500] |
| Estimated rate of annual AIDS-related deaths (per 100,000 population) | WCA | Age 0-14 | 2015 | DL | 21 | 1.007 | 98.9 | 19.058 [12.741, 28.507] |
| Estimated rate of annual AIDS-related deaths (per 100,000 population) | WCA | Age 0-14 | 2016 | REML | 21 | 0.805 | 98.5 | 17.204 [11.397, 25.971] |
| Estimated rate of annual AIDS-related deaths (per 100,000 population) | WCA | Age 0-14 | 2016 | PM | 21 | 0.801 | 98.5 | 17.204 [11.397, 25.971] |
| Estimated rate of annual AIDS-related deaths (per 100,000 population) | WCA | Age 0-14 | 2016 | DL | 21 | 1.129 | 98.9 | 17.216 [11.408, 25.981] |
| Estimated rate of annual AIDS-related deaths (per 100,000 population) | WCA | Age 0-14 | 2017 | REML | 21 | 0.821 | 98.3 | 16.093 [10.615, 24.398] |
| Estimated rate of annual AIDS-related deaths (per 100,000 population) | WCA | Age 0-14 | 2017 | PM | 21 | 0.816 | 98.3 | 16.093 [10.615, 24.397] |
| Estimated rate of annual AIDS-related deaths (per 100,000 population) | WCA | Age 0-14 | 2017 | DL | 21 | 1.153 | 98.8 | 16.106 [10.627, 24.410] |
| Estimated rate of annual AIDS-related deaths (per 100,000 population) | WCA | Age 0-14 | 2018 | REML | 21 | 0.880 | 98.3 | 14.695 [9.555, 22.601] |
| Estimated rate of annual AIDS-related deaths (per 100,000 population) | WCA | Age 0-14 | 2018 | PM | 21 | 0.874 | 98.3 | 14.695 [9.555, 22.601] |
| Estimated rate of annual AIDS-related deaths (per 100,000 population) | WCA | Age 0-14 | 2018 | DL | 21 | 1.285 | 98.8 | 14.716 [9.573, 22.622] |

*Continued on next p*

**Supplementary Table S4 (continued)**

| Indicator | Region | Age group | Year | Method | $k$ | $\tau^2$ | $I^2$ (%) | Pooled estimate [95% |
|---|---|---|---|---|---|---|---|---|
| Estimated rate of annual AIDS-related deaths (per 100,000 population) | WCA | Age 0-14 | 2019 | REML | 21 | 0.917 | 98.1 | 13.408 [8.638, 20.812] |
| Estimated rate of annual AIDS-related deaths (per 100,000 population) | WCA | Age 0-14 | 2019 | PM | 21 | 0.909 | 98.1 | 13.407 [8.637, 20.811] |
| Estimated rate of annual AIDS-related deaths (per 100,000 population) | WCA | Age 0-14 | 2019 | DL | 21 | 1.392 | 98.8 | 13.430 [8.657, 20.833] |
| Estimated rate of annual AIDS-related deaths (per 100,000 population) | WCA | Age 0-14 | 2020 | REML | 21 | 0.935 | 97.7 | 12.314 [7.892, 19.215] |
| Estimated rate of annual AIDS-related deaths (per 100,000 population) | WCA | Age 0-14 | 2020 | PM | 21 | 0.927 | 97.7 | 12.314 [7.891, 19.214] |
| Estimated rate of annual AIDS-related deaths (per 100,000 population) | WCA | Age 0-14 | 2020 | DL | 21 | 1.412 | 98.5 | 12.331 [7.908, 19.230] |
| Estimated rate of annual AIDS-related deaths (per 100,000 population) | WCA | Age 0-14 | 2021 | REML | 21 | 0.908 | 97.3 | 11.134 [7.167, 17.297] |
| Estimated rate of annual AIDS-related deaths (per 100,000 population) | WCA | Age 0-14 | 2021 | PM | 21 | 0.903 | 97.2 | 11.134 [7.167, 17.296] |
| Estimated rate of annual AIDS-related deaths (per 100,000 population) | WCA | Age 0-14 | 2021 | DL | 21 | 1.311 | 98.1 | 11.144 [7.177, 17.306] |
| Estimated rate of annual AIDS-related deaths (per 100,000 population) | WCA | Age 0-14 | 2022 | REML | 21 | 0.876 | 96.9 | 10.461 [6.778, 16.144] |
| Estimated rate of annual AIDS-related deaths (per 100,000 population) | WCA | Age 0-14 | 2022 | PM | 21 | 0.870 | 96.9 | 10.460 [6.778, 16.144] |
| Estimated rate of annual AIDS-related deaths (per 100,000 population) | WCA | Age 0-14 | 2022 | DL | 21 | 1.283 | 97.9 | 10.469 [6.786, 16.151] |
| Estimated rate of annual AIDS-related deaths (per 100,000 population) | WCA | Age 0-14 | 2023 | REML | 21 | 0.931 | 96.7 | 9.602 [6.134, 15.031] |
| Estimated rate of annual AIDS-related deaths (per 100,000 population) | WCA | Age 0-14 | 2023 | PM | 21 | 0.926 | 96.7 | 9.602 [6.134, 15.031] |
| Estimated rate of annual AIDS-related deaths (per 100,000 population) | WCA | Age 0-14 | 2023 | DL | 21 | 1.284 | 97.6 | 9.612 [6.143, 15.042] |
| Estimated rate of annual AIDS-related deaths (per 100,000 population) | WCA | Age 0-14 | 2024 | REML | 21 | 0.825 | 95.9 | 8.816 [5.762, 13.488] |
| Estimated rate of annual AIDS-related deaths (per 100,000 population) | WCA | Age 0-14 | 2024 | PM | 21 | 0.820 | 95.9 | 8.816 [5.762, 13.488] |
| Estimated rate of annual AIDS-related deaths (per 100,000 population) | WCA | Age 0-14 | 2024 | DL | 21 | 1.163 | 97.1 | 8.828 [5.772, 13.504] |
| Estimated rate of annual AIDS-related deaths (per 100,000 population) | WCA | Age 15-19 | 2000 | REML | 22 | 1.135 | 99.3 | 4.663 [2.898, 7.503] |
| Estimated rate of annual AIDS-related deaths (per 100,000 population) | WCA | Age 15-19 | 2000 | PM | 22 | 1.134 | 99.3 | 4.663 [2.898, 7.503] |
| Estimated rate of annual AIDS-related deaths (per 100,000 population) | WCA | Age 15-19 | 2000 | DL | 22 | 2.018 | 99.6 | 4.665 [2.900, 7.505] |
| Estimated rate of annual AIDS-related deaths (per 100,000 population) | WCA | Age 15-19 | 2001 | REML | 22 | 1.114 | 99.4 | 5.214 [3.257, 8.348] |
| Estimated rate of annual AIDS-related deaths (per 100,000 population) | WCA | Age 15-19 | 2001 | PM | 22 | 1.112 | 99.4 | 5.214 [3.257, 8.348] |
| Estimated rate of annual AIDS-related deaths (per 100,000 population) | WCA | Age 15-19 | 2001 | DL | 22 | 1.952 | 99.6 | 5.216 [3.258, 8.349] |
| Estimated rate of annual AIDS-related deaths (per 100,000 population) | WCA | Age 15-19 | 2002 | REML | 22 | 1.105 | 99.4 | 5.814 [3.640, 9.288] |
| Estimated rate of annual AIDS-related deaths (per 100,000 population) | WCA | Age 15-19 | 2002 | PM | 22 | 1.103 | 99.4 | 5.814 [3.640, 9.288] |
| Estimated rate of annual AIDS-related deaths (per 100,000 population) | WCA | Age 15-19 | 2002 | DL | 22 | 2.010 | 99.7 | 5.815 [3.641, 9.288] |
| Estimated rate of annual AIDS-related deaths (per 100,000 population) | WCA | Age 15-19 | 2003 | REML | 22 | 1.086 | 99.5 | 6.512 [4.094, 10.358] |
| Estimated rate of annual AIDS-related deaths (per 100,000 population) | WCA | Age 15-19 | 2003 | PM | 22 | 1.084 | 99.5 | 6.512 [4.094, 10.358] |
| Estimated rate of annual AIDS-related deaths (per 100,000 population) | WCA | Age 15-19 | 2003 | DL | 22 | 1.973 | 99.7 | 6.512 [4.095, 10.357] |

*Continued on next p*

**Supplementary Table S4 (continued)**

| Indicator | Region | Age group | Year | Method | $k$ | $\tau^2$ | $I^2$ (%) | Pooled estimate [95% |
|---|---|---|---|---|---|---|---|---|
| Estimated rate of annual AIDS-related deaths (per 100,000 population) | WCA | Age 15-19 | 2004 | REML | 22 | 1.053 | 99.5 | 7.223 [4.574, 11.406] |
| Estimated rate of annual AIDS-related deaths (per 100,000 population) | WCA | Age 15-19 | 2004 | PM | 22 | 1.051 | 99.5 | 7.223 [4.574, 11.406] |
| Estimated rate of annual AIDS-related deaths (per 100,000 population) | WCA | Age 15-19 | 2004 | DL | 22 | 1.989 | 99.7 | 7.224 [4.576, 11.406] |
| Estimated rate of annual AIDS-related deaths (per 100,000 population) | WCA | Age 15-19 | 2005 | REML | 22 | 1.015 | 99.5 | 7.813 [4.988, 12.237] |
| Estimated rate of annual AIDS-related deaths (per 100,000 population) | WCA | Age 15-19 | 2005 | PM | 22 | 1.013 | 99.5 | 7.813 [4.988, 12.237] |
| Estimated rate of annual AIDS-related deaths (per 100,000 population) | WCA | Age 15-19 | 2005 | DL | 22 | 1.931 | 99.7 | 7.814 [4.990, 12.235] |
| Estimated rate of annual AIDS-related deaths (per 100,000 population) | WCA | Age 15-19 | 2006 | REML | 22 | 0.981 | 99.5 | 8.390 [5.396, 13.043] |
| Estimated rate of annual AIDS-related deaths (per 100,000 population) | WCA | Age 15-19 | 2006 | PM | 22 | 0.978 | 99.5 | 8.390 [5.396, 13.043] |
| Estimated rate of annual AIDS-related deaths (per 100,000 population) | WCA | Age 15-19 | 2006 | DL | 22 | 1.809 | 99.7 | 8.388 [5.397, 13.038] |
| Estimated rate of annual AIDS-related deaths (per 100,000 population) | WCA | Age 15-19 | 2007 | REML | 22 | 0.994 | 99.3 | 8.971 [5.754, 13.988] |
| Estimated rate of annual AIDS-related deaths (per 100,000 population) | WCA | Age 15-19 | 2007 | PM | 22 | 0.991 | 99.3 | 8.971 [5.754, 13.988] |
| Estimated rate of annual AIDS-related deaths (per 100,000 population) | WCA | Age 15-19 | 2007 | DL | 22 | 1.698 | 99.6 | 8.971 [5.755, 13.984] |
| Estimated rate of annual AIDS-related deaths (per 100,000 population) | WCA | Age 15-19 | 2008 | REML | 22 | 0.988 | 99.1 | 9.249 [5.939, 14.406] |
| Estimated rate of annual AIDS-related deaths (per 100,000 population) | WCA | Age 15-19 | 2008 | PM | 22 | 0.985 | 99.1 | 9.249 [5.939, 14.406] |
| Estimated rate of annual AIDS-related deaths (per 100,000 population) | WCA | Age 15-19 | 2008 | DL | 22 | 1.339 | 99.4 | 9.249 [5.939, 14.403] |
| Estimated rate of annual AIDS-related deaths (per 100,000 population) | WCA | Age 15-19 | 2009 | REML | 22 | 0.929 | 98.9 | 9.511 [6.185, 14.625] |
| Estimated rate of annual AIDS-related deaths (per 100,000 population) | WCA | Age 15-19 | 2009 | PM | 22 | 0.928 | 98.9 | 9.511 [6.185, 14.625] |
| Estimated rate of annual AIDS-related deaths (per 100,000 population) | WCA | Age 15-19 | 2009 | DL | 22 | 1.123 | 99.1 | 9.510 [6.184, 14.623] |
| Estimated rate of annual AIDS-related deaths (per 100,000 population) | WCA | Age 15-19 | 2010 | REML | 22 | 0.861 | 98.7 | 10.053 [6.641, 15.220] |
| Estimated rate of annual AIDS-related deaths (per 100,000 population) | WCA | Age 15-19 | 2010 | PM | 22 | 0.859 | 98.7 | 10.054 [6.641, 15.220] |
| Estimated rate of annual AIDS-related deaths (per 100,000 population) | WCA | Age 15-19 | 2010 | DL | 22 | 1.044 | 98.9 | 10.051 [6.640, 15.216] |
| Estimated rate of annual AIDS-related deaths (per 100,000 population) | WCA | Age 15-19 | 2011 | REML | 22 | 0.799 | 98.6 | 10.587 [7.097, 15.794] |
| Estimated rate of annual AIDS-related deaths (per 100,000 population) | WCA | Age 15-19 | 2011 | PM | 22 | 0.798 | 98.6 | 10.587 [7.097, 15.794] |
| Estimated rate of annual AIDS-related deaths (per 100,000 population) | WCA | Age 15-19 | 2011 | DL | 22 | 0.952 | 98.8 | 10.585 [7.095, 15.789] |
| Estimated rate of annual AIDS-related deaths (per 100,000 population) | WCA | Age 15-19 | 2012 | REML | 22 | 0.758 | 98.5 | 11.120 [7.530, 16.422] |
| Estimated rate of annual AIDS-related deaths (per 100,000 population) | WCA | Age 15-19 | 2012 | PM | 22 | 0.757 | 98.5 | 11.120 [7.530, 16.422] |
| Estimated rate of annual AIDS-related deaths (per 100,000 population) | WCA | Age 15-19 | 2012 | DL | 22 | 0.803 | 98.6 | 11.119 [7.529, 16.420] |
| Estimated rate of annual AIDS-related deaths (per 100,000 population) | WCA | Age 15-19 | 2013 | REML | 22 | 0.737 | 98.4 | 11.519 [7.843, 16.918] |
| Estimated rate of annual AIDS-related deaths (per 100,000 population) | WCA | Age 15-19 | 2013 | PM | 22 | 0.735 | 98.4 | 11.519 [7.843, 16.918] |
| Estimated rate of annual AIDS-related deaths (per 100,000 population) | WCA | Age 15-19 | 2013 | DL | 22 | 0.772 | 98.5 | 11.518 [7.843, 16.916] |

*Continued on next p*

**Supplementary Table S4 (continued)**

| Indicator | Region | Age group | Year | Method | $k$ | $\tau^2$ | $I^2$ (%) | Pooled estimate [95% |
|---|---|---|---|---|---|---|---|---|
| Estimated rate of annual AIDS-related deaths (per 100,000 population) | WCA | Age 15-19 | 2014 | REML | 22 | 0.706 | 98.4 | 11.639 [7.991, 16.954] |
| Estimated rate of annual AIDS-related deaths (per 100,000 population) | WCA | Age 15-19 | 2014 | PM | 22 | 0.704 | 98.4 | 11.639 [7.991, 16.955] |
| Estimated rate of annual AIDS-related deaths (per 100,000 population) | WCA | Age 15-19 | 2014 | DL | 22 | 0.732 | 98.4 | 11.639 [7.990, 16.953] |
| Estimated rate of annual AIDS-related deaths (per 100,000 population) | WCA | Age 15-19 | 2015 | REML | 22 | 0.683 | 98.3 | 11.330 [7.821, 16.414] |
| Estimated rate of annual AIDS-related deaths (per 100,000 population) | WCA | Age 15-19 | 2015 | PM | 22 | 0.682 | 98.3 | 11.330 [7.821, 16.414] |
| Estimated rate of annual AIDS-related deaths (per 100,000 population) | WCA | Age 15-19 | 2015 | DL | 22 | 0.671 | 98.2 | 11.331 [7.821, 16.415] |
| Estimated rate of annual AIDS-related deaths (per 100,000 population) | WCA | Age 15-19 | 2016 | REML | 22 | 0.696 | 98.2 | 10.888 [7.488, 15.834] |
| Estimated rate of annual AIDS-related deaths (per 100,000 population) | WCA | Age 15-19 | 2016 | PM | 22 | 0.696 | 98.2 | 10.888 [7.488, 15.834] |
| Estimated rate of annual AIDS-related deaths (per 100,000 population) | WCA | Age 15-19 | 2016 | DL | 22 | 0.715 | 98.2 | 10.888 [7.487, 15.833] |
| Estimated rate of annual AIDS-related deaths (per 100,000 population) | WCA | Age 15-19 | 2017 | REML | 22 | 0.706 | 98.1 | 10.355 [7.104, 15.094] |
| Estimated rate of annual AIDS-related deaths (per 100,000 population) | WCA | Age 15-19 | 2017 | PM | 22 | 0.705 | 98.1 | 10.355 [7.104, 15.094] |
| Estimated rate of annual AIDS-related deaths (per 100,000 population) | WCA | Age 15-19 | 2017 | DL | 22 | 0.793 | 98.3 | 10.353 [7.103, 15.091] |
| Estimated rate of annual AIDS-related deaths (per 100,000 population) | WCA | Age 15-19 | 2018 | REML | 22 | 0.704 | 97.9 | 9.689 [6.652, 14.114] |
| Estimated rate of annual AIDS-related deaths (per 100,000 population) | WCA | Age 15-19 | 2018 | PM | 22 | 0.702 | 97.9 | 9.689 [6.652, 14.114] |
| Estimated rate of annual AIDS-related deaths (per 100,000 population) | WCA | Age 15-19 | 2018 | DL | 22 | 0.820 | 98.2 | 9.689 [6.652, 14.112] |
| Estimated rate of annual AIDS-related deaths (per 100,000 population) | WCA | Age 15-19 | 2019 | REML | 22 | 0.690 | 97.6 | 9.089 [6.260, 13.196] |
| Estimated rate of annual AIDS-related deaths (per 100,000 population) | WCA | Age 15-19 | 2019 | PM | 22 | 0.688 | 97.6 | 9.089 [6.260, 13.196] |
| Estimated rate of annual AIDS-related deaths (per 100,000 population) | WCA | Age 15-19 | 2019 | DL | 22 | 0.752 | 97.8 | 9.089 [6.260, 13.196] |
| Estimated rate of annual AIDS-related deaths (per 100,000 population) | WCA | Age 15-19 | 2020 | REML | 22 | 0.677 | 97.3 | 8.367 [5.779, 12.115] |
| Estimated rate of annual AIDS-related deaths (per 100,000 population) | WCA | Age 15-19 | 2020 | PM | 22 | 0.676 | 97.3 | 8.367 [5.779, 12.115] |
| Estimated rate of annual AIDS-related deaths (per 100,000 population) | WCA | Age 15-19 | 2020 | DL | 22 | 0.709 | 97.4 | 8.367 [5.779, 12.115] |
| Estimated rate of annual AIDS-related deaths (per 100,000 population) | WCA | Age 15-19 | 2021 | REML | 22 | 0.661 | 96.9 | 7.653 [5.306, 11.037] |
| Estimated rate of annual AIDS-related deaths (per 100,000 population) | WCA | Age 15-19 | 2021 | PM | 22 | 0.658 | 96.9 | 7.652 [5.306, 11.037] |
| Estimated rate of annual AIDS-related deaths (per 100,000 population) | WCA | Age 15-19 | 2021 | DL | 22 | 0.717 | 97.1 | 7.654 [5.307, 11.038] |
| Estimated rate of annual AIDS-related deaths (per 100,000 population) | WCA | Age 15-19 | 2022 | REML | 22 | 0.637 | 96.5 | 6.858 [4.780, 9.837] |
| Estimated rate of annual AIDS-related deaths (per 100,000 population) | WCA | Age 15-19 | 2022 | PM | 22 | 0.636 | 96.5 | 6.857 [4.780, 9.837] |
| Estimated rate of annual AIDS-related deaths (per 100,000 population) | WCA | Age 15-19 | 2022 | DL | 22 | 0.673 | 96.7 | 6.858 [4.781, 9.838] |
| Estimated rate of annual AIDS-related deaths (per 100,000 population) | WCA | Age 15-19 | 2023 | REML | 22 | 0.638 | 96.3 | 6.053 [4.216, 8.690] |
| Estimated rate of annual AIDS-related deaths (per 100,000 population) | WCA | Age 15-19 | 2023 | PM | 22 | 0.637 | 96.3 | 6.053 [4.216, 8.690] |
| Estimated rate of annual AIDS-related deaths (per 100,000 population) | WCA | Age 15-19 | 2023 | DL | 22 | 0.667 | 96.5 | 6.054 [4.217, 8.691] |

*Continued on next p*

**Supplementary Table S4 (continued)**

| Indicator | Region | Age group | Year | Method | $k$ | $\tau^2$ | $I^2$ (%) | Pooled estimate [95% |
|---|---|---|---|---|---|---|---|---|
| Estimated rate of annual AIDS-related deaths (per 100,000 population) | WCA | Age 15-19 | 2024 | REML | 22 | 0.642 | 96.1 | 5.387 [3.748, 7.744] |
| Estimated rate of annual AIDS-related deaths (per 100,000 population) | WCA | Age 15-19 | 2024 | PM | 22 | 0.640 | 96.1 | 5.387 [3.747, 7.744] |
| Estimated rate of annual AIDS-related deaths (per 100,000 population) | WCA | Age 15-19 | 2024 | DL | 22 | 0.683 | 96.4 | 5.389 [3.749, 7.746] |

Notes: $k$ = contributing countries. Differences across estimators reflect alternative moment/likelihood choices for $\tau^2$; main text uses REML + Hartung–Knapp.
