## Supplementary Table S5 for "Place, gender, and uneven progress in pediatric and adolescent HIV across sub-Saharan Africa: a regional meta-analytic assessment (2000–2024)"

Table 1: Parametric bootstrap medians and 95% intervals for observed and required annualised rate of change (ARC); $B = 5{,}000$ draws per stratum. Back-transformation uses log/logit as appropriate.

| Region | Indicator | Age | Obs. ARC | Obs. ARC (bs, %/yr) [95% CI] | Req. ARC | Req. ARC (bs, %/yr) [95% CI] |
|---|---|---|---|---|---|---|
| ESA | Incidence (per 1,000) | Age 0–14 | -8.8 | -8.8 (-13.9, -3.6) | -15.6 | -15.5 (-25.8, -3.5) |
| ESA | Incidence (per 1,000) | Age 15–19 | -5.4 | -5.4 (-13.2, 2.8) | -22.5 | -22.4 (-36.1, -5.3) |
| ESA | MTCT (%) | Age 0–4 | -6.7 | -6.7 (-10.0, -3.5) | -20.0 | -20.0 (-26.0, -13.0) |
| ESA | Mortality (per 100,000) | Age 0–14 | -10.4 | -10.5 (-15.4, -5.3) | -11.9 | -11.8 (-22.6, 0.8) |
| ESA | Mortality (per 100,000) | Age 10–19 | -6.5 | -6.6 (-12.6, -0.3) | -20.3 | -20.2 (-31.5, -6.8) |
| ESA | Mortality (per 100,000) | Age 15–19 | -3.2 | -3.2 (-10.2, 3.9) | -26.6 | -26.5 (-37.7, -12.5) |
| WCA | Incidence (per 1,000) | Age 0–14 | -7.4 | -7.4 (-11.0, -3.7) | -18.6 | -18.5 (-25.6, -10.5) |
| WCA | Incidence (per 1,000) | Age 15–19 | -6.6 | -6.6 (-12.0, -1.2) | -20.1 | -20.1 (-30.0, -8.3) |
| WCA | MTCT (%) | Age 0–4 | -3.6 | -3.6 (-5.0, -2.4) | -25.7 | -25.7 (-27.9, -23.3) |
| WCA | Mortality (per 100,000) | Age 0–14 | -9.0 | -9.1 (-12.5, -5.7) | -15.0 | -15.0 (-21.9, -7.0) |
| WCA | Mortality (per 100,000) | Age 10–19 | -6.6 | -6.7 (-10.0, -3.3) | -20.0 | -20.0 (-26.2, -12.9) |
| WCA | Mortality (per 100,000) | Age 15–19 | -4.4 | -4.4 (-7.9, -0.8) | -24.4 | -24.4 (-30.5, -17.4) |

pt = point estimate from pooled 2010 & 2024 levels; bs = parametric bootstrap median over $B = 5{,}000$ draws on the transformed scale; pooling used inverse-variance random-effects with HK/SJ; required ARC targets a reduction to 10% of the 2010 level by 2030 (6-year window from 2024).
